# Parent-Reported Child Mania Rating Scale: Reliability, Validity, and Norms in a Nationally Representative US Sample

**DOI:** 10.64898/2026.10.12.26365308

**Authors:** Marissa B. McClellan, Cristian Alcaino, Yinuo Liu, Phoebe A. Rodda, Jennifer K. Youngstrom, Robert L. Findling, Eric A. Youngstrom

## Abstract

**Objective:** To evaluate the psychometric properties, factor structure, validity, and age-and sex-specific norms of the 21-and 10-item parent-report Child Mania Rating Scale (CMRS-P) in a nationally representative U.S. sample of youth aged 6–18 years.

**Method:** Parents of 2,229 youth aged 6–18 years [weighted *M*(*SD*)=12.0(3.3) years; 45.9% female] completed the CMRS-P and related measures between July 2025 and March 2026. Data were weighted to represent U.S. demographics. A clinical comparison sample included parents of 522 youth seeking outpatient mental health services. Complete-case analyses evaluated reliability, factor structure, item response theory, measurement equivalence, retest stability, and convergent and discriminant validity. GAMLSS estimated age-and sex-specific norms.

**Results:** Weighted mean scores were 8.5(*SD*=9.0) for CMRS-P-21 and 4.0(4.3) for CMRS-P-10. Internal consistency was high (ω=.97/.93), and forms correlated.95. A general severity factor explained most reliable variance (ω*H*=.94). Three items differed in response thresholds across samples, with an average impact below one point on expected total scores. Both forms correlated strongly with attention problems, suicidality, depression, externalizing symptoms, and emotion dysregulation (*r=*.51–.63 for CMRS-P-21) and negligibly with pubertal status. Reliable-change thresholds were 14.5 (CMRS-P-21) and 6.8 (CMRS-P-10) points. Weighted GAMLSS CMRS-P-21 95th percentiles declined with age in males (30–23) and increased in females (22–30), with wide bootstrap intervals at age extremes; the female trend was not replicated without weights.

**Conclusions:** Findings support both CMRS-P forms for assessing youth manic symptoms. The first nationally representative U.S. norms provide age-and sex-adjusted benchmarks for interpreting scores and informing further clinical assessment.

---

Pediatric Bipolar Disorder (PBD) is a serious condition that has gained increasing recognition over the past three decades (Goldstein et al., 2017; E. A. Youngstrom et al., 2008). PBD is characterized by episodes of [hypo]mania, with elevated or irritable mood and abnormally and persistently increased activity or energy (American Psychiatric Association [APA], 2022; Saxena et al., 2020; World Health Organization [WHO], 2019). As in adults with BD, grandiosity, decreased need for sleep, pressured speech, racing thoughts, and distractibility are common indicators of [hypo]mania, and depressive episodes are frequently observed as well (American Psychiatric Association, 2022; Freeman et al., 2009; Van Meter et al., 2016; World Health Organization, 2019). However, youth with PBD may be more likely to experience mixed states (Saxena et al., 2020), higher recurrence (E. A. Youngstrom et al., 2008), lower quality of life (Freeman et al., 2009; McGinty et al., 2023; Morton et al., 2021), increased risk of suicide (Hauser et al., 2013), more severe mood symptoms (E. A. Youngstrom et al., 2008), psychiatric comorbidity (Fahrendorff et al., 2023), and fewer periods of remission over time (Birmaher, 2007; Lee et al., 2020). When PBD goes untreated, risk of these adverse outcomes is exacerbated and youth often experience a progressive illness course characterized by treatment resistance, impairment across psychosocial domains, and increased risk of hospitalization and suicide (Joslyn et al., 2016; Wozniak et al., 2025; E. A. Youngstrom et al., 2009).

Despite this, adults and youths with BD experience impairing symptoms for an average of 5 to 10 years before obtaining a diagnosis (Keramatian et al., 2022; Marchand et al., 2006; Scott et al., 2022), with greater diagnostic and treatment delays being observed in cases with early symptom onset (Berk et al., 2007; Cirone et al., 2021; Hafeman et al., 2016; Hirschfeld et al., 2003). As such, there is a clear need for tools that can efficiently identify symptoms of PBD in both clinical and nonclinical populations (Goldstein et al., 2017; E. A. Youngstrom, 2026). While clinical interviews are best for diagnosing PBD (e.g., Kaufman et al., 2016), they are often inaccessible as they require trained interviewers and substantial time to adequately administer (Townsend et al., 2020). Most providers also have received little formal training about bipolar disorder, compounding the challenges for early and accurate recognition. Questionnaires have shown classification validity, with parent ratings showing the largest effect sizes, followed by youth and teacher report (Alcaino et al., 2024; E. A. Youngstrom et al., 2015).

The Child Mania Rating Scale (CMRS-P-21) is one of the three top performing scales in terms of discriminating bipolar disorder from other diagnoses. The CMRS-P-21 has been adapted into a 10-item short-form (Henry et al., 2008; CMRS-P-10), tested in many different demographic subpopulations (Westerholm, 2009), translated into numerous languages (Meyer et al., 2024; Serrano et al., 2011), and applied to a wide variety of cultural contexts (Mendez et al., 2020; Rucklidge et al., 2019; Taylor et al., 2022; Verstraeten et al., 2012). It includes items intended to identify manic symptoms in younger age groups and has been tested in children as young as five years of age (Westerholm, 2009). Across samples, the internal consistency of the CMRS-P is good (De Back et al., 2023; McArthur et al., 2017) to excellent (Gruhn et al., 2021; Henry et al., 2008; Kiker, 2010; MacPherson et al., 2016; Pavuluri et al., 2006; Serrano et al., 2013). Preliminary analyses (i.e., Henry et al., 2008; Pavuluri et al., 2006) reported excellent 1-week test-retest reliability (0.96-0.97) and criterion validity for differentiating PBD from both ADHD (AUC=0.91) and healthy controls (AUC=0.97), albeit in distilled bipolar versus healthy control case comparisons that increase the between-case variance and maximize these statistics (E. Youngstrom et al., 2006; E. A. Youngstrom et al., 2015). Importantly, it also shows treatment sensitivity (Findling et al., 2011; MacPherson et al., 2016; West et al., 2011).

The performance of the CMPS-P has not yet been evaluated in a large, nationally representative, non-treatment seeking population. This is an especially important gap because the symptoms of hypomania and mania overlap with behaviors that would be developmentally typical, or attributable to other more common conditions such as attention-deficit/hyperactivity disorder (ADHD) or unipolar depression (Fahrendorff et al., 2023). There are a variety of instruments that do have norms, and they most often adjust for age and sex of the youth because these moderate what is typical behavior (e.g., ASEBA, Achenbach & Rescorla, 2001; BASC, Reynolds & Kamphaus, 2015).

To address these limitations, the present analysis assessed the score distribution, factor structure, reliability, item response characteristics, and validity of both the full-length and brief versions of the CMRS-P in a nationally representative sample of US youths. We also performed secondary analyses on a clinical dataset where primary caregivers completed the CMRS-P as part of a research battery enrolling families seeking outpatient mental health services for the youth. We hypothesized that mean scores would be lower in the general population than in the clinical sample, with positively skewed distributions in both. We anticipate that the psychometric performance of both the full-length and 10-item scales will be slightly poorer in the general population than the clinical sample, but it will still be adequate when compared to commonly used benchmarks for evaluating pediatric symptom screeners. Finally, we expect that the majority of variance will be accounted for by a single factor, as in prior analyses (Henry et al., 2008; Pavuluri et al., 2006), although there might be multiple underlying group factors, as found in other measures of manic symptoms (Depue et al., 1981; Gamma et al., 2013). We test for differential item functioning across sex, age, and clinical versus general population status, and we report continuous norms using a GAMLSS model to model nonlinear age and sex effects using an appropriate distributional model.

## Methods

This section summarizes the design, measures, and primary analyses. Expanded procedures, measure descriptions, scoring specifications, and analytic diagnostics appear in the corresponding Methods subsections of the Supplemental Materials.

### Participants and Procedure

The BENCHMARK 2025 Project recruited parents of youths aged 6–18 years through YouGov’s online panel to develop national norms and evaluate pediatric mental and behavioral health measures (E. Youngstrom et al., 2025). Of 2,252 parent respondents, 2,229 had complete CMRS-P-21 data and formed the analytic sample for both CMRS-P versions. Participants represented all 50 states and the District of Columbia; Table 1 summarizes demographics. Data were collected between July 11, 2025 and March 7, 2026. Details appear under Participants, Procedure, and Demographics and Eligibility in the Supplemental Materials.

**Table 1.** Sample Demographics.

|  | *General Population*<br>*N = 2229* | *Clinical*<br>*N = 522* |
| --- | --- | --- |
| **Youth Characteristics** |  |  |
| 6 to 11 y.o. sample, *n* (%) | 993 (39.0%) | 4–11: 295 (56.5%) |
| Child age, *M* (*SD*) | 8.43 (1.67) | 8.03 (2.08) |
| 12 to 18 y.o. sample, *n* (%) | 1236 (61.0%) | 12–18: 227 (43.5%) |
| Child age, *M* (*SD*) | 14.26 (1.67) | 14.00 (1.64) |
| Full sample age, *M* (*SD*) | 11.98 (3.30) | 10.63 (3.52) |
| Sex, *n* (%) |  |  |
| Male | 1171 (54.1%) | 318 (60.9%) |
| Female | 1058 (45.9%) | 204 (39.1%) |
| Race %<sup>a</sup> |  |  |
| White | 74.4% | 23.9% |
| Other | 24.5% | 76.1% |
| Prefer not to say | 1.1% | 0.0% |
| Ethnicity |  |  |
| Hispanic or Latino | 24.1% | 2.5%† |
| Not Hispanic or Latino | 70.9% | 97.5%† |
| Other | 3.1% | — |
| Prefer not to say | 1.9% | — |
| **Parent Characteristics** |  |  |
| Parent age, *M* (*SD*) | 42.85 (7.79 years) | 39.48 (11.03 years) |
| Range (Median) | 23 – 81 (42 years) | 21.5 – 80.6 (37.7 years) |
| Gender, *n* (%) |  |  |
| Male | 765 (47.6%) | 33 (6.4%) |
| Female | 1445 (51.5%) | 479 (93.6%) |
| Non-binary or Other | 19 (0.9%) | — |
| Race, *n* (%) |  |  |
| White | 1678 (66.1%) | 137 (27.1%) |
| Black or African American | 195 (10.6%) | 345 (68.3%) |
| Asian | 83 (4.7%) | 0 (0.0%)<sup>a</sup> |
| Native American | 38 (2.2%) | 1 (0.2%) |
| Native Hawaiian or Pacific Islander | 6 (0.6%) | see Asian<sup>a</sup> |
| Other | 104 (8.9%) | 10 (2.0%) |
| Multi-racial | 91 (4.0%) | — |
| Ethnicity – Hispanic or Latino, *n* (%) | 354 (24.4%) | 12 (2.4%) |
| Education, *n* (%) |  |  |
| High school or less | 446 (33.0%) | 286 (56.4%) |
| Some college | 458 (17.5%) | 167 (32.9%) |
| 2-year, 4-year, or post graduate degree | 1325 (49.5%) | 54 (10.7%) |
| Marital Status – Married, *n* (%) | 1483 (65.4%) | 151 (29.7%) |
| Family Income, *n* (%) |  |  |
| < \$10,000 | 94 (4.4%) | ~40% |
| \$10,000 – 39,999 | 367 (18.8%) | ~47% |
| \$40,000 – 69,999 | 467 (21.0%) | ~5% |
| \$70,000 – 99,999 | 398 (18.2%) | ~4% |
| > \$100,000 | 796 (33.0%) | ~4% |
| U.S. Region, *n* (%) |  |  |
| Northeast | 419 (15.9%) | 0 (0%) |
| Midwest | 493 (19.4%) | 522 (100%) |
| South | 844 (39.5%) | 0 (0%) |
| West | 473 (25.2%) | 0 (0%) |

*Note.* <sup>a</sup>The clinical sample coded Asian and Pacific Islander as one combined category. Response bins not identical for incomes across samples, so clinical sample shows best approximations.

The Nationwide Children’s Hospital Institutional Review Board designated the BENCHMARK procedures Not Human Subjects Research because investigators had no direct participant contact and received no individual identifiers.

The clinical comparison sample comprised 522 parents with complete CMRS-P-21 data from 828 families enrolled in an NIMH-funded outpatient study in Ohio (R01 MH066647). Youths were aged 4–18 years. The CMRS-P was added during the third year through an IRB-approved modification; earlier enrollees did not receive it. Trained raters administered the KSADS-PL Plus to parents and youths and established diagnoses through longitudinal expert evaluation of all data. This regional, unweighted sample provided a clinical reference for factor structure, item functioning, and clinical-significance benchmarks. It did not include the other BENCHMARK measures, precluding direct comparisons of criterion correlations. Diagnostic procedures and supporting references appear under Clinical Comparison Sample in the Supplemental Materials.

### Measures

The CMRS-P-21 assesses parent-observed manic symptoms over the past month. Items are rated from 0 (Never/Rare) to 3 (Very Often), with higher summed scores indicating greater symptom severity (American Psychiatric Association, 1994; Pavuluri et al., 2006). We evaluated both the 21-item form and the established 10-item short form (CMRS-P-10). In BENCHMARK, the standard 21-item order was modified so that the 10 short-form items appeared first, to approximate item presentation of the standalone brief-form. The clinical study used the standard 21-item order, with short-form scores calculated from the corresponding items within the full scale. Item content and brief-form membership are provided in Supplemental Table S1; administration details and validation references appear under “Assessment Battery” in the Supplemental Materials.

Convergent and discriminant validity indicators covered depression (VQIDS-P-A), emotion dysregulation (EDI reactivity and dysphoria), internalizing, externalizing, and attention problems (PSC-17), ADHD symptoms and performance (VADPRS), suicide risk (CHRT-16), family mood disorder history (FIRM), sleep-related impairment (PROMIS-SRI), quality of life (NQLS), pubertal development (PDS), and global parental concern (N-Gauge). Parents also reported school functioning, diagnoses, and treatment history. Age, sex, and pubertal status provided developmental correlates. The supplement’s BENCHMARK Convergent and Discriminant Validity Indicators subsection provides measure descriptions, response options, scoring, and references, including parent-report adaptations.

### Statistical Analyses

Analyses used *R*, with complete CMRS-P-21 cases retained for both forms and no prorating.

### Weighting

General-population analyses applied YouGov weights; clinical analyses were unweighted. The general-population analytic sample had a Kish design effect of 1.93 and an effective sample size of approximately 1,156. Counts are unweighted; general-population means and percentages are weighted. “Statistical Analyses” in the Supplemental Materials describes model-specific weighting procedures, effective-sample-size adjustments, and unweighted comparisons.

Weighted estimation included polychoric correlations, weighted least squares, mean-and variance-adjusted (WLSMV) factor models, survey-weighted item response models, and prior weights in continuous-norming models. Limited-information model fit and quantile-residual diagnostics were evaluated using repeated pseudo-populations sampled with probability proportional to the weights at the effective sample size. The supplement explains these procedures so that model fit and uncertainty can be interpreted in relation to the unequal sampling weights.

### Factor Structure and Item Response Theory

The clinical sample anchored evaluation of factor structure because it included a broader range of clinically significant mood symptoms. Ordered-categorical analyses used polychoric correlations and WLSMV estimation. We evaluated correlated-factor, bifactor, and higher-order confirmatory models, with a priori clinical exploratory analyses informing assignment to Elated Mood, Activity, Irritable Mood, and Strange Thoughts and Behaviors factors. Structural equivalence was assessed with a joint multigroup configural model. Fit interpretation emphasized the comparative fit index (CFI), Tucker–Lewis index (TLI), and standardized root-mean-square residual (SRMR); root-mean-square error of approximation (RMSEA) was reported descriptively. Models with persistent convergence problems, out-of-bounds standardized loadings, or negative residual variances were rejected.

General-factor dominance and the distinctiveness of group factors were evaluated using explained common variance, omega-hierarchical, omega-hierarchical-subscale, *H*, and the proportion of uncontaminated correlations. The supplement’s Factor Structure subsection gives model specifications and interpretive criteria; Supplemental Tables S5–S9 provide supporting structural results.

Graded response models evaluated the 21-and 10-item forms separately in each sample, with supplementary factor-level calibrations and bifactor models when unidimensional fit was poor. Diagnostics addressed overall fit, item fit, local dependence, and person fit. Clinical item-fit deviations were compared with weight-proportional general-population subsamples of the same size to distinguish substantive differences from sample-size effects. We estimated empirical reliability of latent scores and related them to expected raw totals. The supplement’s Item Response Theory subsection specifies diagnostic thresholds and calibration procedures; Supplemental Tables S10–S14 report fit and item parameters.

### Measurement Equivalence Between Samples

We screened differential item functioning (DIF) with ordinal logistic regression (R² change ≥.02, α=.01), then tested configural, metric, and scalar invariance using multiple-group graded response models. A CFI decrease greater than.010 indicated noninvariance. When full scalar invariance failed, items not flagged for DIF served as anchors in a partial-scalar model; flagged parameters were freely estimated. The general-population latent mean and variance were fixed at 0 and 1, and clinical values were estimated. DIF was also screened by child sex and across three child-age strata (6–9, 10–12, and 13–18 years) in the general-population sample with the same criteria, with and without the weights (Supplemental Materials).

This linking assumes that anchor items function equivalently across samples; anchor noninvariance could bias latent mean differences and expected-score comparisons. Differential test functioning was summarized across θ = −3 to 3 and weighted by the clinical latent distribution, then compared with reliable-change and minimal important difference thresholds. Full linking specifications and effect-size calculations appear under Measurement Equivalence Between Samples in the Supplemental Materials.

### Reliability and Validity

Internal consistency was summarized with mean inter-item correlations, Cronbach’s alpha (α), and McDonald’s omega (ω). Validity analyses used weighted Pearson (*r*) correlations among respondents complete on both CMRS-P forms and all non-sex-specific criterion scales. We compared the two forms’ correlations using Steiger’s test for dependent, overlapping correlations, using Kish’s effective sample size for *N*, and summarized differences with Cohen’s *q* effect size. We anticipated strong associations with depression, internalizing and externalizing symptoms, and family mood-disorder history; at least moderate associations with sleep and attention problems; and smaller developmental associations. Scoring conventions and statistical details appear under Reliability and Validity in the Supplemental Materials, with supporting results in Supplemental Tables S16–S18.

### Retest Stability and Clinical Benchmarks

A random subset of 167 parents completed the CMRS-P again approximately two to three weeks later. Retest dependability used the absolute-agreement intraclass correlation, ICC(A,1), with baseline weights (effective *N*=81) and confidence intervals obtained by bootstrapping pairs. The baseline standard deviation and ICC yielded standard errors of measurement and reliable-change thresholds at 95% and 90% confidence. Omega-based thresholds were calculated for comparison but exclude time-varying error.

The distribution-based minimal important difference was 0.5 *SD*. Clinical-significance benchmarks followed Jacobson and Truax’s Methods A (clinical mean minus 2 *SD*), B (general-population mean plus 2 *SD*), and C (the weighted midpoint between means)(Jacobson & Truax, 1991). Formulas, assumptions, and weighting details appear under Retest Stability and Clinical Benchmarks in the Supplemental Materials; Table 2 presents the primary estimates.

**Table 2.**
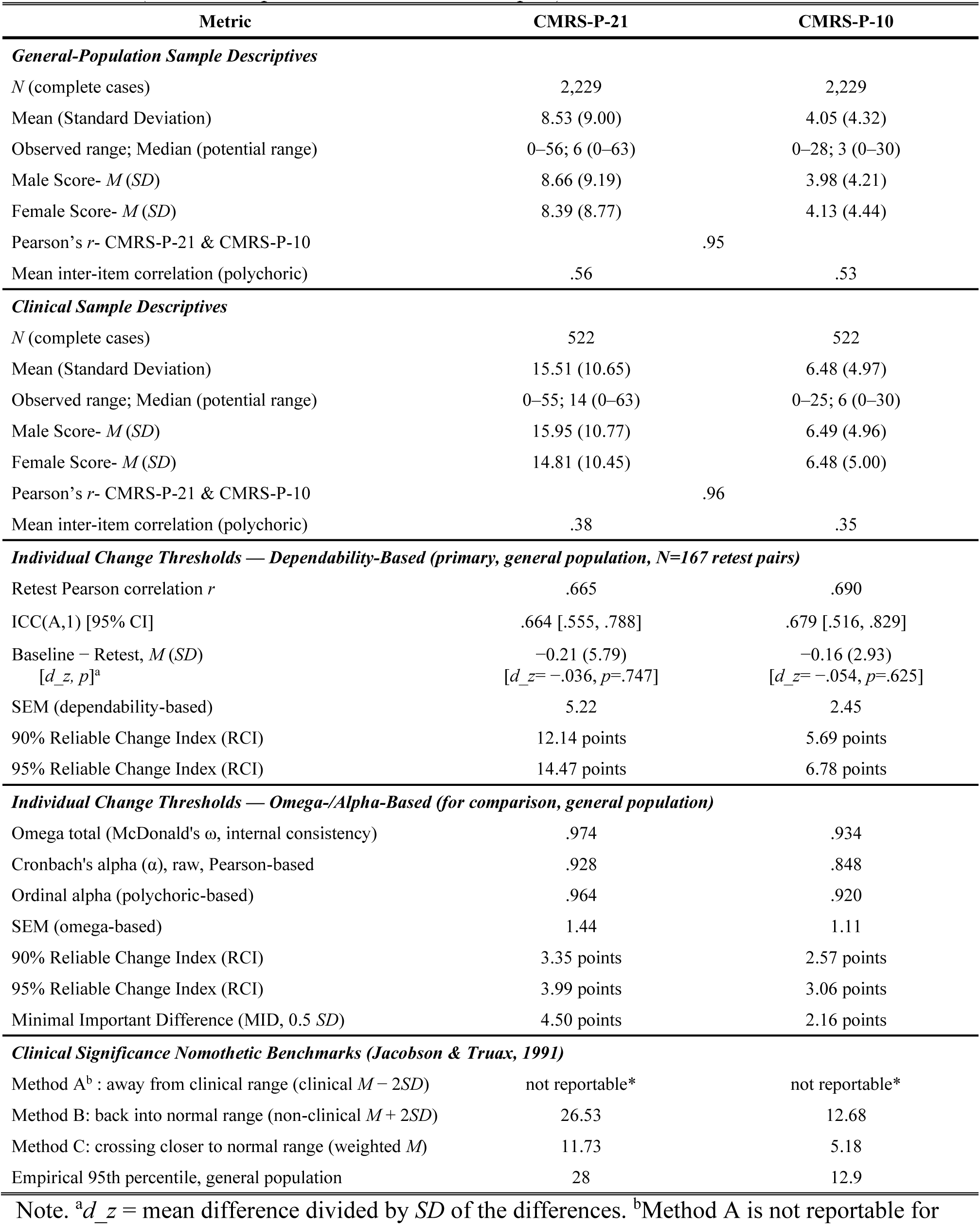
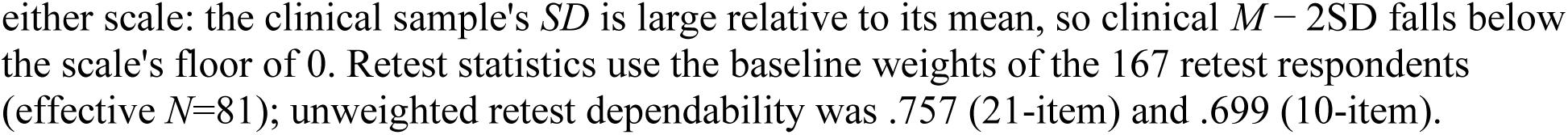
Descriptive Statistics, Reliability, and Clinical Significance Benchmarks: CMRS-P-21 and CMRS-P-10 (General-Population and Clinical Samples)

| Metric | CMRS-P-21 | CMRS-P-10 |
| --- | --- | --- |
| **General-Population Sample Descriptives** |  |  |
| *N* (complete cases) | 2,229 | 2,229 |
| Mean (Standard Deviation) | 8.53 (9.00) | 4.05 (4.32) |
| Observed range; Median (potential range) | 0–56; 6 (0–63) | 0–28; 3 (0–30) |
| Male Score- *M* (*SD*) | 8.66 (9.19) | 3.98 (4.21) |
| Female Score- *M* (*SD*) | 8.39 (8.77) | 4.13 (4.44) |
| Pearson's *r*- CMRS-P-21 & CMRS-P-10 | .95 |  |
| Mean inter-item correlation (polychoric) | .56 | .53 |
| **Clinical Sample Descriptives** |  |  |
| *N* (complete cases) | 522 | 522 |
| Mean (Standard Deviation) | 15.51 (10.65) | 6.48 (4.97) |
| Observed range; Median (potential range) | 0–55; 14 (0–63) | 0–25; 6 (0–30) |
| Male Score- *M* (*SD*) | 15.95 (10.77) | 6.49 (4.96) |
| Female Score- *M* (*SD*) | 14.81 (10.45) | 6.48 (5.00) |
| Pearson's *r*- CMRS-P-21 & CMRS-P-10 | .96 |  |
| Mean inter-item correlation (polychoric) | .38 | .35 |
| **Individual Change Thresholds — Dependability-Based (primary, general population, *N*=167 retest pairs)** |  |  |
| Retest Pearson correlation *r* | .665 | .690 |
| ICC(A,1) [95% CI] | .664 [.555, .788] | .679 [.516, .829] |
| Baseline – Retest, *M* (*SD*)<br>[*d<sub>z</sub>*, *p*]<sup>a</sup> | –0.21 (5.79)<br>[*d<sub>z</sub>* = –.036, *p* = .747] | –0.16 (2.93)<br>[*d<sub>z</sub>* = –.054, *p* = .625] |
| SEM (dependability-based) | 5.22 | 2.45 |
| 90% Reliable Change Index (RCI) | 12.14 points | 5.69 points |
| 95% Reliable Change Index (RCI) | 14.47 points | 6.78 points |
| **Individual Change Thresholds — Omega-/Alpha-Based (for comparison, general population)** |  |  |
| Omega total (McDonald's $\omega$, internal consistency) | .974 | .934 |
| Cronbach's alpha ($\alpha$), raw, Pearson-based | .928 | .848 |
| Ordinal alpha (polychoric-based) | .964 | .920 |
| SEM (omega-based) | 1.44 | 1.11 |
| 90% Reliable Change Index (RCI) | 3.35 points | 2.57 points |
| 95% Reliable Change Index (RCI) | 3.99 points | 3.06 points |
| Minimal Important Difference (MID, 0.5 *SD*) | 4.50 points | 2.16 points |
| **Clinical Significance Nomothetic Benchmarks (Jacobson & Truax, 1991)** |  |  |
| Method A<sup>b</sup> : away from clinical range (clinical *M* – 2*SD*) | not reportable* | not reportable* |
| Method B: back into normal range (non-clinical *M* + 2*SD*) | 26.53 | 12.68 |
| Method C: crossing closer to normal range (weighted *M*) | 11.73 | 5.18 |
| Empirical 95th percentile, general population | 28 | 12.9 |

Note. <sup>a</sup>*d<sub>z</sub>* = mean difference divided by *SD* of the differences. <sup>b</sup>Method A is not reportable for
either scale: the clinical sample's $SD$ is large relative to its mean, so clinical $M - 2SD$ falls below the scale's floor of 0. Retest statistics use the baseline weights of the 167 retest respondents (effective $N=81$ ); unweighted retest dependability was .757 (21-item) and .699 (10-item).

### Continuous Norming

We estimated age-and sex-referenced percentiles using generalized additive models for location, scale, and shape (GAMLSS). Weighted cross-validation and Bayesian information criterion (BIC) guided selection among polynomial age and sex specifications; this post hoc rule selected linear age, sex, and their interaction. With this mean structure fixed, discrete distributions were compared using BIC and residual diagnostics, prioritizing diagnostics when they disagreed. Bootstrapping quantified percentile uncertainty. Sensitivity analyses examined unweighted, cubic, covariate-standardized, and flexible-spline alternatives. Full selection rules, weighting, bootstrap procedures, and sensitivity specifications appear under “Continuous Norming” in the Supplemental Materials; Table S19, accompanying model-comparison tables, and Figures S1–S2 provide supporting comparisons and diagnostics.

## Results

Of 2,252 potential respondents, 2,229 parents completed all 21-items of the CMRS-P-21 and 2,240 completed all 10-items of the CMRS-P-10, suggesting that the difference in response burden was negligible. All analyses used the 2,229 complete cases and, for the general-population sample, the YouGov weights (design effect=1.93). Table 1 describes participant characteristics (counts are unweighted and percentages are weighted), and Table 2 presents mean CMRS-P scores, reliability metrics, and clinical significance benchmark information. Weighted total scores averaged 8.53 (*SD=*9.00; range=0–56; possible range=0–63) for the CMRS-P-21 and 4.05 (*SD=*4.32; range=0–28; possible range=0–30) for the CMRS-P-10. McDonald’s omega was.97 for the 21-item scale and.93 for the 10-item scale; other reliability estimates are reported in Table 2 for comparability with prior work. The CMRS-P-21 and CMRS-P-10 correlated.95. Figures 1a and 1b show score distributions in the general-population and clinical samples, and Table S1 reports item means and *SD*s. Differences in item averages all small to negligible, with a few reaching statistical significance (supplemental materials and Table S4 and S5).

**Figure 1.**
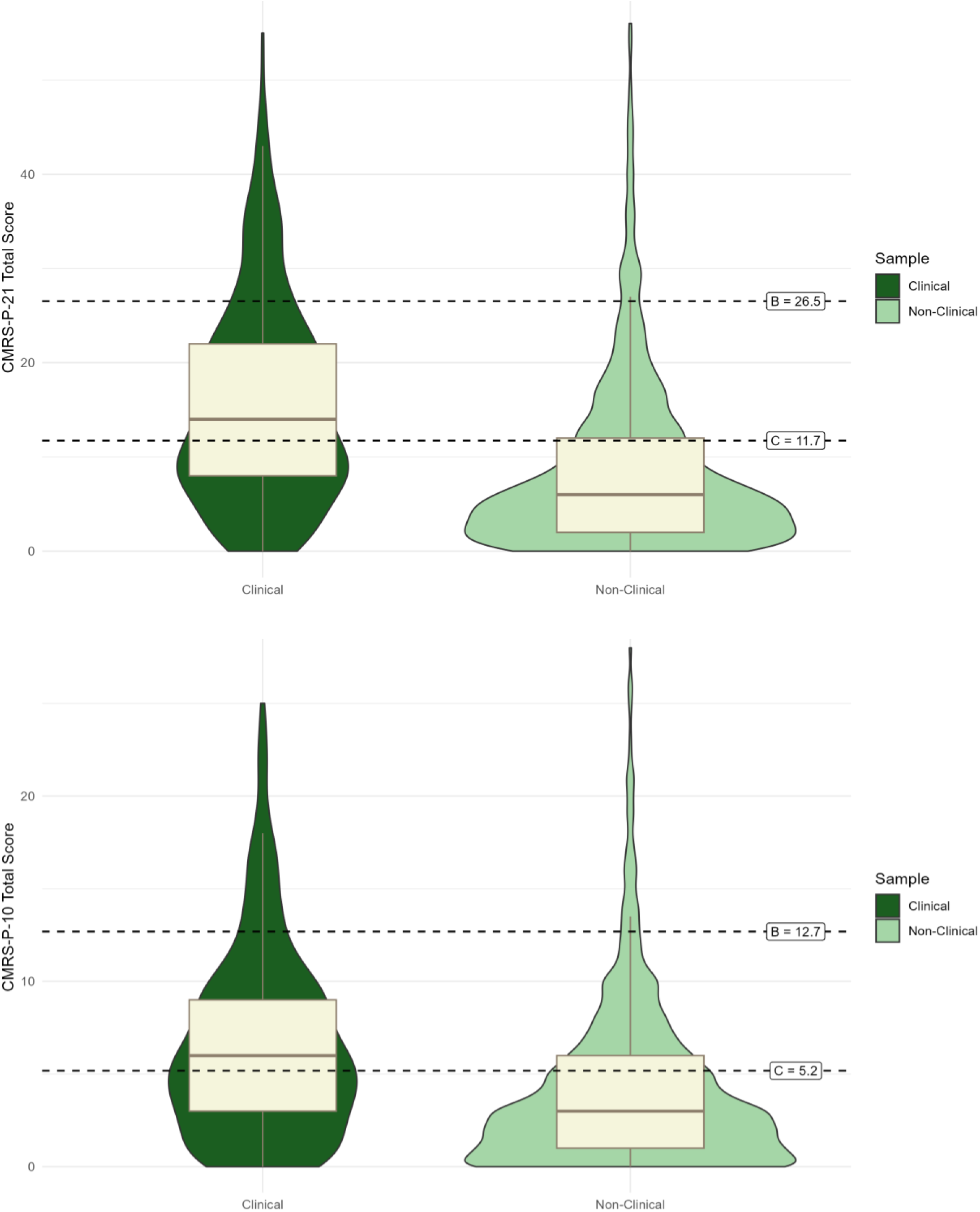
Total Score Distribution, General Population vs. Clinical and Nomothetic Benchmarks for CMRS-P-21 (top) and CMRS-P-10 (bottom) (non-clinical weighted to be nationally representative) Note. Dashed lines: B = non-clinical M + 2 SD; C = weighted M (clinical/non-clinical) (Jacobson & Truax Methods B and C, as in *Table 2*). Method A (away from the clinical range, clinical M − 2 SD) is not shown because it falls below the scale floor of 0.

### Factor Structure

Factor analyses evaluated the number and organization of constructs underlying the CMRS-P items. The clinical sample showed four moderately to highly correlated factors, well explained by a second-order factor. A conventional bifactor model was inadmissible because the Elated Mood factor correlated approximately 1.0 with the general factor, motivating the S-1 specification. A one-factor model fit acceptably in the general-population sample but poorly in the clinical sample. The bifactor-(S-1) model, with Elated Mood defining the general factor, was the best-fitting admissible representation and fit well in both samples (clinical: CFI=.993, TLI=.992, RMSEA=.043, SRMR=.063; weighted general population: CFI=.995, TLI=.994, RMSEA=.030, SRMR=.052; Figure 2; Table S5).

**Figure 2.**
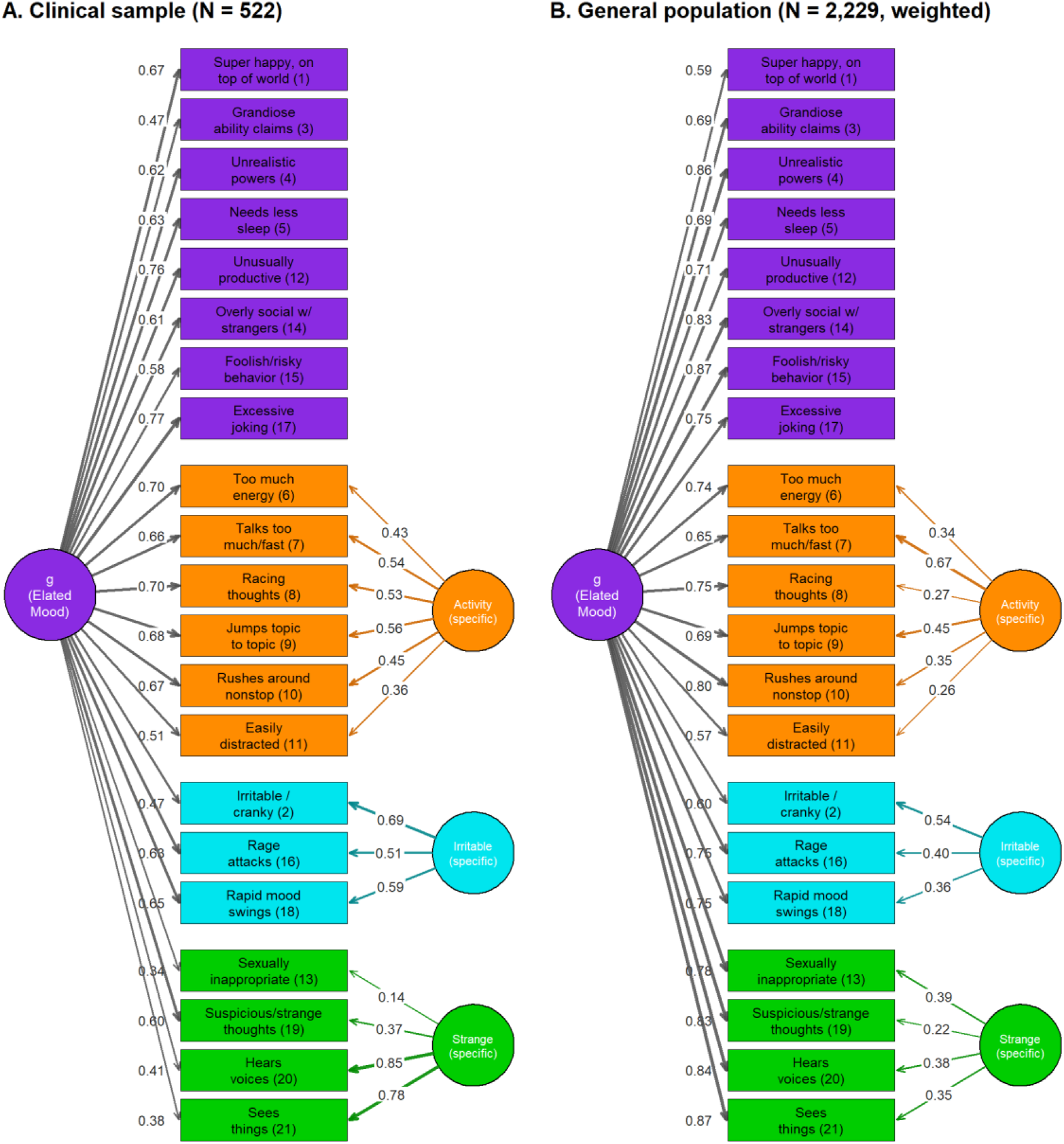
**Bifactor-(S-1) Model with Elated Mood as the Reference (General) Factor: Clinical and General-Population Samples** *Note*. Standardized path diagrams of the bifactor-(S-1) model in the clinical sample (Panel A, *N*=522, unweighted) and the general-population sample (Panel B, *N*=2,229, weighted coefficients). All 21 items load on a general factor (g) defined by the Elated Mood items, which serve as the reference domain; Activity, Irritable Mood, and Strange Thoughts and Behaviors are orthogonal specific factors (each item loads on g and on at most one specific factor). Numbers are standardized loadings. Fit statistics are in Table S5.

Multigroup CFA supported configural and metric invariance and, under the stated joint fit-change convention, scalar invariance (Table S6). For the S-1 model, imposing scalar constraints changed CFI by −.009, within the.010 tolerance, although the RMSEA increase of.016 was borderline. Freeing the three items flagged by item response theory (IRT) brought both changes within their respective tolerances (ΔCFI=−.006, ΔRMSEA=.011). Thus, conventional CFA criteria supported comparable measurement across samples while the more focused IRT analyses below detected item-level DIF.

Measurement invariance does not necessarily imply identical factor variances. In the scalar higher-order model, disturbance variances—the portions of first-order factor variance not explained by the general factor—were larger in the clinical sample: Activity,.822 versus.438; Irritable Mood,.585 versus.232; and Strange Thoughts and Behaviors,.438 versus.053 (1.9, 2.5, and 8.3 times larger, respectively; Table 3). Elated Mood had essentially no disturbance variance in either sample. General-factor variances were similar (1.333 versus 1.457). The larger clinical disturbances account for lower standardized second-order loadings despite equal unstandardized loadings. Constraining disturbance variances to equality changed CFI by only −.003, illustrating that global fit-change criteria can overlook these differences.

**Table 3.** First-Order Disturbance Variances and Standardized Second-Order Loadings by Sample: Scalar Higher-Order Model, Clinical vs. General Population.

| Factor | Disturbance variance |  |  | Standardized second-order loading |  |
| --- | --- | --- | --- | --- | --- |
|  | Clinical | General population | Ratio (clinical ÷ general) | Clinical | General population |
| *Weighted (general population weighted; clinical unweighted)* |  |  |  |  |  |
| Elated Mood (8 items) | -.003 | .016 | — | 1.00 | .99 |
| Activity (6 items) | .822 | .438 | 1.9 | .79 | .88 |
| Irritable Mood (3 items) | .585 | .232 | 2.5 | .71 | .86 |
| Strange Thoughts & Behaviors (4 items) | .438 | .053 | 8.3 | .63 | .93 |
| General factor variance (for reference) | .333 | 1.457 | .9 |  |  |
| *Unweighted* |  |  |  |  |  |
| Elated Mood (8 items) | -.004 | .018 | — | 1.00 | .99 |
| Activity (6 items) | .801 | .421 | 1.9 | .79 | .87 |
| Irritable Mood (3 items) | .588 | .288 | 2.0 | .71 | .82 |
| Strange Thoughts & Behaviors (4 items) | .443 | .073 | 6.1 | .63 | .90 |
| General factor variance (for reference) | 1.299 | 1.343 | 1.0 |  |  |

**Note.** Estimates are from the multiple-group second-order model in which item thresholds, first-order loadings, second-order loadings and item intercepts were constrained equal across samples (scalar model; WLSMV, theta parameterization; clinical sample as reference; $N = 522$ clinical, $N = 2,229$ general population). Disturbance variance is the variance of each first-order factor that the general (second-order) factor does not explain; it and the general-factor variance were free to differ by sample. Because the unstandardized second-order loadings are equal across samples by constraint, the differences in standardized loadings arise from differences in factor variances, chiefly the larger disturbance variances in the clinical sample. Ratios are clinical divided by general-population variance; none is shown for Elated Mood, whose disturbance variance is essentially zero in both samples (a small negative estimate in the clinical sample) because that factor is nearly identical to the general factor. Constraining the four disturbance variances to be equal across samples cost little in overall fit ( $\Delta CFI = -0.003$ weighted, $-0.002$ unweighted; $\Delta RMSEA = +0.004, +0.003$ ), so the conventional fit-index cutoffs are not sensitive to these differences.

### Item Response Theory

Essential unidimensionality tests supported giving the total score greatest interpretive weight, especially in the general population. The general factor accounted for most common variance (ECV=.857; omega-hierarchical=.936), and no group factor met all distinctiveness criteria (Table S9). The clinical sample retained a strong general factor but more specific-factor variance (ECV=.66; omega-hierarchical=.85). Accordingly, single-factor graded response models fit the general-population data better than the clinical data, and adding group factors substantially improved clinical fit (CFI=.968, RMSEA=.062; Table S10). The 10-item form showed a similar sample contrast. These findings support a general-severity score while qualifying the unidimensional approximation, particularly in the clinical sample; detailed diagnostics appear in the Supplemental Materials.

*Sample DIF.* Three of 21 items showed DIF between the clinical and weighted general-population samples (Table S15, Panel A). At the same underlying severity, parents in the clinical sample endorsed rage attacks/prolonged tantrums (item 16) more strongly, and hearing voices (item 20) and seeing things (item 21) less strongly. Changes in *R*² were.041,.029, and.034, respectively. Grandiose abilities (item 3), flagged without weights, fell just below the effect-size threshold when weights were applied. Figure 3 orders the items by severity (the mean of the three graded-response thresholds) on a common metric on which the general population has mean 0 and SD 1. The two samples gave nearly the same order (Spearman ρ=.92) because 18 items were constrained equal; the three DIF items moved: hearing voices and seeing things sat much higher in the clinical sample (mean thresholds 3.94 and 4.31 vs. 2.43 and 2.27), and rage attacks lower (1.20 vs. 2.01; Table S22).

**Figure 3.**
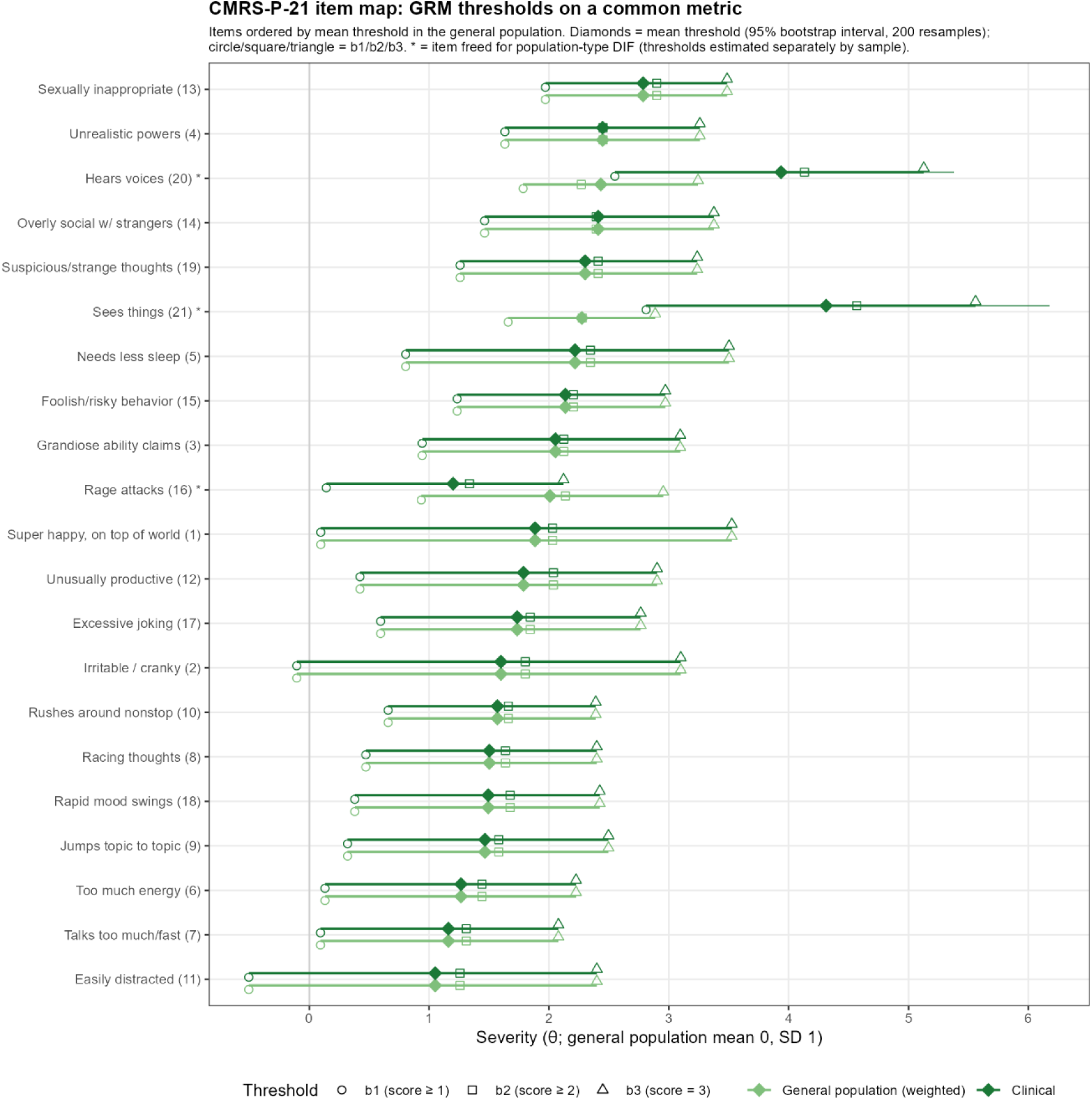
CMRS-P-21 Item Map: Graded Response Thresholds on a Common Metric, Ordered by Mean Threshold ***Note.*** Graded response thresholds (*b1, b2, b3*) and mean threshold (*b̄*, diamond; 95% bootstrap interval) for each CMRS-P-21 item, general-population (weighted) and clinical samples on one linked metric (general population mean 0, SD 1). Items are ordered by *b̄* in the general population, most severe at the top. * = item freed for population-type DIF. Values are in Table S22.

In the unidimensional IRT models, full scalar invariance failed (ΔCFI=−.020); freeing the three DIF items improved fit but left the partial-scalar model just outside the.010 tolerance (ΔCFI=−.012; Table S15, Panel B). Nevertheless, the small total-score effects converged with the multigroup CFA evidence for approximate measurement equivalence. Linking on the 18 items without detected sample DIF placed both samples on a common θ metric, on which the clinical mean was 0.67 SD higher. This comparison depends on the anchor items functioning equivalently across samples.

Figure 4 compares the expected 21-item totals for youths at the same underlying severity: the close curves indicate that sample membership changes the expected total little over most of the trait range. The clinical curve is slightly higher near average severity (maximum difference +0.82 points) but lower at the upper extreme (−2.42 points at θ=3). The mean absolute difference was 0.56 points across θ=−3 to 3 and 0.42 within θ=±2, below the 4.50-point minimal important difference and 14.47-point reliable-change threshold. Thus, most of the approximately seven-point observed mean difference reflected severity rather than differential test functioning.

**Figure 4.**
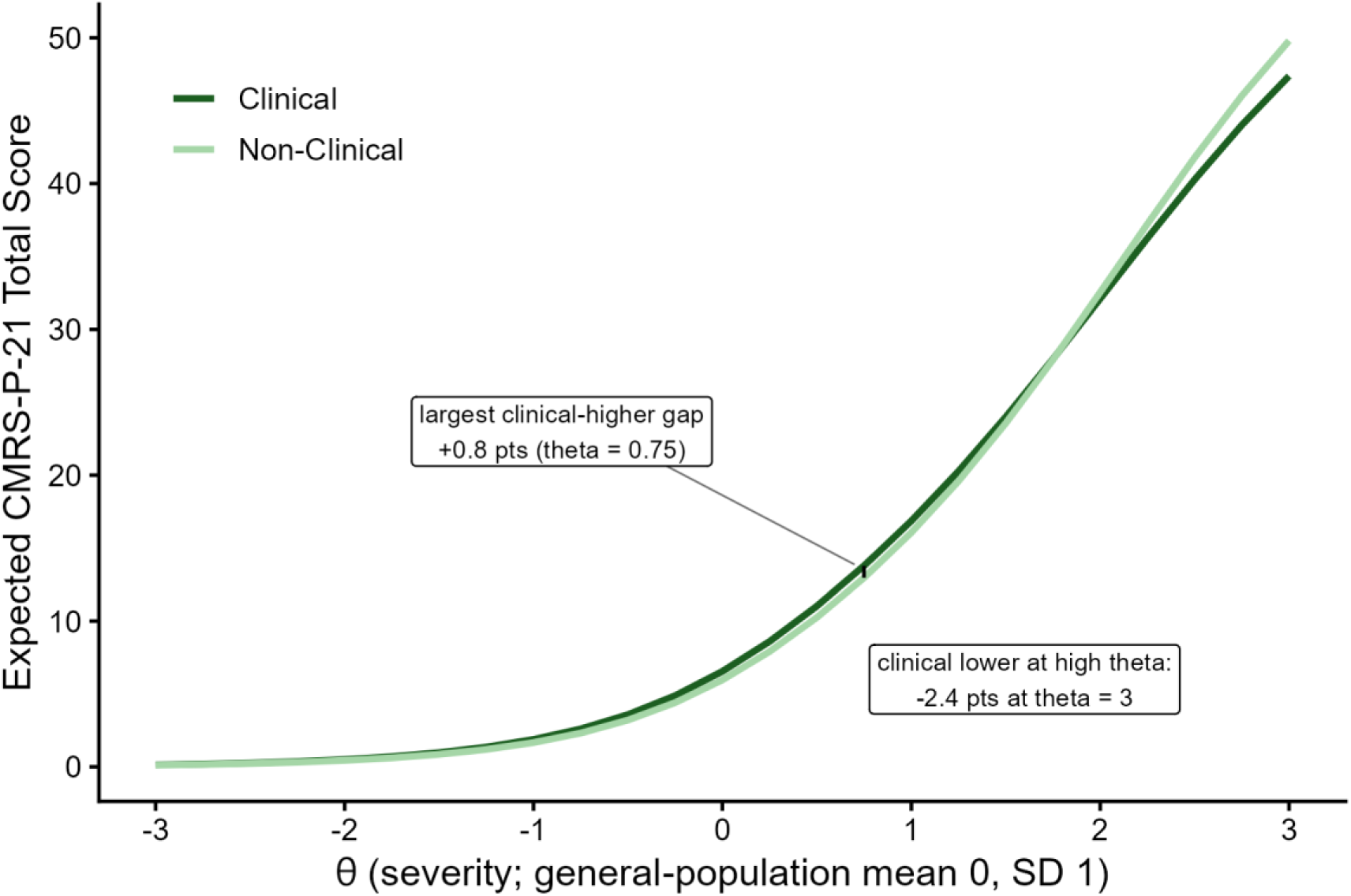
Differential Test Functioning: CMRS-P-21 Total Score by Population Type ***Note.*** Expected CMRS-P-21 total score as a function of latent severity (θ), general population (light green) vs. clinical (dark green), on a common latent metric (general-population mean 0, SD 1) (partial-scalar model: 18 anchor items constrained equal across groups, group means and variances free). The two curves would coincide if the total score meant exactly the same thing in both samples. The visible gap is the differential test functioning (DTF, quantified in Table S15, Panel C).

No items showed DIF by sex, and none were flagged across the three age strata in the weighted analyses; two mood-lability items (items 2 and 18) were flagged for age DIF unweighted but were inconsistently flagged under weighting (33% and 60% of 60 weighted bootstrap resamples; the weighted age screen was unstable overall, with no item flagged in more than 60% of resamples), and multiple-group scalar invariance held by both sex and age stratum (unweighted ΔCFI =.001 and.002).

### Reliability

Omega total provides the best estimate here of overall total-score reliability because it includes reliable variance from both the general and group factors (Table 2). It was.974 for the CMRS-P-21 and.934 for the CMRS-P-10 in the weighted general-population sample. Alpha, conventionally interpreted under a single-common-factor, essentially tau-equivalent model, was.928 and.848, respectively. Omega total therefore better represents the reliable multidimensional variance retained in these summed scores; omega-hierarchical addresses the narrower question of reliability attributable to the general factor alone.

The unidimensional IRT models instead treat the items as indicators of one general severity factor and estimate the precision of the scale’s general-factor score. Empirical reliability was.865 and.768 for the 21-and 10-item forms in the general population and.905 and.809 in the clinical sample (Table S10). Figure 5 (Panels A and B) shows how conditional reliability changes across underlying severity: both forms are most precise in the middle-to-upper range and less precise at the extremes, with the 21-item form maintaining higher reliability over a broader range. Figures 3–5 express θ on the linked clinical and weighted general-population calibration (general-population mean 0, SD 1), with the 18 items without detected sample DIF anchoring a consistent θ metric across samples. Panel C of Figure 5 shows reliability as a function of the raw total score that clinicians observe. The three panels share one θ axis. For the 21-item form, reliability reached.80 at raw scores of 3 in the general population and 2 in the clinical sample and.90 at scores of 7 and 8, and remained above those levels through the top of the observed score range (apart from a slight dip above a score of 53 in the clinical sample). The 10-item form reached.80 at raw scores of 4 and 5 but did not reach.90 at any score. Reliability was lowest at the lowest raw scores (0–2), which are common in the general population.

**Figure 5.**
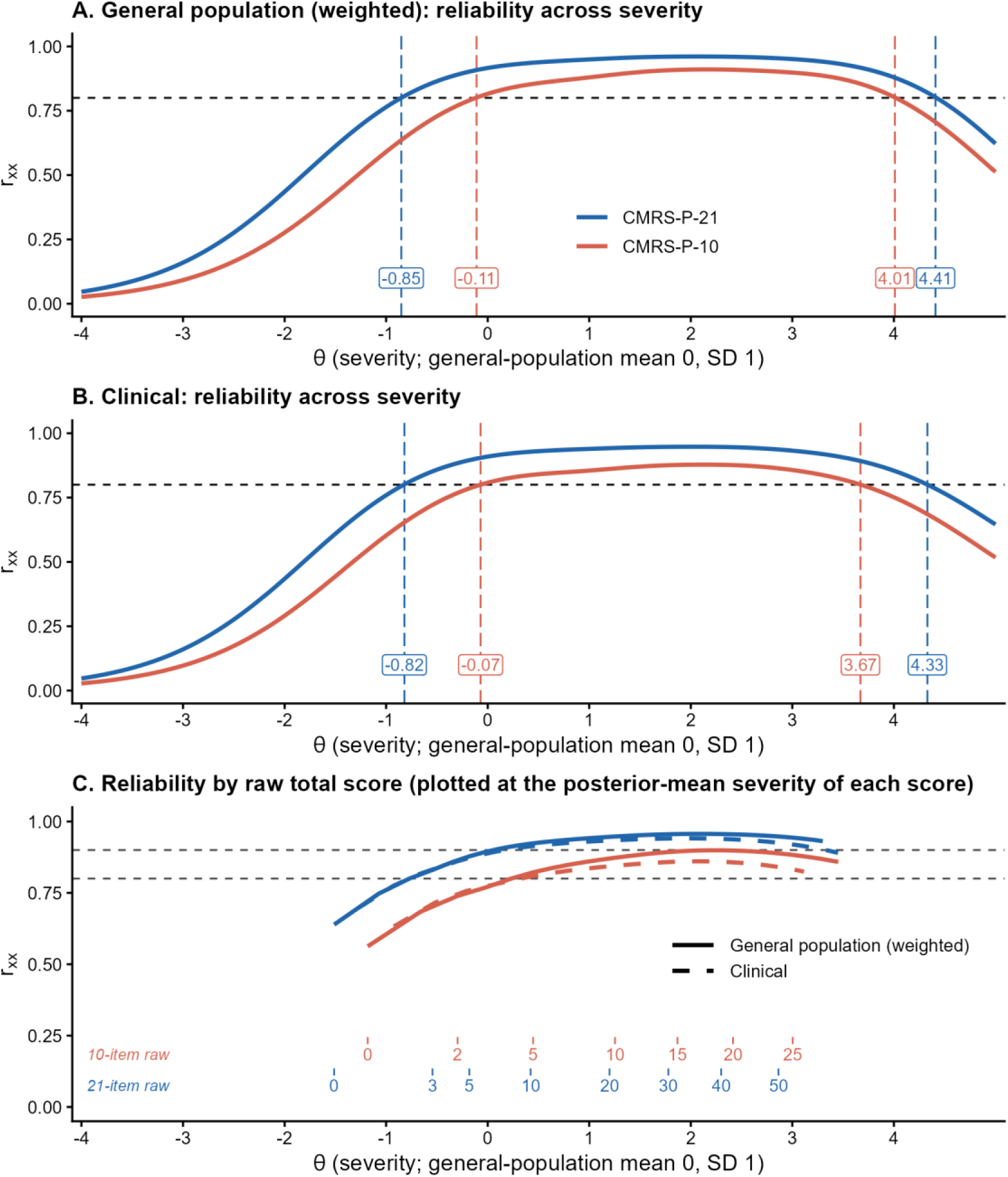
CMRS-P Reliability Across the Severity Range for the 21-and 10-Item Forms: General-Population (Panel A) and Clinical (Panel B) Samples, and Across Raw Total Scores (Panel. **C)** *Note.* Panels A and B: graded response model test-information-based reliability, Iσ²/(Iσ²+1), as a function of latent severity (θ), where I is the test information and σ² is the sample’s latent variance (1.00 in the general population, 0.90 in the clinical sample). The linked calibration uses 18 items without detected population-type DIF as anchors across the weighted general-population and clinical samples, with θ expressed on the metric on which the general population has mean 0 and SD 1 (clinical mean 0.67), as in Figures 3 and 4. Dashed lines mark reliability of.80 and the corresponding severity boundaries for each form. Panel C: reliability of the severity estimate obtained from each raw total score; dashed lines mark reliability of.80 and.90. Solid lines, general population (weighted); dashed lines, clinical sample..

Over 2-3 weeks, 167 parents provided matched retest data (weighted effective *N*=81). Retest correlations were.665 for the CMRS-P-21 and.690 for the CMRS-P-10; absolute-agreement ICCs were.664 (95% CI [.555,.788]) and.679 [.516,.829], respectively (Table 2). Bland–Altman plots showed little average change (baseline minus retest: −0.21 and −0.16 points), but appreciable individual variation, with limits of agreement of −11.55 to 11.13 and −5.90 to 5.58 points (Figure 6). Proportional bias was nonsignificant for the 21-item form (slope=−.110, *p*=.117) and significant for the 10-item form (slope=−.219, *p*=.001), for which retest scores tended to exceed baseline scores more at higher average severity. Stable group means therefore did not imply close agreement for every individual.

**Figure 6.**
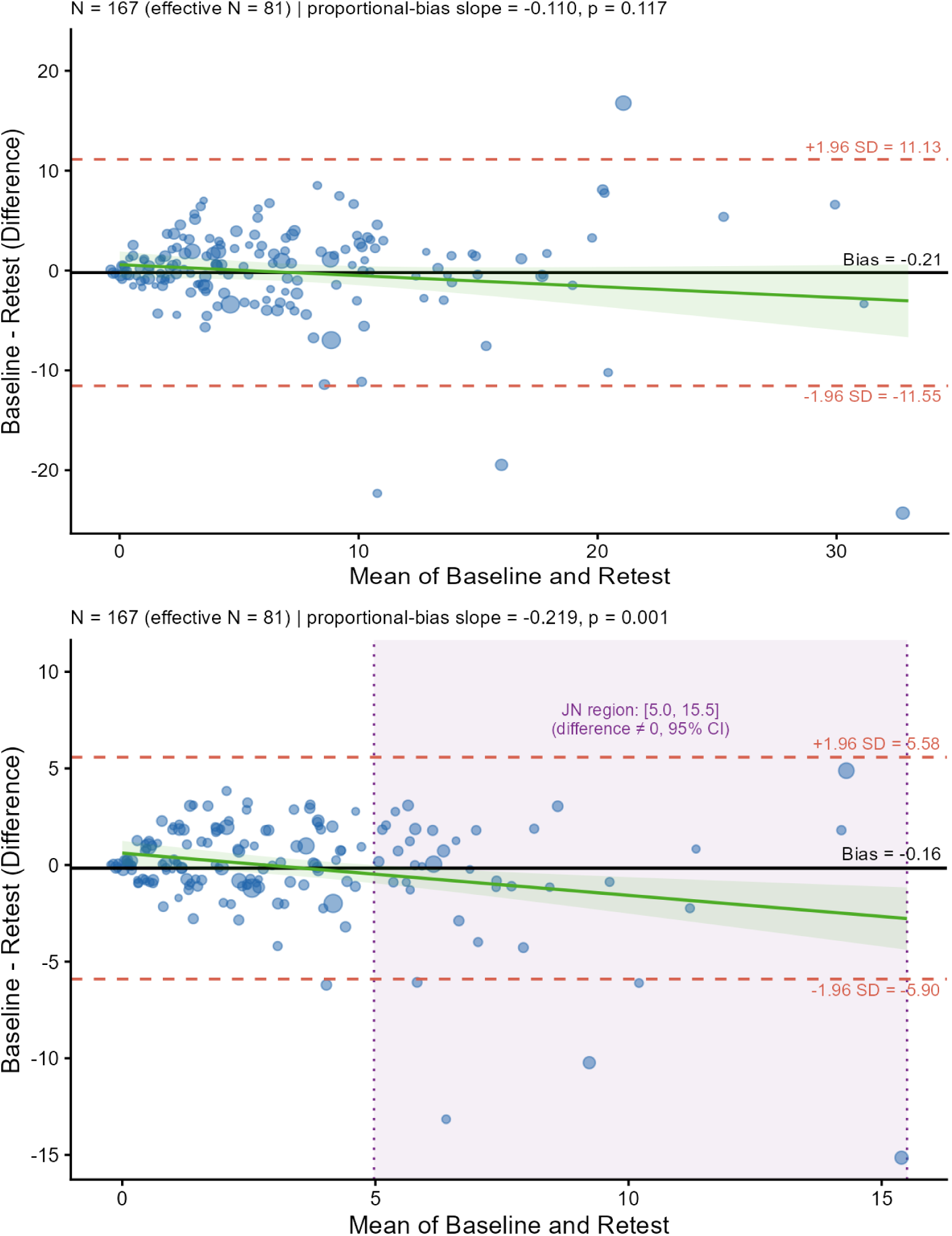
Bland-Altman Plots of Baseline vs. Retest Weighted Total Scores: CMRS-P-21 (top) and CMRS-P-10 (bottom) (weighted analysis) ***Note.*** Matched baseline and retest observations from 167 parents, weighted with baseline weights (effective *N*=81); point sizes reflect weights. Differences are baseline minus retest. Mean differences were −0.21 (21-item) and −0.16 (10-item) points; limits of agreement were −11.55 to 11.13 and −5.90 to 5.58 points, respectively. Proportional-bias slopes were −.110 (*p*=.117) and −.219 (*p*=.001). Shading in the 10-item panel identifies pair means from 5.0 to 15.5 where the fitted difference differs from zero at the 95% level.

Table 2’s standard errors quantify uncertainty in individual scores in raw point units. Omega-based SEMs were 1.44 points for the 21-item form and 1.11 for the 10-item form; the retest-based SEMs were larger, 5.22 and 2.45 points, because they also incorporate variation across occasions. For monitoring change, retest-based SEs are the relevant estimates: after allowing for uncertainty at both assessments, changes exceeding 14.47 and 6.78 points meet the 95% reliable-change criterion (12.14 and 5.69 at 90%). Smaller omega-based thresholds would overstate confidence in longitudinal change. Reliable change also was more conservative than the distribution-based minimal important difference (4.50 and 2.16 points).

### Nomothetic Clinical Significance Benchmarks

Clinical-significance benchmarks were 26.53 (CMRS-P-21) and 12.68 (CMRS-P-10) by Method B and 11.73 and 5.18 by Method C (Table 2). Method A was not reportable because the clinical mean minus 2 *SD* fell below the scale floor. Method B was close to the weighted empirical 95th percentile in the general population (28 and approximately 13, respectively). Table 2 reports the thresholds, and Figure 1 shows them superimposed over the violin plots for the clinical and weighted general population samples.

### Validity

Because this is the first evaluation of the CMRS-P in a normative sample, we examined associations with measures of co-occurring or phenotypically similar constructs (Table S16; Figure 7). The CMRS-P-21 showed large positive correlations with attention and hyperactivity problems, suicidality, depression, externalizing symptoms, and emotion dysregulation (*r*=.51–.63). Associations with sleep-related impairment (*r*=.49), global emotional severity (*r*=.48), and broad internalizing symptoms (*r*=.45) were also substantial and close to Cohen’s descriptive signpost for a large correlation. Associations were more moderate with poorer quality of life (*r*=.30, with the direction reversed for presentation) and family mood-disorder history (*r*=.28), and smaller with academic and social performance problems (*r*=.21–.26). Correlations with pubertal development were negligible (*r*=.01,.00, and.03 for total, boys’, and girls’ PDS scores, respectively), supporting discrimination from developmental maturation.

**Figure 7.**
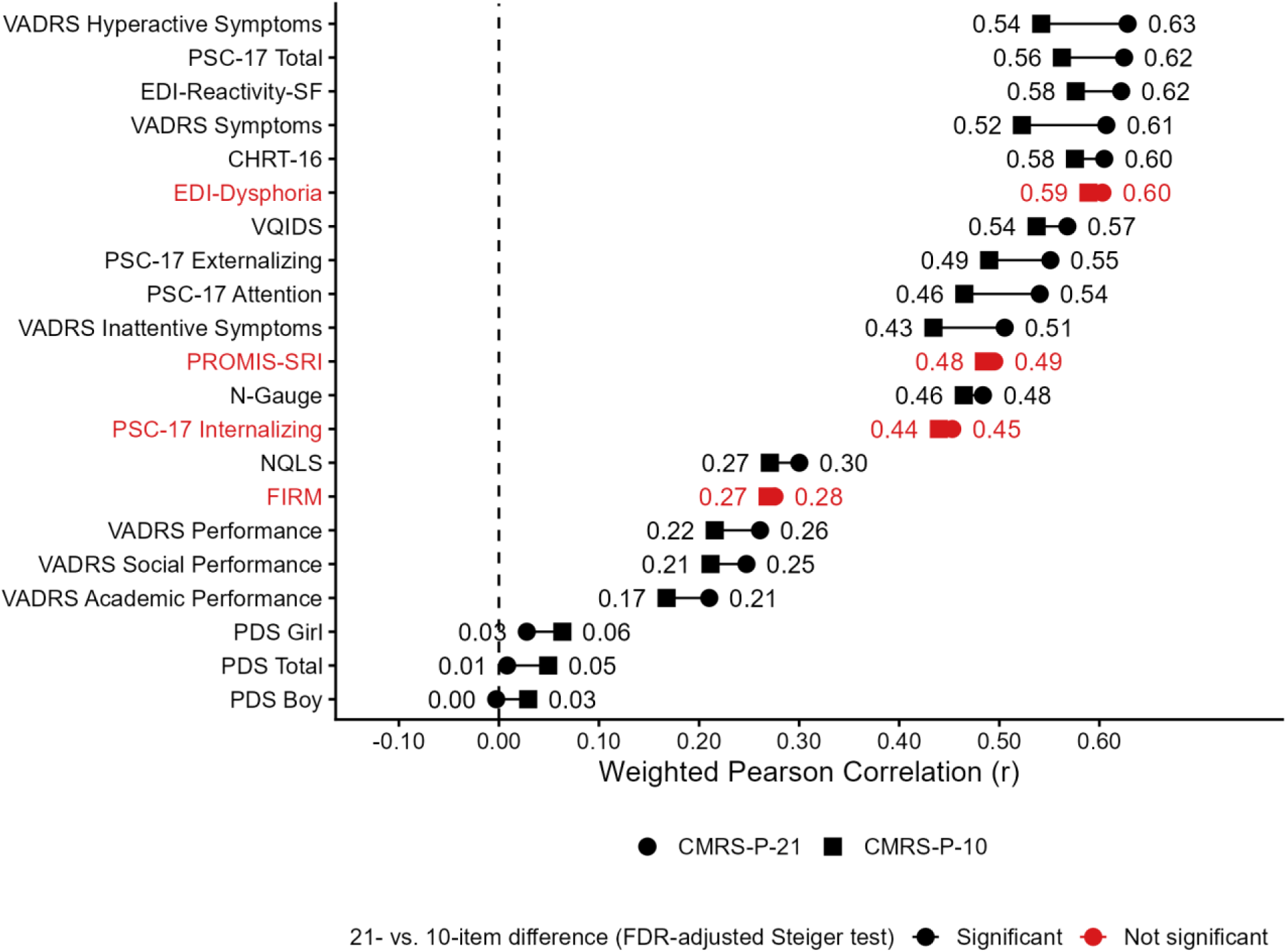
CMRS-P Cross-Scale Pearson Correlations (r) with Parent-Reported Criteria (weighted sample) ***Note.*** NQLS is reverse-keyed (higher = better quality of life) and its sign is flipped here, as in Table S16, so that all correlations point toward greater problems. Red indicates difference not statistically significant (Steiger’s *Z* for dependent, overlapping correlations; Benjamini–Hochberg FDR-adjusted *p* ≥.05). Listwise deletion (*N*=2,169; PDS Girl *n*=1,029, PDS Boy *n*=1,140), as in Table S16.

The CMRS-P-10 showed a closely similar pattern, with generally slightly weaker correlations. Its associations with PSC-17 Attention and Externalizing scores (*r*=.46 and.49) remained substantial and near the large-effect signpost; they did not represent a qualitatively different level of convergence from the corresponding full-form correlations. Likewise, the quality-of-life association (*r*=.27) was comparable in magnitude to that of the full form (*r*=.30). Associations with sleep-related impairment, global emotional severity, and internalizing symptoms were also similar across forms (*r*=.44–.48 for the brief form), whereas pubertal-development correlations remained negligible (*r*=.03–.06). Although many between-form differences were statistically significant, their magnitudes were small (absolute Cohen’s *q*≤.13), with the largest differences involving attention, hyperactivity, and externalizing symptoms. Overall, shortening the scale largely preserved its pattern of associations with related constructs. The pattern of correlations was largely consistent across age groups. After correction for multiple comparisons, the CMRS-P-21 showed stronger associations with emotion dysregulation in adolescents than in children. Both forms also showed age differences in their associations with pubertal development, although correlations were small and of limited practical significance. Supplemental materials include detailed age-group comparisons under Validity.

Differences between scoring methods (unit-weighted versus IRT-based EAP scores) are shown in Tables S17a, S17b, and S17c, and brief-form psychometrics for the CMRS-P-10 are in Table S18. Overall, simple sum scores showed comparable or slightly stronger associations with external criteria than IRT-based scores, while the brief form retained an essentially unidimensional structure and closely tracked the full-scale total (*r*=.95).

### Continuous Norms

Based on the above analyses, norming concentrated on the raw total score for both the 21-and 10-item versions. Cross-validation favored a linear age-by-sex mean structure for both forms (Table S20). The age pattern of the CMRS-P-21 percentiles differed by sex (Table 4, with *T*-score equivalents; Figure 8 and Figures S2a and S2b). Male percentiles declined with age, and this was robust across sensitivity analyses (see Supplemental materials). Female percentiles were flat for the median or lower, but increased significantly with age for the higher percentiles in the weighted analyses; this was less robust in the sensitivity analyses. CMRS-P-10 percentiles showed the same sex-specific pattern.

**Figure 8.**
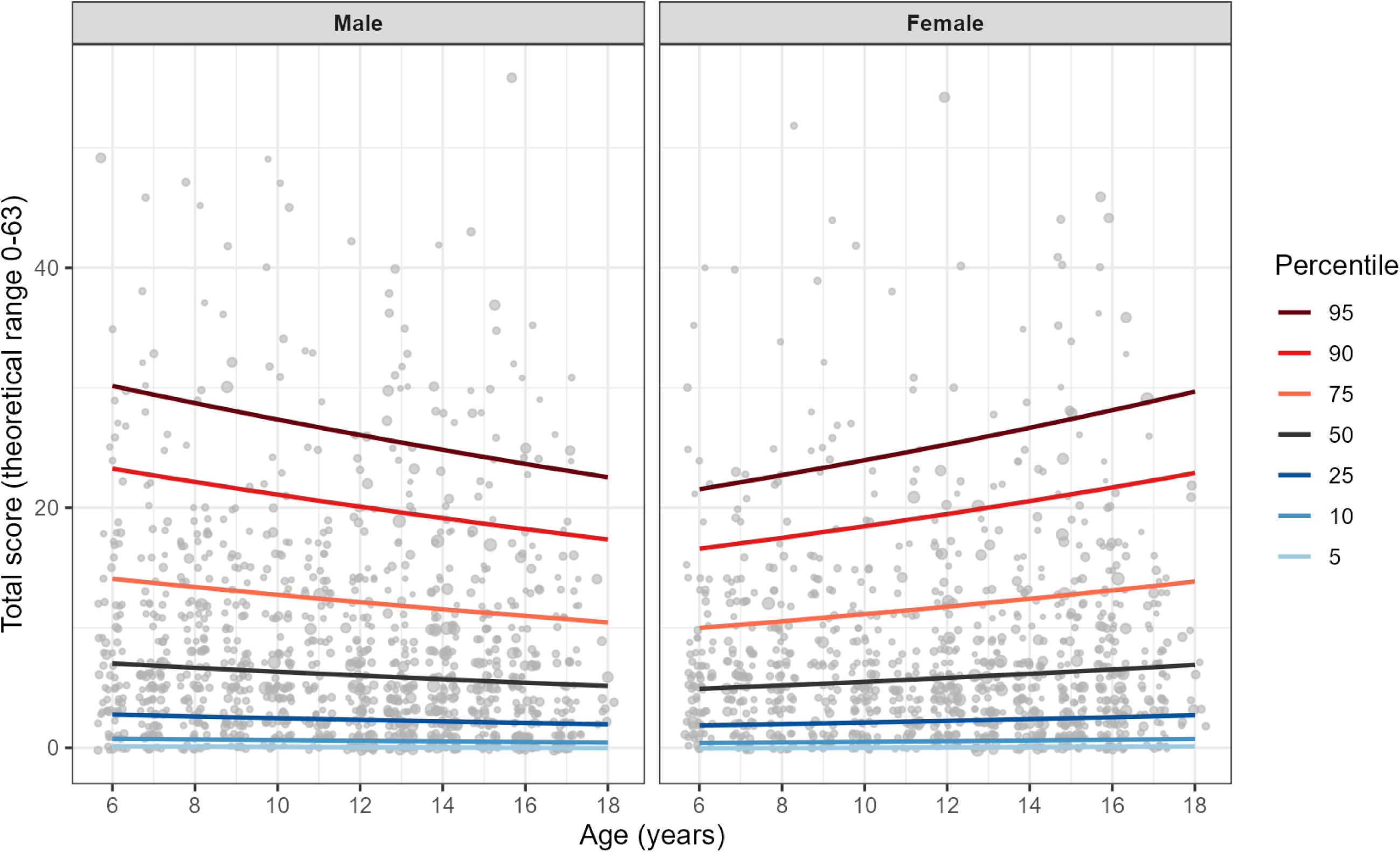
CMRS-P-21 Age-Based Percentile Growth Curves (survey-weighted Negative Binomial type I distribution [NBI], linear age x sex)

**Table 4.** CMRS-P Total Score Percentiles and T-Score Equivalents by Age and Sex (General-Population Norms, Complete Cases, N=2,229; Survey-Weighted Negative Binomial Model with Linear Age, Sex, and Age × Sex)

| Age | Male |  |  |  |  |  |  | Female |  |  |  |  |  |  |
| --- | --- | --- | --- | --- | --- | --- | --- | --- | --- | --- | --- | --- | --- | --- |
|  | P95 | P90 | P75 | P50 | P25 | P10 | P5 | P95 | P90 | P75 | P50 | P25 | P10 | P5 |
|  | T 66 | 63 | 57 | 50 | 43 | 37 | 34 | T 66 | 63 | 57 | 50 | 43 | 37 | 34 |
| **CMRS-P-21 (Range 0–63)** |  |  |  |  |  |  |  |  |  |  |  |  |  |  |
| 6 | 30 | 23 | 14 | 7 | 3 | 1 | 0 | 22 | 17 | 10 | 5 | 2 | 0 | 0 |
| 7 | 29 | 23 | 14 | 7 | 3 | 1 | 0 | 22 | 17 | 10 | 5 | 2 | 0 | 0 |
| 8 | 29 | 22 | 13 | 7 | 3 | 1 | 0 | 23 | 17 | 11 | 5 | 2 | 0 | 0 |
| 9 | 28 | 22 | 13 | 6 | 3 | 1 | 0 | 23 | 18 | 11 | 5 | 2 | 0 | 0 |
| 10 | 27 | 21 | 13 | 6 | 2 | 1 | 0 | 24 | 18 | 11 | 5 | 2 | 0 | 0 |
| 11 | 27 | 21 | 12 | 6 | 2 | 1 | 0 | 25 | 19 | 11 | 6 | 2 | 1 | 0 |
| 12 | 26 | 20 | 12 | 6 | 2 | 1 | 0 | 25 | 19 | 12 | 6 | 2 | 1 | 0 |
| 13 | 25 | 20 | 12 | 6 | 2 | 1 | 0 | 26 | 20 | 12 | 6 | 2 | 1 | 0 |
| 14 | 25 | 19 | 12 | 6 | 2 | 1 | 0 | 27 | 21 | 12 | 6 | 2 | 1 | 0 |
| 15 | 24 | 19 | 11 | 6 | 2 | 1 | 0 | 27 | 21 | 13 | 6 | 2 | 1 | 0 |
| 16 | 24 | 18 | 11 | 5 | 2 | 0 | 0 | 28 | 22 | 13 | 7 | 3 | 1 | 0 |
| 17 | 23 | 18 | 11 | 5 | 2 | 0 | 0 | 29 | 22 | 13 | 7 | 3 | 1 | 0 |
| 18 | 23 | 17 | 10 | 5 | 2 | 0 | 0 | 30 | 23 | 14 | 7 | 3 | 1 | 0 |
| **CMRS-P-10 (Range 0–30)** |  |  |  |  |  |  |  |  |  |  |  |  |  |  |
| 6 | 13 | 10 | 6 | 3 | 1 | 0 | 0 | 10 | 7 | 4 | 2 | 1 | 0 | 0 |
| 7 | 13 | 10 | 6 | 3 | 1 | 0 | 0 | 10 | 8 | 5 | 2 | 1 | 0 | 0 |
| 8 | 13 | 10 | 6 | 3 | 1 | 0 | 0 | 11 | 8 | 5 | 2 | 1 | 0 | 0 |
| 9 | 13 | 10 | 6 | 3 | 1 | 0 | 0 | 11 | 8 | 5 | 2 | 1 | 0 | 0 |
| 10 | 13 | 10 | 6 | 3 | 1 | 0 | 0 | 11 | 9 | 5 | 3 | 1 | 0 | 0 |
| 11 | 12 | 10 | 6 | 3 | 1 | 0 | 0 | 12 | 9 | 6 | 3 | 1 | 0 | 0 |
| 12 | 12 | 9 | 6 | 3 | 1 | 0 | 0 | 12 | 10 | 6 | 3 | 1 | 0 | 0 |
| 13 | 12 | 9 | 6 | 3 | 1 | 0 | 0 | 13 | 10 | 6 | 3 | 1 | 0 | 0 |
| 14 | 12 | 9 | 5 | 3 | 1 | 0 | 0 | 14 | 10 | 6 | 3 | 1 | 0 | 0 |
| 15 | 12 | 9 | 5 | 3 | 1 | 0 | 0 | 14 | 11 | 7 | 3 | 1 | 0 | 0 |
| 16 | 11 | 9 | 5 | 3 | 1 | 0 | 0 | 15 | 11 | 7 | 3 | 1 | 0 | 0 |
| 17 | 11 | 9 | 5 | 3 | 1 | 0 | 0 | 15 | 12 | 7 | 4 | 1 | 0 | 0 |
| 18 | 11 | 9 | 5 | 2 | 1 | 0 | 0 | 16 | 12 | 8 | 4 | 1 | 0 | 0 |

*Note.* P denotes percentile. The row below the percentile headings gives corresponding *T* scores. Percentiles are the smallest integer scores at which the fitted survey-weighted negative binomial cumulative distribution reaches the specified probability. Both forms use linear age, sex, and their interaction, with constant dispersion. Bootstrap intervals and effective sample sizes are in Table S21; estimates at ages 17–18 are provisional. Model specifications and sensitivity analyses are described under Norms in the Supplemental Materials.

## Discussion

The goal of the present study was to create continuous age-and sex-based norms for the parent-report Child Mania Rating Scale. We gathered a nationally representative sample of 2,229 caregivers of youths aged 6–18 years old, and 167 (7.5%) of them completed the scale again 2-3 weeks later to evaluate change over time in the absence of treatment. Because mania is a clinical construct that is rare in the general population of youths, yet can look similar to other more common issues, we used rigorous statistical methods to evaluate whether the CMRS-P measures one thing versus a set of related problems (“essential unidimensionality”), supporting the use of the total score as the main basis for interpretation. We used a large sample of 522 caregivers seeking outpatient mental health services for their child to establish how the CMRS-P works in a clinical context, creating a foil to test the stability of the measurement properties of the scale when shifting between the general population and a clinical setting. We evaluated the 10-item short version as well as the 21-item full length scale. Our goal at each analytic step was to provide something clinically useful (norms, a short form, benchmarks for evaluating clinically significant change over brief intervals), putting it on the strongest statistical footing feasible, and testing some of the potential limits where application might need additional caution.

### Are All the Symptoms on the CMRS Measuring One Thing?

Users can feel confident working with the total score. A general severity factor carried nearly all reliable variance; no group factor cleared distinctiveness thresholds in the general population sample. Even though a four-factor structure showed metric invariance in both the clinical and the general population samples, the reliable variance distinct to the specific group factors was much larger in the clinical sample. All methods (CFA, omega decomposition, IRT) converged on a strong general factor underlying both the 21-and 10-item sets (“essential unidimensionality”), strongly supporting clinical interpretation of a total score. Conversely, the specific group factors did not have enough reliable unique variance to warrant investigation in the general population sample. This is consistent with prior studies emphasizing the total score (Pavuluri 2006; Henry 2008; Meyer 2024).

### How Well Does the CMRS-P Measure Manic Symptoms? Reliability and Scoring

We evaluated reliability of both 21-and 10-item versions multiple ways. Internal consistency was excellent for both forms. Omega_Total_ is the most appropriate summary of internal consistency (instead of the more common alpha) because of the underlying factor complexity. Both indicate good internal consistency reliability for both lengths. Analyses found no advantage to using IRT scoring instead of unit-weighting, using a simple sum of the items. Clinicians can simply add up the items at no loss of validity compared to more complex scoring (Wainer, 1976).

Retest stability was evaluated over approximately two weeks, an interval relevant to measurement-based care. Because participants were recruited from the general population rather than a treatment study, these estimates provide a practical benchmark for score fluctuations outside a study-directed intervention, reflecting both measurement error and short-term changes in mood. Retest dependability was lower than internal consistency (ICC[A,1]=.66 and.68 for the CMRS-P-21 and CMRS-P-10, respectively). Average scores changed negligibly, decreasing by 0.21 and 0.16 points, but the Bland–Altman plots showed wide limits of agreement. Thus, stable group means masked substantial individual variation, underscoring the need to distinguish reliable change from ordinary fluctuation when interpreting repeated assessments. Clinicians now have a yardstick for mood changes that are typical of childhood and adolescence.

### Built in the Clinic…Does it Work Similarly in the General Population?

Tests of measurement invariance showed strongly configural, metric, and mostly scalar invariance between the clinical and general population samples, and supplemental analyses also found no evidence of differential item functioning (DIF) between males and females, and only weak instances across age strata. The differences in item thresholds that were observed in three items did not amount to substantively meaningful differences in the performance of the total score. On a common latent metric, however, the flagged items shift the expected total score by less than a point on average (about +0.8 points near average severity and −2.9 points at the extreme upper end). Most of the 7.0-point observed difference between the samples reflects a real difference in severity (about *0.67 SD*).

### Does the CMRS Score Measure Mood Symptoms? Criterion, Convergent, and Discriminant Validity

Scores correlated strongly with attention problems, suicidality, depression, externalizing symptoms, and emotion dysregulation, but negligibly with pubertal status. This broad convergence underscores that elevated CMRS-P scores signal psychopathology without establishing specificity to mania. The pattern and magnitude of associations with externalizing, internalizing, and attention problems resemble those reported for other parent-rated mania scales in clinical samples (E. A. Youngstrom et al., 2020). Associations with sleep problems and the negligible relationship with pubertal development extend that evidence. Establishing diagnostic specificity will require testing classification accuracy against structured diagnostic interviews (Rettew et al., 2009) or longitudinal expert evaluation of all data (LEAD) diagnoses (Spitzer, 1983). Meta-analyses including studies that have used these diagnostic definitions have found that the CMRS-P is in the top tier of classification accuracy among all scales with published results (Alcaino et al., 2024; E. A. Youngstrom et al., 2015).

### Clinical Benchmarks for Measurement-Based Care and Clinical Signifance

Different clinical benchmarks address distinct questions: whether change may be noticeable, whether it exceeds measurement error and short-term fluctuation, and whether a score is more characteristic of a clinical or general-population distribution. The distribution-based minimal important differences (MIDs; 4.5 points for the CMRS-P-21 and 2.2 for the CMRS-P-10) are benchmarks for potentially noticeable change (Norman et al., 2003), although this interpretation ideally would be confirmed with patient-or caregiver-reported improvement. Jacobson–Truax Methods A, B, and C place scores relative to the clinical distribution, the general-population distribution, and the boundary between them, respectively. Methods A and C depend on the clinical reference sample and would shift with its severity and diagnostic composition; here, Method A fell below the scale floor and was not interpretable.

For evaluating individual change, the dependability-based reliable change indices are conceptually appropriate benchmarks: approximately 14.5 points on the CMRS-P-21 and 6.8 on the CMRS-P-10 at 95% confidence, or 12.1 and 5.7 points at 90% confidence. The corresponding unweighted 95% thresholds were 11.5 and 6.2 points. Omega-based thresholds were substantially smaller (4.0 and 3.1 points) because they capture internal-consistency error but omit variation across occasions; they make less sense for identifying reliable longitudinal change. However, retest dependability is rarely reported for scales, so it is commonplace to use internal-consistency based reliable change benchmarks (Ogles et al., 2001). Retest dependability of.66 and.68 therefore limits detection of small changes within individuals: a change may be noticeable yet remain below the threshold for distinguishing it confidently from measurement error and short-term fluctuation. These findings favor using the scales to identify clinically meaningful elevations and evaluate group-level change, with greater caution when interpreting modest changes in an individual’s score. We note that the less conservative benchmarks often still are helpful as interim milestones to keep families engaged with treatment.

### Norms: (Finally) Seeing the Range of Developmentally Typical Moodiness

Weighted norms were derived from a parsimonious negative binomial model incorporating linear age, sex, and their interaction. Cross-validation favored this specification over a flexible smoother, whose irregular mid-adolescent rise did not improve out-of-sample prediction. Upper percentiles declined with age in males and increased in females, yielding higher reference scores for males at younger ages and females in adolescence. Although sex differences have previously been reported in a community sample of 183 children (Verstraeten et al., 2012), the age-dependent pattern observed here requires replication: the female increase did not persist in unweighted or covariate-standardized analyses, and estimates at the oldest ages rested on few effective respondents. Norms for ages 17–18 should therefore be treated as provisional, and model selection is undergoing further statistical review.

### Clinical Implications

Clinical interpretation should emphasize the total score, as the findings do not support interpreting subscale profiles at the individual level. Either form is defensible for screening, given their strong correspondence (*r*=.95) and precedent for community administration of the brief form (Taylor et al., 2022). Age-referenced norms can contextualize how unusual a score is and inform treatment goals, using the appropriate reference population because a score’s relative standing differs across settings. The norms assume complete responses; prorating a small number of missing items may provide an approximation, but was not validated here and should not be extended to substantial missingness. Elevated scores should prompt deeper assessment rather than be treated as evidence of mania.

### Generalizability

The weighted normative sample was demographically representative of U.S. youths aged 6–18, supporting generalizability within that population across age, sex, and region. The CMRS has also been translated and evaluated in convenience samples in Brazil, Spain, and Sweden, with broadly consistent psychometric findings (Meyer et al., 2024; Serrano et al., 2011, 2013; Taylor et al., 2022). To our knowledge, this is the first CMRS study designed to represent national demographics. However, demographic representativeness within one country does not establish the applicability of its norms elsewhere. International ASEBA research has identified societal differences in average scores despite relatively stable measurement structures (Rescorla et al., 2012). Because norms locate an individual’s score within a reference distribution, applying these U.S. norms in other societies requires caution and further evaluation of both measurement equivalence and local score distributions.

### Limitations

Several limitations constrain interpretation. Parents completed the CMRS-P without assistance and may not have consistently applied its instruction to endorse behaviors only when troublesome or developmentally unusual (Pavuluri et al., 2006); some responses may therefore capture occurrence rather than impairment. All criterion measures shared the same informant and online format, likely inflating convergence relative to independent assessments. Psychosis-spectrum items also showed pronounced floor effects, limiting estimation of their properties in the general population. Comparisons with the regional, unweighted Ohio outpatient sample confound clinical status with geography and recruitment, and adding the CMRS-P after enrollment began restricted analyses to a subset of clinical participants. Diagnostic accuracy was not evaluated here, and the single clinical assessment precluded evaluating stability in that sample.

Weighting improved demographic alignment but reduced precision: the effective sample was approximately 1,156 respondents, with standard errors roughly 1.4 times larger than under simple random sampling. Broad conclusions about reliability, dimensionality, and criterion validity survived unweighted analyses, but some invariance, DIF, local-dependence, and age-moderation findings changed. Retest estimates were particularly sensitive, resting on only about 81 effective pairs with weights developed for the full sample. The norms warrant additional caution at the distributional tails and age extremes, where effective samples were small and percentile intervals wide (Table S21). Cross-validation favored a parsimonious linear age-by-sex model, but residual diagnostics indicated missed age patterns, and the female upper-percentile increase did not persist in unweighted or covariate-standardized analyses. The latter sensitivity analysis also omitted some variables used to construct the weights. Because the age-function selection rule was specified after inspecting the CMRS-P results; the current age-and sex-specific curves should be treated as provisional.

### Future Directions

Future research should use cognitive interviewing to determine whether parents distinguish developmentally unusual or impairing symptoms from normative behaviors, and refine item wording where that distinction breaks down. Evaluating youth self-report could broaden assessment, whereas teacher-report adaptations would need particular attention to distinguishing manic symptoms from ADHD. Classification accuracy should be tested against rigorous diagnostic assessments; until then, the 95th percentile is best treated as a prompt for further evaluation rather than a diagnostic threshold. Studies across clinical settings of differing acuity should also test whether item information would support different brief forms within a common measurement framework across a system of care.

### Clinical Take Home Messages

The CMRS-P is a reliable and valid measure of manic symptoms in the general population. Interpretation should focus on the total raw score. In the general US population, there are age and sex differences in typical mood and energy that now have norms available. Elevated scores should be treated as a flag for further evaluation, not “testing positive for bipolar disorder.” The CMRS-P does not show evidence of meaningful sore bias between clinical outpatient and general population samples, nor sex or age bias. A range of clinical significance benchmarks are now added to the measurement-based care toolkit for working with mood symptoms in youths, including conservative definitions of reliable change compared to everyday shifts in mood for youths.

## Data Availability

All data produced in the present study are available upon reasonable request to the authors.

