## Supplemental Materials for "Parent-Reported Child Mania Rating Scale: Reliability, Validity, and Norms in a Nationally Representative US Sample"

Supplemental Material

### Reliability, Preliminary Validity, and Continuous Norms for the Child Mania Rating Scale (CMRS-P) in a Nationally Representative Sample of 6–18-Year-Olds: Results from the BENCHMARK Project

#### Methods

The main manuscript presents a condensed Methods section. The corresponding subsections below provide expanded details on recruitment, measures, statistical analyses, and continuous norming. Supporting tables and figures follow the supplemental Results.

*Participants*

Eligibility required being a parent or guardian of a youth in the target age range. Parents supplied demographic information and, when more than one child was eligible, selected the child whose birthday was closest to a randomly generated date. Quality controls included attention checks and exclusion of the fastest respondents and those with the most missing responses. YouGov matched respondents to a sampling frame derived from the 2023 American Community Survey and applied propensity-score and post-stratification weights (YouGov, 2026).

The final sample for the present study included 993 parents ($M_{age}$=40.0 years, *SD*=6.7; 60.6% female) of a child 6 to 11 years old ($M_{age}$=8.4 years, *SD*=1.7; 46.1% female) and 1,236 parents ($M_{age}$=46.9 years, *SD*=8.5; 68.2% female) of a child 12 to 18 years old ($M_{age}$=14.4 years, *SD*=1.7; 48.5% female) representing all 50 states of the United States of America and the District of Columbia. The census regions of study participants closely resembled that of the US population, with slight overrepresentation of participants located in the Northeast and Midwest and slight underrepresentation of participants who resided in the South and Midwest [CITE ACS

2023 FROM CENSUS.GOV] (United States Census Bureau, 2023). main-text Table 1 provides key demographic data.

*Procedure*

Participants were recruited from a large panel curated by an international online survey company, YouGov. They completed a preregistered battery designed to develop nationally representative norms and evaluate psychometrics for 15 parent-report and youth self-report mental, behavioral, and physical health assessments, including the Child Mania Rating Scale (E. Youngstrom et al., 2025). The Nationwide Children's Hospital's Behavioral Evaluation and Norms for Child Health and MARKers of Well-being (BENCHMARK) 2025 Project—is a cross-sectional study with three strata: 1,000 parents of a child aged 6–11 years, 1,252 parents of a child aged 12–18 years, and 1,008 children aged 12–18 years, completing matching dyads with their parent. Of the parents of 12–18-year-olds, 246 did not have a child who participated in the study.

Data quality was ensured through attention check questions and qualitative and quantitative cleaning procedures, including the removal of the top 2% of "speeders" and "skippers" (respondents who completed the survey faster than and/or skipped more items than 98% of the sample). Following data cleaning, remaining survey panelists were then matched to a sampling frame on gender, age, race, and education constructed by stratified sampling from the full 2023 American Community Survey (ACS) 1-year sample. The matched cases were weighted to the sampling frame using propensity scores. Finally, the weights were then post-stratified on a four-way stratification of gender, age (4-categories), race (4-categories), and education (4-categories), to produce the final weight (YouGov, 2026). The weights apply to the general-population sample only; the clinical comparison sample is unweighted. All results for the general-population sample reported in this paper, including main-text Tables 1 and 2, apply these weights (percentages and means are weighted; counts are unweighted *n*; see Statistical Analyses for the weighted methods and the design effect).

All study procedures were reviewed and designated Not Human Subjects Research (NHSR) by the Institutional Review Board at Nationwide Children's Hospital because there was no direct contact, nor were individual identifiers included in the data frame. Data collection for the current study sample occurred between July 11, 2025 and March 7, 2026.

*Measures*

***Demographics and Eligibility***

Prior to gaining access to the main survey, parent participants provided their own birth year, race, ethnicity, education level, ZIP code, and the age, and sex assigned at birth of all children in their care. Respondents with more than one child aged 6–11 or 12–18 were instructed to complete the survey about their child whose birthday (month/day) fell closest to a randomly generated date (month/day). Respondents who were not a parent or guardian of a child aged 6–11 or 12–18 were redirected out of the survey to the YouGov website homepage.

***Clinical Comparison Sample***

The CMRS-P-21 was added to the protocol for an NIMH funded project (R01 MH066647) (E. Youngstrom et al., 2005) in the third year of data collection as an IRB-approved modification. Families enrolled before the modification were not given the scale. The clinical dataset includes 828 families; the 522 parents with complete data on all 21-items are analyzed here (the others either enrolled before the CMRS was added or had incomplete CMRS data). They were parents of youths aged 4 to 18 years seeking outpatient mental health services in Ohio. The parent and youth completed a semi-structured diagnostic interview (KSADS-PL Plus;

Geller et al., 2001; Kaufman et al., 1997) with highly trained raters, using a longitudinal expert evaluation of all data (LEAD) consensus process (Spitzer, 1983) to establish diagnoses for the youth. These data provide a clinical comparison sample to evaluate the factor structure and item response characteristics in a multigroup framework, including potential differential item functioning as the severity of the underlying trait increases. The research protocol did not include any of the other scales in the BENCHMARK battery, precluding the possibility of direct comparisons of criterion correlations. The clinical sample is regional and unweighted, whereas the general-population sample is national and weighted to national margins.

***Assessment Battery***

The BENCHMARK 2025 assessment battery consisted of a total of 15 free scales assessing internalizing, externalizing, and attention problems; sleep; pubertal status; quality of life; academic performance; and overall functioning in children and adolescents (E. Youngstrom et al., 2025). The measures administered to each informant depended on the sample group—parents of 6–11-year-olds completed 14 measures, parents of 12–18-year-olds completed all 15 measures, and youth aged 12–18 completed 13 measures. While many assessments already had multi-informant versions, the exceptions were adapted for either parent-report or youth self-report so that matching scales were available for both informants. In these cases, instructions and item wording were adjusted to match the respondent, with changes made only for grammatical clarity and ease of comprehension; the underlying item content was unchanged. Additional details about the full assessment battery broken down by respondent can be found in the Supplemental Materials and preregistration, while those used in analyses for the current study are described below.

**Child Mania Rating Scale, Parent-Report (CMRS-P)**: The CMRS-P is a 21-item, parent-rated scale used to assess the presence of *DSM-IV*-identified manic symptoms, distinguish between clinical presentations of bipolar disorder (BD) and other psychiatric conditions, and monitor treatment and symptom progress (American Psychiatric Association, 1994; Pavuluri et al., 2006). Among the most used mania screening instruments in clinical settings, the CMRS-P has been translated and validated in multiple languages and has a 10-item short form (Chi et al., 2023; Henry et al., 2008; Meyer et al., 2024; Serrano et al., 2011). Items are answered on a 4-point Likert-type scale (0: Never/Rare, 1: Sometimes, 2: Often, 3: Very Often) with respondents instructed to base answers on the youth's mood and behavior over the past month. Higher scores indicate greater manic symptom severity. The original study reported internal consistency (Cronbach's α) of .96 (Pavuluri et al., 2006). Henry and colleagues used Rasch modeling to select 10-items to make a short form of the CMRS-P. We moved the 10-items included on the short form to the front of the CMRS used in the BENCHMARK protocol so that the presentation order and context effects would better generalize to applications only giving the 10-item version, while also gathering the additional items to be able to evaluate the 21-item version as well. The clinical sample used the standard order for the 21-items, so analyses of the 10-item form in that sample are using embedded items. Prior research comparing embedded and extracted administration formats on another mania scale in the clinical sample found that changes in item response characteristics were negligible (Freeman et al., 2012).

##### *BENCHMARK Convergent and Discriminant Validity Indicators*

**Concise Health Risk Tracking (CHRT-16):** The CHRT-16 is a 16-item screening tool for assessing suicidal ideation and risk for suicide based on feelings or actions over the past week. Items are rated on a 5-point Likert scale from 0 (Strongly Disagree) to 4 (Strongly Agree).

As there is not a parent-informant CHRT-16, parents were instructed to best describe how *their child* has been feeling or acting. Total scores may range 0 to 64, with higher scores predicting greater risk for suicidality and frequency of suicidal ideations (Trombello et al., 2023).

**Emotion Dysregulation Inventory (EDI):** The EDI is a 24-item scale with two subscales that measure emotion regulation impairment symptoms often present in youth with autism spectrum disorder. Parents received the 7-item short form reactivity subscale (EDI-R-SF) and the 6-item dysphoria subscale (EDI-D), for a total of 13 Likert-based items answered from 0 (Not At All) to 4 (Very Severe). Each subscale measures one independent, but correlated, factor; they are scored separately and not combined for a total score (Mazefsky, Conner, et al., 2021; Mazefsky, Yu, et al., 2021).

**Family Index of Risk for Mood (FIRM):** The FIRM is a parent-report tool used to assess a youth's risk for mood disorders based on the informant's family history (grandparents, parents, aunts/uncles, brothers/sisters, and children not included in the study) of suicide, alcohol/drug use, mental health hospitalization, depression, and bipolar disorder. The measure is formatted as a 25-cell "check all that apply" grid with two yes/no items asking the parent informant if they have ever been told by a health professional that they themselves have bipolar/manic disorder or depression. Higher scores increase the likelihood rate for BSDs and any mood disorder (Algorta et al., 2013).

**Nationwide Gauge of Emotional Severity (N-Gauge):** The N-Gauge is a single item, global measure that asks parent informants to rate their level of concern about their child's mental health over the past week on a 0 to 5 scale (0: No Problems, 1: Mostly Okay, 2: Mild, 3: Moderate, 4: Bad, 5: Very Bad; McClellan et al., 2025).

**Nationwide Quality of Life Scale (NQLS):** The NQLS is a 7-item brief scale used to assess well-being and quality of life in children and adolescents (Covarrubias & Fristad, 2025). The parent report NQLS was adapted for this study, as the original NQLS is a youth self-report measure. Parents were asked "How satisfied are you with your child's… friendships, family relationships, grades, overall school functioning, health, life in general, and (anticipated) future", ranking each on a 0 (Very Unsatisfied) to 4 (Very Satisfied) Likert scale.

**Pediatric Symptom Checklist (PSC-17):** The PSC-17 is a 17-item multi-informant screening measure to assess internalizing (PSC-17-I, 5 items), externalizing (PSC-17-E, 7 items), and attention (PSC-17-A, 7 items) symptoms of various mental and behavioral disorders in youth. Each item belongs to one of the three subscales and asks the respondent to select whether the behavior occurs Never (0), Sometimes (1), or Often (2). Higher subscale scores and total measure scores indicate increased likelihood of psychosocial problems.

**PROMIS Pediatric Sleep-Related Impairment (PROMIS-SRI):** The PROMIS-SRI assesses perceptions of impairment due to sleep problems, such as "daytime sleepiness, sleep offset, and the impact of sleepiness on cognitive functioning, affect and behaviors, and daily activities" (Forrest et al., 2018). Parents completed the 13-item Parent Proxy Item Bank v1.0, which inquiries about the frequency of their child's sleep problems over the past week (1: Never, 2: Almost Never, 3: Sometimes, 4: Almost Always, and 5: Always).

**School Demographics, Mental Health Diagnoses, and Treatment History:** Parents answered 12 questions regarding their child's academic experience (type of school, current year/grade in school, academic performance, and enrollment in special accommodation resources), clinical diagnoses (anxiety disorders, ADHD, BD, autism, aggression or disruptive behavior disorder, sleep disorder, learning disability, or none of the above), and mental health treatment history (prescription medication, non-prescription medications, herbs, or supplements, and therapy attendance).

**Vanderbilt ADHD Diagnostic Parent Rating Scale (VADPRS):** The full National Initiative of Child Healthcare Quality (NICHQ) VADPRS is a 55-item parent-report scale used to identify symptoms of ADHD, externalizing disorders, and overall functioning in youth over a 6-month period. In the current study, parents completed 26 items: the first 18 items of the Symptoms section (9 predominantly inattentive, 9 predominantly hyperactive/impulsive) and the Performance section (Items 48-55). The administered Symptoms items, covering all 18 *DSM-IV* criteria for ADHD, are answered on a 4-point Likert scale (0: Never, 1: Occasionally, 2: Often, 3: Very Often) and can be scored separately from the rest of the Symptoms section for an ADHD total score, predominantly inattentive items only, and predominantly hyperactive/impulsive items only. The Performance section is on a 5-point Likert scale (1: Excellent, 2; Above Average, 3: Average, 4: Somewhat of a Problem, 5: Problematic) and also can be scored independently (Wolraich et al., 2003).

**Very Quick Inventory of Depressive Symptomatology (VQIDS):** The VQIDS-SR is a 5-item short form of the QIDS-16 used to measure the core major depression symptom domains: sad mood, self-outlook, involvement, fatigue, and psychomotor slowing (De La Garza et al., 2017). Parents completed the VQIDS-P-A, an unpublished parent-rated version of the self-report VQIDS created by the authors of the original measure.

*Statistical Analyses*

Data management and statistical analyses were conducted in RStudio v.4.5.2. The complete markdown files listing the packages and versions used are in the supplemental materials. All analyses used complete cases: a respondent was included in the analysis of a form only if every item of that form was answered, and no prorating was applied (2,229 of the 2,252 general-population respondents and 522 clinical respondents had complete CMRS-P-21 data; the CMRS-P-10 analyses used the same cases). Analyses of the general-population sample applied the YouGov post-stratification weights; the clinical sample, for which no weights exist, was analyzed unweighted (weight=1), so the multigroup analyses combine a weighted general-population sample with an unweighted clinical sample. Weighted estimation used weighted polychoric correlations (wCorr), lavaan sampling weights with WLSMV, mirt survey weights (pseudo-maximum likelihood), lordif weights, prior weights in the GAMLSS models (rescaled to sum to the sample size), weighted Pearson correlations, and Kish's effective sample size in place of *N* for test statistics and standard errors. Limited-information fit statistics ($M_2$) and quantile-residual diagnostics are not defined for weighted likelihoods, so they were computed on pseudo-populations drawn with probability proportional to the weights at the effective sample size (medians across repeated draws). The Kish design effect of the weights in the analytic sample was 1.93, which corresponds to an effective sample size of about 1,156 of 2,229 respondents. The design effect is the factor by which the variance of an estimate is inflated by unequal weights, relative to a simple random sample of the same size: a value of 1.0 means that the weights cost no precision, values between about 1.5 and 2.5 are common for post-stratified online panel samples and mean that standard errors would be about 1.2–1.6 times (the square root of the design effect) larger if the weights were used, and values above about 3 indicate that a few respondents carry so much weight that weighted estimates become unstable. The effective sample size (*N* divided by the design effect) is the size of a simple random sample that would carry the same precision. Unweighted analyses of the same models are reported in the supplemental analysis files and are compared with the weighted results in the Limitations.

*Factor Structure.*

We used the clinical sample to establish the baseline model for the factor structure, and then conducted multigroup analyses treating the clinical sample as the reference. We chose this approach because the clinical sample was enriched for cases with mood disorder, increasing both the base rate and the availability of more severe presentations that would add variance in items that might be rarely endorsed in the general population. Because the CMRS-P is designed to measure a clinical construct and the typical use case would be in clinical settings, it makes sense to treat the factor structure in a clinical sample as the anchor model. The 25:1 participant to item ratio was more than sufficient for adequate factor estimation (MacCallum et al., 1996). Because responses were ordered categorical, analyses in both samples were based on polychoric correlation matrices (Olsson, 1979). Models were estimated with WLSMV and indicators specified as ordered (Muthén et al., 1997). Confirmatory factor analysis of the CMRS-P-21 was conducted both samples starting with four correlated factors and testing bifactor and higher-order factor models, assigning items based on the EFA results in the clinical sample to Elated Mood, Activity, Irritable Mood, and Strange Thoughts and Behaviors. Item 13 (sexually inappropriate behavior) was assigned to Strange Thoughts and Behaviors, leaving eight items on Elated Mood, six on Activity, three on Irritable Mood, and four on Strange Thoughts and Behaviors. Fit was interpreted using CFI, TLI and SRMR; RMSEA is reported descriptively, recognizing that with WLSMV estimation it is more likely to indicate a floor on model fit rather than a ceiling (Savalei, 2021). All solutions in both samples were screened for inadmissible estimates, defined as standardized loadings outside bounds or negative residual variances. Persistent convergence and Heywood cases were treated as equivalent grounds for rejection. Structural equivalence was assessed through a multigroup configural model fit jointly across both samples (Millsap & Tein,

2004). Group-factor distinctiveness was evaluated in both samples using omega-hierarchical-subscale (Rodriguez et al., 2016) and the *H* coefficient of factor determinacy (Hancock & Mueller, 2001). General-factor dominance was additionally quantified by explained common variance and omega-hierarchical (Reise et al., 2010; Rodriguez et al., 2016), with the proportion of uncontaminated correlations reported as a qualifier on their interpretation against published benchmarks (Reise et al., 2013). Because the Elated Mood factor was nearly redundant with the general factor in both samples (second-order loadings ≥ .97), we also fit a bifactor-(S-1) model (Eid et al., 2017) separately in each sample, in which Elated Mood is the reference domain that defines the general factor and Activity, Irritable Mood, and Strange Thoughts and Behaviors are orthogonal specific factors (the general-population model used the sampling weights).

*Item Response Theory*

A graded response model (Samejima, 1968) was fit to both CMRS versions in both samples. A single unidimensional calibration of all 21-items served as the primary model, with four supplementary factor-level models calibrated on their own items only. The 10-items also were evaluated separately as a set. Overall model fit was evaluated with the limited-information $M_2$ ($C_2$) statistic and the RMSEA, SRMSR, TLI and CFI based on it (Cai & Hansen, 2013); when the unidimensional model fit poorly, a bifactor graded response model with a general factor and the four group factors was fit for comparison. Item discrimination was interpreted against conventional descriptive bands (Baker & Kim, 2017). Model adequacy was assessed at three levels. Item fit used the root mean square deviation, with values above 0.10 flagged (Yamamoto et al., 2013), computed in the clinical sample against a matched-sample-size reference (the general-population RMSD distribution across 100 random subsamples of $n$=522, drawn with probability proportional to the weights), so that an item was called substantively different between samples only when its clinical RMSD exceeded both 0.10 and the 97.5th percentile of that reference, supplemented by option characteristic curve plots as a visual diagnostic. Local independence used Yen's *Q3* residual correlations across all item pairs, flagging a pair when |Q3| exceeded the mean absolute Q3 by more than .20 (Feuerstahler et al., 2019; Yen, 1984). Person fit used the standardized *lz* statistic, with values exceeding an absolute value of 3 flagged (Drasgow et al., 1985). Empirical reliability of the resulting latent trait estimates was computed from each calibrated model and expected-score functions were used to relate trait estimates to raw sum scores (Bock & Mislevy, 1982), computed with a threshold-count-general specification appropriate to four-category items.

*Measurement Equivalence Between Samples*

Differential item functioning (DIF) between the general-population and clinical samples was screened with ordinal logistic regression on all 21-items (lordif; $R^2$ change ≥ .02, α=.01; Choi et al., 2011). Invariance was then tested with a configural–metric–scalar sequence of multiple-group graded response models (Chalmers, 2012), taking a decrease in CFI greater than .010 between nested models as evidence against invariance (Chen, 2007; Cheung & Rensvold, 2002). Because full scalar invariance did not hold, we fit a partial-scalar model in which the items not flagged for DIF served as anchors constrained equal across samples, while the flagged items' parameters were estimated freely in each sample; the general-population sample defined the reference metric (latent mean 0, variance 1) and the clinical sample's latent mean and variance were estimated freely. This links the two samples to a common latent metric, so that expected scores implied by each sample's item parameters can be compared directly. The linking rests on the assumption that the anchor items function equivalently in both samples. That assumption cannot be tested with these data, and a non-invariant anchor would bias the latent mean difference and the expected-score comparisons. Differential test functioning was summarized as the difference between the two samples' expected total scores across the latent range ($\theta = -3$ to 3), as signed and unsigned areas between item response functions (Raju, 1988, 1990), and as a net difference weighted by the clinical sample's latent distribution, and was interpreted against the reliable-change and minimal important difference thresholds for the total score. Invariance of the higher-order and bifactor-(S-1) factor models was tested with multiple-group WLSMV models for ordered-categorical items (theta parameterization, with identification following Wu & Estabrook, 2016), adding constraints in sequence: item thresholds, first-order loadings, second-order loadings (higher-order model), and item intercepts, with the same ΔCFI and ΔRMSEA criteria ($\Delta\text{CFI} \leq -.010$ with $\Delta\text{RMSEA} \geq .015$; Chen, 2007) and with the DIF-flagged items freed in partial models. Because invariance of measurement parameters does not constrain factor variances, we also compared the variances of the first-order factors that the general factor does not explain, and the resulting standardized second-order loadings, between samples.

*Retest Stability and Clinical Benchmarks*

A random subset of the general-population sample (167 parents with matched baseline and retest data) completed the CMRS-P again about two to three weeks later. Retest dependability was estimated as the absolute-agreement intraclass correlation, ICC(A,1), which is penalized by systematic shifts as well as by inconsistency. The standard error of measurement was $\text{SEM} = SD \times \sqrt{(1 - \text{ICC})}$, using the baseline *SD*, and the reliable change index at the 95% level was $\text{RCI} = 1.96 \times \sqrt{2} \times \text{SEM}$ (1.645 for the 90% level; Jacobson & Truax, 1991). For comparison, the same thresholds were also computed from internal-consistency (omega-based) standard errors, which exclude time-varying error. The minimal important difference (MID) was set at 0.5 *SD* (Norman et al., 2003). Clinical-significance benchmarks followed Jacobson and Truax (1991): Method A, the clinical mean minus 2 *SD*; Method B, the non-clinical mean plus 2 *SD*; and Method C, the weighted midpoint between the two means. The retest analyses used the baseline weights of the 167 retest respondents (effective *N*=81); intervals for the ICC were obtained by bootstrapping pairs.

*Reliability and Validity*

We calculated internal consistency (average item correlation, Cronbach's alpha, α; McDonald's omega, ω) for the full (21-item) and brief (10-item) CMRS-P within sample groups and across the entire sample using the *psych* package in *R*. In Likert-scale questions, Cronbach's α above .90 is considered "excellent", .80 to .89 is "good", .70 to .79 is "acceptable", .60 to .69 is "questionable", .50 to .59 is "poor", and values below .50 are "unacceptable". In Likert data, McDonald's ω provides a more accurate estimate of reliability, as it is not dependent on normal distribution of scores or unidimensionality (Kalkbrenner, 2024). Omega also is more appropriate when there are multiple factors underlying an item set, as alpha assumes a single factor.

We also assessed convergent and discriminant validity of the full and brief CMRS-P in comparison to other measures in the BENCHMARK assessment battery. Pearson's correlation coefficient tests (*r*) determined the relationships between total scores of the CMRS-P-21 and CMRS-P-10 and measures of co-morbid symptom domains administered in the BENCHAMRK study. Correlation strengths were interpreted using Cohen's criteria: 0 (no correlation), ~ 0.1 small, ~0.3 medium, and ~0.5 large (Cohen, 1988). Criterion correlations were weighted Pearson correlations computed on respondents with complete data on both CMRS-P forms and all non-sex-specific criterion scales (listwise deletion), and the difference between the 21- and 10-item correlations with each criterion was tested with Steiger's (1980) Z for dependent, overlapping correlations, with the Kish effective sample size in place of *N*, and summarized as Cohen's *q*. We anticipated that the CMRS scores would show large correlations with measures of depression, internalizing, and family history of mood disorder, and at least moderate correlations with measures of sleep and attention problems. Mania scales also show high correlation with measures of externalizing, and more small correlations with age and sex (E. A. Youngstrom et al., 2020, 2021). We also expected small correlations with pubertal status.

*Continuous Norming*

We established age- and sex-specific national norms for the CMRS-P with Generalized Additive Models for Location, Scale, and Shape (GAMLSS; Rigby & Stasinopoulos, 2005), a continuous norming approach that can achieve higher norming accuracy even with samples substantially smaller than ours (Frazier et al., 2026). Norms were derived from the 2,229 complete cases, with the YouGov weights (rescaled to sum to *N*) entered as prior weights in every model, so that the fitted percentiles describe the weighted national population rather than the sample as recruited. Because total scores are integer-valued counts with a floor at zero, only discrete (integer-valued) distribution families were considered: Poisson (PO), negative binomial types I and II (NBI, NBII), Poisson inverse Gaussian (PIG), beta binomial (BB), and the zero-inflated and zero-adjusted versions of these (ZIP, ZAP, ZINBI, ZANBI, ZIPIG, ZAPIG, ZABB). Model building followed two steps. First, to limit overfitting of the weighted upper percentiles, which rest on few effective observations at any single age, the age function was restricted to low-dimensional polynomials and chosen by one rule applied uniformly to both forms: sex-only, linear, quadratic and cubic age (centered at 12 years), each with and without age-by-sex terms, were fit to the weighted data with a negative binomial distribution and compared by 10 × 10 repeated, stratified 10-fold cross-validation of the weighted held-out log-likelihood of the full score distribution and by the BIC of the weighted fit; we retained the least complex age function whose cross-validated performance was within one standard error (of the paired difference) of the best, accepting added complexity only if the BIC also improved. Flexible P-spline smoothers were fit initially and are reported only as an exploratory comparison. This rule was specified after the CMRS-P results; the preregistered cubic age-by-sex form is reported as a sensitivity analysis [confirm wording against the preregistration]. Second, with the mean structure fixed ($\log \mu = \beta_0 + \beta_1 \cdot \text{sex} + \beta_2 \cdot \text{age} + \beta_3 \cdot \text{sex} \times \text{age}$; constant dispersion σ; constant zero-mass probability ν for zero-inflated or zero-adjusted families), we compared the candidate distributions using global and local fitness indices, including the BIC, worm plots (van Buuren & Fredriks, 2001), and Q-statistics computed from randomized quantile residuals (median *p*-values over 25 refits, from pseudo-populations drawn in proportion to the weights at the effective sample size; Timmerman et al., 2021, 2026), and selected the best-fitting model; when BIC and the residual diagnostics disagreed, the diagnostics took precedence. For the selected model, percentiles (5th, 10th, 25th, 50th, 75th, 90th, 95th) were taken from the fitted distribution as the smallest integer score at which the cumulative probability reached the percentile, plotted separately for males and females, and tabulated for clinical reference with the corresponding T scores ($T = 50 + 10 \times z$, so that the 5th to 95th percentiles correspond to T = 34, 37, 43, 50, 57, 63, and 66). Uncertainty was quantified with a nonparametric bootstrap of respondents (1,000 resamples, refitting the weighted model each time) and is reported as percentile 95% intervals. Sensitivity analyses compared the primary norms with (a) the same model without weights; (b) the weighted cubic age-by-sex model; (c) a covariate-standardized alternative, in which an unweighted model that included the weighting variables (parent gender, education, Hispanic ethnicity, race, region, and parent age centered within child age) was fit and its predicted score distributions were mixed over the weighted national covariate composition before percentiles were extracted; and (d) the flexible weighted P-spline model.

#### Results

##### *Age-Group Descriptive Comparisons*

In parents of children aged 6–11 (*n*=993), weighted CMRS-P-21 and -10 scores averaged 8.77 (*SD*=8.75) and 4.01 (*SD*=4.01), respectively. In parents of adolescents aged 12–18 (*n*=1,236), scores averaged 8.38 (*SD*=9.16) and 4.08 (*SD*=4.50). Descriptive statistics and reliability by age group are in Table S2, and clinical benchmarks separated by age group are in Table S3 of the Supplemental Materials.

Additionally, item means and standard deviations by sample age group for the 21-items of the CMRS-P are located in Table S4 of Supplemental Materials. Item endorsements (defined as selecting a 1, 2, or 3, as opposed to selecting 0 or skipping the item entirely) by age group were generally similar in numbers; however, items describing irritability and mood lability (e.g., "irritable, cranky, or mad," endorsed by 56% of weighted parents of adolescents vs. 46% of parents of children) were endorsed at higher frequencies by parents of adolescents, and items describing high energy and hyperactivity, symptoms shared with attention disorders (e.g., "periods of too much energy," 40% vs. 57%), were endorsed more frequently by parents of children.

##### *Factor Analyses*

The one-factor model fit acceptably by CFI and TLI in the general-population sample (CFI=.986, TLI=.985, RMSEA=.048) and poorly in the clinical sample (CFI=.947, RMSEA=.112; Table S5). The four-factor structure fit well in both samples (CFI=.989 in the clinical sample and .994 in the general-population sample). A multigroup configural model fit jointly across both samples also fit well (CFI=.993, TLI=.992, RMSEA=.036, SRMR=.056), with no inadmissible estimates in either group. Fit of the one-factor model within each content scale is shown in Table S7.

***Multigroup Factor Structure***

The bifactor-(S-1) model was the best-fitting admissible model in both samples (main-text Figure 2; Table S5). Detailed multigroup invariance comparisons are reported in Table S6.

Measurement invariance did not imply identical factor variances. Figure S4 illustrates the higher-order structure in the general-population sample; main-text Table 3 compares disturbance variances across samples. In the scalar higher-order model, disturbance variances—the portions of first-order factor variance not explained by the general factor—were larger in the clinical sample: Activity, .822 versus .438; Irritable Mood, .585 versus .232; and Strange Thoughts and Behaviors, .438 versus .053 (1.9, 2.5, and 8.3 times larger, respectively; main-text Table 3). Elated Mood had essentially no disturbance variance in either sample, with a small negative clinical estimate (−.003). General-factor variances were similar (1.333 versus 1.457). The larger clinical disturbances account for lower standardized second-order loadings despite equal unstandardized loadings. Constraining disturbance variances to equality changed CFI by only −.003, illustrating that global fit-change criteria can overlook these differences.

For the higher-order multigroup CFA, successive equality constraints on thresholds, first-order loadings, second-order loadings, and item intercepts changed weighted CFI by .000, −.001, −.001, and −.004, respectively ($\Delta$RMSEA $\leq$ .007). Constraining first-order factor intercepts changed CFI by a further −.005. The unweighted analyses gave the same substantive conclusion. For the S-1 model, weighted metric $\Delta$CFI was −.002; scalar $\Delta$CFI was −.009 with $\Delta$RMSEA=.016, whereas unweighted changes were −.007 and .014. Partial scalar constraints, freeing the three population-DIF items, yielded weighted ΔCFI=−.006 and ΔRMSEA=.011 (Table S6). These CFA results meet the stated joint rule requiring both a CFI decrease of at least .010 and an RMSEA increase of at least .015 to reject invariance, although the weighted full-scalar RMSEA change warrants qualification.

*Essential Unidimensionality and Item Response Theory*

Table S9 reports general-factor dominance and specific-factor distinctiveness. The general-population model had ECV=.857 and omega-hierarchical=.936; no group factor met all distinctiveness criteria. The clinical sample retained more specific-factor variance (ECV=.66; omega-hierarchical=.85). Overall IRT fit and empirical reliability are reported in Table S10, factor-level calibrations in Table S11, and item parameters and diagnostics in Tables S12–S14.

Shared findings. The single unidimensional calibration yielded empirical reliability of .905 in the clinical sample and .865 in the weighted general-population sample (Table S10). Absolute fit differed between samples. It was acceptable in the general-population sample (M2-based RMSEA=.080, CFI=.966, TLI=.963) but not in the clinical sample (RMSEA=.111, CFI=.886, TLI=.874), and the same pattern held for the 10-item form (general population RMSEA=.088, CFI=.960; clinical RMSEA=.108, CFI=.890). The clinical misfit reflects multidimensionality rather than a defect of the total score: a bifactor graded response model with a general factor and the four group factors fit the clinical data well (M2=502.7, df=168, RMSEA=.062, CFI=.968). Person fit was unremarkable in both samples, with 1.15% of clinical respondents and 1.10% of weighted general-population respondents flagged. Items 20 (voices) and 21 (see) were flagged for local dependence in the clinical sample, consistent with shared perceptual-disturbance content, but not in the weighted general-population sample, and item 11 (distracted) was flagged for item fit in both. Activity was the most reliably measured factor-level calibration in both samples (Table S11).

***Clinical Sample***

Discrimination was very high for seven items and high for three (items 1, 16 and 18), with ten items in the moderate band and item 13 (sex) the single low discriminator (a=0.65; Table S12). Local dependence was more extensive than in the general-population sample (Q3 mean=−.028, maximum=.622), flagging six pairs: items 20 and 21 (.622), all three pairwise combinations of the Irritable Mood items 2, 16 and 18 (.416 to .493), and items 8 and 16 (−.302) and 10 and 17 (−.305). Item fit flagged items 11, 15, 16 and 18 above the RMSD threshold; items 15, 16 and 18 also exceeded the matched-sample-size reference and were therefore clinical-specific (Table S13). Among factor-level calibrations, empirical reliability was .767 for Elated Mood, .864 for Activity, .812 for Irritable Mood and .551 for Strange Thoughts and Behaviors, where item 13 was the weakest discriminator (a=0.55).

***General-Population Sample***

Discrimination was very high for 16 of 21-items. The five weaker items were items 1 (excited), 2 (irritable) and 11 (distracted), all moderate, and items 3 (anything) and 5 (sleep), both high, corresponding to the lowest general-factor loadings (Table S14). Local dependence was minimal (Q3 mean=−.040, maximum=.287), and no pair exceeded the flag threshold (.291). Item fit flagged two items above the threshold, items 2 and 11, again the weakest general-factor loaders. Among factor-level calibrations, empirical reliability was .710 for Elated Mood, .804 for Activity, .683 for Irritable Mood and .417 for Strange Thoughts and Behaviors. The Strange Thoughts and Behaviors items discriminated most strongly of any factor (a=2.74 to 5.11, including item 13 at a=3.13), but their information is concentrated at the upper end of the trait, so reliability across the general-population range was low. Conditional reliability across the score range is shown in main-text Figure 5 (the general-population panel uses the weighted calibration).

Item severity ordering. On a common metric defined by the general-population sample, item severity (the mean of the three GRM thresholds, $\bar{b}$) ranged from 1.05 for easily distracted (item 11) to 2.79 for sexually inappropriate (item 13) (Table S22, main-text Figure 3). The attention and energy items were the least severe and the psychosis-type and sexually inappropriate items the most severe; the middle of the order is a band of overlapping items (rank intervals in Table S22). The two samples gave nearly the same order (Spearman $\rho = .92$) because 18 items were constrained equal; hearing voices and seeing things (items 20 and 21) were much more severe on the clinical calibration ($\bar{b} = 3.94$ and 4.31) than in the general population (2.43 and 2.27), and rage attacks (item 16) less severe (1.20 vs. 2.01), consistent with the DIF reported above.

*Differential Item and Test Functioning*

**Sample DIF.** Three of 21 items showed differential item functioning (DIF) between the clinical and weighted general-population samples (Table S15, Panel A). At the same underlying severity, parents in the clinical sample endorsed rage attacks/prolonged tantrums (item 16) more strongly, and hearing voices (item 20) and seeing things (item 21) less strongly. Changes in $R^2$ were .041, .029, and .034, respectively. Grandiose abilities (item 3), flagged without weights, fell just below the effect-size threshold when weights were applied.

**Age DIF.** Differential item functioning across three child-age strata (6–9 years, n=675; 10–12 years, n=531; 13–18 years, n=1,023) was screened with the same ordinal logistic regression procedure used for sample DIF (lordif, graded response model, McFadden $R^2$ change $\geq .02$, $\alpha = .01$), applied to the 2,229 complete cases with the YouGov weights in the primary analysis and without weights for comparison (Table S23). In the weighted analysis no item was flagged; the largest $R^2$ changes were .018 for item 18 (rapid mood swings), .017 for item 6 (too much energy), and .017 for item 2 (irritable, cranky or mad). In the unweighted analysis two mood-lability items were flagged, item 2 (irritable, cranky or mad) ($R^2$=.032) and item 18 (rapid mood swings) ($R^2$=.028), both running higher in the 13–18 stratum than in the 6–9 stratum at the same latent severity (linked signed area 0.26 and 0.10 points). The weighted age screen was unstable: in 60 weighted bootstrap resamples (respondents resampled with their weights), items were flagged as follows: item 18 in 60%, item 20 in 52%, item 6 in 40%, item 21 in 38%, item 13 in 37%, item 2 in 33%, and item 14 in 32% of resamples, although no item was flagged in the full weighted sample; the instability reflects the effective sample size of about 1,156 (of 2,229) and the heavy weights, so the weighted age-stratum result should be read as 'no consistent age DIF' rather than as evidence that individual items are free of it. The total-score impact was small. With the two flagged items freed in a linked partial-scalar model (13–18 vs. 6–9 years, unweighted), expected total scores differed by 0.45 points on average across $\theta = -3$ to 3 (largest difference 0.79 points at $\theta$=0.25), and the older stratum's latent mean was 0.41 SD lower; in the weighted analysis, freeing the two items closest to the threshold changed expected totals by 0.19 points on average (largest 0.70 points at $\theta$=2.55). All of these differences are well below the 95% reliable-change threshold (14.47 points) and the minimal important difference (4.50 points).

**Sex DIF.** No item showed DIF by child sex (1,171 boys, 1,058 girls) in either the weighted or the unweighted analysis (Table S23); the largest $R^2$ changes were .010 for item 6 (too much energy) and .010 for item 18 (rapid mood swings) in the weighted analysis and .008 (unweighted), all far below the .02 criterion, and no item was flagged in more than 10% of 60 weighted bootstrap resamples. Freeing the two items closest to the criterion in a linked model changed expected total scores by 0.07 points on average (weighted; largest difference 0.21 points at θ=2.30) and 0.04 points (unweighted), and the latent mean difference between girls and boys was 0.01 SD (weighted).

In the unidimensional IRT models, full scalar invariance failed (ΔCFI=−.020); freeing the three DIF items improved fit but left the partial-scalar model just outside the .010 tolerance (ΔCFI=−.012; Table S15, Panel B). Nevertheless, the small total-score effects converged with the multigroup CFA evidence for approximate measurement equivalence. Linking on the 18 items without detected sample DIF placed both samples on a common θ metric, on which the clinical mean was 0.67 SD higher. This comparison depends on the anchor items functioning equivalently across samples.

Main-text Figure 4 compares the expected 21-item totals for youths at the same underlying severity: the close curves indicate that sample membership changes the expected total little over most of the trait range. The clinical curve is slightly higher near average severity (maximum difference +0.82 points) but lower at the upper extreme (−2.42 points at θ=3). The mean absolute difference was 0.56 points across θ=−3 to 3 and 0.42 within θ=±2, below the 4.50-point minimal important difference and 14.47-point reliable-change threshold. Thus, most of the approximately seven-point observed mean difference reflected severity rather than differential test functioning.

*Reliability*

Main-text Table 2 reports internal consistency, retest stability, standard errors of measurement, and reliable-change thresholds for both forms. Omega total includes reliable variance attributable to general and group factors; omega-hierarchical isolates the general-factor contribution (Table S9).

The unidimensional IRT models instead treat the items as indicators of one general severity factor and estimate the precision of the scale's general-factor score. Empirical reliability was .865 and .768 for the 21- and 10-item forms in the general population and .905 and .809 in the clinical sample (Table S10). main-text Figure 5 shows how conditional reliability changes across underlying severity: both forms are most precise in the middle-to-upper range and less precise at the extremes, with the 21-item form maintaining higher reliability over a broader range. Main-text Figures 5 and 4 use the linked clinical and weighted general-population calibration, with the 18 items without detected sample DIF anchoring a consistent θ metric across samples.

Over approximately two to three weeks, 167 parents provided matched retest data (weighted effective N=81). Retest correlations were .665 for the CMRS-P-21 and .690 for the CMRS-P-10; absolute-agreement ICCs were .664 (95% CI [.555, .788]) and .679 [.516, .829], respectively (main-text Table 2). Bland–Altman plots showed little average change (baseline minus retest: −0.21 and −0.16 points), but appreciable individual variation, with limits of agreement of −11.55 to 11.13 and −5.90 to 5.58 points (main-text Figure 6). Proportional bias was nonsignificant for the 21-item form (slope=−.110, p=.117) and significant for the 10-item form (slope=−.219, p=.001), for which retest scores tended to exceed baseline scores more at higher average severity. Stable group means therefore did not imply close agreement for every individual.

The standard errors of measurement in main-text Table 2 quantify uncertainty in individual scores in the scales' original units. Omega-based SEMs were 1.44 points for the 21-item form and 1.11 for the 10-item form; the retest-based SEMs were larger, 5.22 and 2.45 points, because they also incorporate variation across occasions. For monitoring change, the retest-based SEMs are the relevant estimates: after allowing for uncertainty at both assessments, changes exceeding 14.47 and 6.78 points meet the 95% reliable-change criterion (12.14 and 5.69 at 90%). The smaller omega-based thresholds would overstate confidence in longitudinal change. Reliable change also differs from the distribution-based minimal important difference (4.50 and 2.16 points), which does not establish that an individual change exceeds measurement error.

*Validity*

We used Fisher's *r*-to-*z* tests for independent correlations to determine if CMRS-P correlations differed significantly ($p < .05$) across age groups (6–11, 12–18 years; Fisher, 1921). Before correction, 7 of the 21 comparisons for the CMRS-P-21 and 5 for the CMRS-P-10 were significant at $p < .05$: the correlations with the PDS (which were small and positive in children and near zero or slightly negative in adolescents), with the emotion dysregulation scales (EDI-Reactivity $r$=.56 in children vs. .66 in adolescents; EDI-Dysphoria .54 vs. .65), and, to a lesser degree, with the CHRT-16 and VADPRS-Hyperactive subscale (CMRS-P-21 only for the latter). We then applied the Benjamini–Hochberg False Discovery Rate (FDR) procedure to reduce the risk of falsely reporting significant results given the large number of correlation comparisons (Benjamini & Hochberg, 1995). After adjustment, the CMRS-P-21 differences for the PDS (total $p$=.017, boys $p$=.007) and for EDI-Reactivity ($p$=.024) and EDI-Dysphoria ($p$=.027) remained significant, and for the CMRS-P-10 only the PDS total and boys differences remained ($p$=.041 and .039).

*Norms*

The linear age-by-sex model had the highest weighted held-out log-likelihood and the lowest BIC of the seven nested models; adding quadratic or cubic age terms lowered predictive performance (paired $\Delta$=−0.0021 and −0.0034 per observation for the CMRS-P-21, −0.0016 and

−0.0026 for the CMRS-P-10), and the sex-additive linear model did worse than a sex-only model, consistent with an age trend that differed by sex. The linear age-by-sex model outperformed the sex-only reference (paired Δ=0.0018 [SE 0.0006] for the CMRS-P-21; 0.0040 [SE 0.0008] for the CMRS-P-10), although its BIC advantage was only 1.1 points for the CMRS-P-21 (versus 10.0 for the CMRS-P-10). With this mean structure, the NBI distribution was retained for both forms (Table S19). ZANBI had the lowest BIC for the CMRS-P-21 and ZIPIG for the CMRS-P-10, but only 2.6 and 5.7 points below NBI, and the residual diagnostics did not favor them over NBI (Figures S1a and S1b). The center Q-statistic (Z1) was significant in 84%–100% of pseudo-population refits for every candidate, including NBI, which indicates that a linear age function does not reproduce all of the age-group means of the weighted data (notably the mid-adolescent rise seen with flexible smoothers); the dispersion, skewness and kurtosis statistics (Z2–Z4) were comparable across the leading candidates. Because the weighted diagnostics come from resampled pseudo-populations they are a coarser check than unweighted ones.

For males, the 95th percentile fell from 30 at age 6 to 23 at age 18, the 75th from 14 to 10, and the median from 7 to 5. In females, they rose: the 95th percentile from 22 to 30, the 75th from 10 to 14, and the median from 5 to 7; the two curves crossed at about age 12–13. A floor effect was apparent at the lower end of the distribution: the 5th percentile was 0 at every age in both sexes and the 10th percentile was 0 or 1. The bootstrap 95% intervals were widest at the ends of the age range: the 95th percentile was 30 [26, 35] in males and 22 [19, 25] in females at age 6, 26 [24, 28] and 25 [23, 28] at age 12, and 23 [18, 27] and 30 [24, 35] at age 18 (Table S21). These intervals reflect the weighting: the effective sample was 1,156 overall, but only about 34–69 respondents per sex within each year of age from 6 to 16, 27 (females) and 38 (males) at age 17, and 9 and 8 at age 18. The male decline was reproduced when the same model was fit without weights, when the weighted cubic model was used, and in the covariate-standardized analysis (95th percentile at ages 6, 12 and 18: 30, 25, 21 unweighted; 29, 27, 26 covariate-standardized), whereas the rise in females was not: unweighted female percentiles were flat (95th percentile 23 at ages 6, 12 and 18), and the covariate-standardized female percentiles did not rise (95th percentile 27, 25, 24) (Table S21; Figure S3). The female trend, especially above age 15, should therefore be interpreted cautiously. The flexible weighted P-spline norms, which we judged overfit, had 95th percentiles of 24–30 (males) and 23–29 (females) with an irregular mid-adolescent peak.

The CMRS-P-10 showed the same patterns. In males, the 95th percentile fell from 13 at age 6 to 11 at age 18 (75th percentile 6 to 5; median 3 to 2), whereas in females, it rose from 10 to 16 (75th percentile 4 to 8; median 2 to 4); the bootstrap interval for the female 95th percentile at age 18 was [13, 19]. A strong floor effect was similarly observed, as the 5th and 10th percentiles were 0 for both sexes at every age and the 25th percentile was 1.

For main-text Table 4, percentiles are the smallest integer score at which the cumulative probability from the fitted survey-weighted negative binomial (NBI) model reaches the percentile ($\log \mu = \beta_0 + \beta_1 \cdot \text{sex} + \beta_2 \cdot \text{age} + \beta_3 \cdot \text{sex} \times \text{age}$, constant $\sigma$). CMRS-P-21: $\beta_0 = 2.153$, sex (female) = -0.032, age (per 6 years) = -0.151, sex × age = 0.318, $\sigma = 0.919$; CMRS-P-10: 1.379, 0.026, -0.104, 0.374, $\sigma = 0.830$. Bootstrap 95% intervals, sensitivity analyses, and effective sample sizes by age and sex are in Table S21 and Figure S3; model selection is in Table S20. Estimates at ages 17–18 rest on few effective respondents (Table S21).

#### Tables and Figures

**Table S1**

*Item-Level Comparison, General Population vs. Clinical Sample in descending order of difference between samples*

| Item | Content[a] | Gen. *M* | Gen. *SD* | Clin. *M* | Clin. *SD* | Cohen's *d* | *p* (FDR) |
|---|---|---|---|---|---|---|---|
| 16[b] | Rage attacks/prolonged tantrums | .32 | .65 | 1.13 | 1.06 | 1.02 | <.001 |
| 18 | Rapid mood swings | .47 | .70 | 1.16 | 1.09 | .83 | <.001 |
| 3 | Grandiose abilities for age | .34 | .66 | .98 | 1.06 | .79 | <.001 |
| 7 | Talks too much/loud/fast | .64 | .81 | 1.28 | 1.10 | .71 | <.001 |
| 11 | Trouble staying on track | .87 | .86 | 1.49 | 1.06 | .67 | <.001 |
| 17 | Excessively silly/jokes/laughs | .41 | .69 | .89 | .93 | .62 | <.001 |
| 2[b] | Irritable, cranky, or mad | .67 | .76 | 1.15 | 1.00 | .57 | <.001 |
| 6 | Periods of too much energy | .61 | .77 | 1.08 | 1.11 | .53 | <.001 |
| 10 | Rushes around nonstop | .39 | .70 | .79 | .98 | .49 | <.001 |
| 15 | Foolish or risky behavior | .22 | .56 | .47 | .75 | .40 | <.001 |
| 19 | Suspicious or strange thoughts | .21 | .52 | .40 | .71 | .32 | <.001 |
| 8[b] | Racing thoughts | .51 | .77 | .75 | .95 | .29 | <.001 |
| 20[b] | Hears voices others don't | .11 | .43 | .25 | .61 | .28 | <.001 |
| 12[b] | Unusually productive/creative | .50 | .72 | .71 | .86 | .28 | <.001 |
| 9[b] | Jumps topic to topic | .57 | .78 | .79 | .95 | .26 | <.001 |
| 13[b] | Sexually inappropriate behavior | .13 | .45 | .23 | .64 | .20 | .001 |
| 4[b] | Unrealistic powers, acts on them | .16 | .49 | .25 | .64 | .18 | .002 |
| 21 | Sees things others don't | .12 | .44 | .20 | .54 | .18 | .002 |
| 14 | Overly outgoing with strangers | .21 | .55 | .28 | .61 | .13 | .018 |
| 5[b] | Needs less sleep, not tired | .42 | .68 | .49 | .83 | .10 | .079 |
| 1[b] | Super happy / on top of the world | .67 | .78 | .72 | .83 | .07 | .216 |

Note: [a]Item descriptions from Henry 2008 paper (Henry et al., 2008). [b]Items included in CMRS-P-10.

**Table S2**

*Descriptive Statistics and Reliability by Age Group: CMRS-P-21 and CMRS-P-10, General-Population and Clinical Samples*

| | General population | | | | Clinical sample | | | |
|---|---|---|---|---|---|---|---|---|
| | Children (6–11) | | Adolescents (12–18) | | Children (<12) | | Adolescents (12–18) | |
| **Metric** | **P-21** | **P-10** | **P-21** | **P-10** | **P-21** | **P-10** | **P-21** | **P-10** |
| *N* (complete cases) | 993 | 993 | 1,236 | 1,236 | 295 | 295 | 227 | 227 |
| Mean (*SD*) | 8.77 (8.75) | 4.01 (4.01) | 8.38 (9.16) | 4.08 (4.50) | 16.72 (11.14) | 6.92 (5.20) | 13.93 (9.77) | 5.93 (4.62) |
| Observed range; Median | 0–52; 6 | 0–24; 3 | 0–56; 5 | 0–28; 3 | 0–55; 15 | 0–24; 6 | 0–47; 12 | 0–25; 5 |
| Male score, *M* (*SD*) | 9.67 (9.42) | 4.40 (4.26) | 7.99 (8.99) | 3.71 (4.16) | 17.30 (11.28) | 7.07 (5.25) | 13.57 (9.39) | 5.47 (4.27) |
| Female score, *M* (*SD*) | 7.67 (7.73) | 3.54 (3.65) | 8.83 (9.34) | 4.50 (4.84) | 15.43 (10.78) | 6.59 (5.10) | 14.30 (10.18) | 6.39 (4.93) |
| Pearson's *r*, P-21 & P-10 | .95 | | .96 | | .96 | | .96 | |
| Mean inter-item correlation (polychoric) | .53 | .49 | .58 | .56 | .40 | .36 | .36 | .35 |
| Cronbach's alpha (α) | .92 | .82 | .93 | .86 | .90 | .80 | .88 | .77 |
| Omega total (McDonald's ω) | .97 | .92 | .98 | .94 | .95 | .88 | .95 | .87 |
| Empirical 95th percentile | 27 | 12 | 28 | 13 | 37.3 | 17 | 34 | 15.7 |

*Note.* P-21 = CMRS-P-21 total; P-10 = CMRS-P-10 total. Complete-case totals, as in main-text Table 2. General population: children 6–11 and adolescents 12–18 by parent-reported child age. Clinical sample: children are all youth under 12 (including 41 aged 4–5) and adolescents are 12–18. Mean inter-item correlations are polychoric. Cronbach's alpha is computed from Pearson correlations; omega total is from a Schmid-Leiman solution on the polychoric matrix (four factors for the 21-item form, two for the 10-item form), the same conventions as main-text Table 2. Retest-based reliable-change thresholds are not shown by age group because the retest subsample (*N*=167) is too small to split.

**Table S3**

*Clinical Significance Benchmarks by Age Group: CMRS-P-21 and CMRS-P-10*

| | All ages (6–18) | | Children | | Adolescents | |
|---|---|---|---|---|---|---|
| **Benchmark** | **P-21** | **P-10** | **P-21** | **P-10** | **P-21** | **P-10** |
| Minimal Important Difference (MID, 0.5 *SD*, general population) | 4.50 | 2.16 | 4.37 | 2.01 | 4.58 | 2.25 |
| Method A: away from clinical range (clinical *M* − 2 *SD*) | not reportable* | not reportable* | not reportable* | not reportable* | not reportable* | not reportable* |
| Method B: back into normal range (non-clinical *M* + 2 *SD*) | 26.53 | 12.68 | 26.27 | 12.04 | 26.70 | 13.08 |
| Method C: crossing closer to normal range (weighted *M*) | 11.73 | 5.18 | 12.26 | 5.28 | 11.07 | 4.99 |
| Empirical 95th percentile, general population | 28 | 12.9 | 27 | 12 | 28 | 13 |

*Note.* Jacobson & Truax (1991) nomothetic benchmarks computed within each age group from the general-population (non-clinical) and clinical complete-case samples. *Method A is not reportable: clinical *M* − 2 *SD* falls below the scale floor of 0 (about −5.6 for the 21-item form and −3.3 to −3.5 for the 10-item form). Method C is the *SD*-weighted mean of the clinical and non-clinical means. The all-ages column reproduces main-text Table 2.

**Table S4**

*Item-Level Means, Standard Deviations and Endorsement by Age Group, General-Population Sample*

| Item | Content | Children M (SD) | Adolescents M (SD) | % endorsed, children | % endorsed, adolescents | d | *p* (FDR) |
|---|---|---|---|---|---|---|---|
| 2 | Irritable, cranky, or mad | .56 (.70) | .74 (.79) | 45.7 | 56.0 | .24 | <.001*** |
| 18 | Rapid mood swings | .39 (.68) | .51 (.70) | 30.6 | 40.8 | .17 | .012* |
| 20 | Hears voices others don't | .07 (.33) | .13 (.48) | 5.0 | 8.3 | .15 | .025* |
| 13 | Sexually inappropriate behavior | .10 (.41) | .15 (.48) | 7.0 | 10.5 | .11 | .169 |
| 5 | Needs less sleep, not tired | .38 (.65) | .44 (.69) | 30.7 | 33.8 | .08 | .305 |
| 21 | Sees things others don't | .10 (.44) | .13 (.45) | 6.3 | 8.7 | .06 | .524 |
| 15 | Foolish or risky behavior | .20 (.53) | .22 (.58) | 15.6 | 16.0 | .04 | .675 |
| 4 | Unrealistic powers, acts on them | .15 (.46) | .16 (.50) | 11.1 | 11.4 | .03 | .695 |
| 19 | Suspicious or strange thoughts | .20 (.54) | .22 (.51) | 15.4 | 17.7 | .03 | .695 |
| 8 | Racing thoughts | .51 (.77) | .50 (.78) | 38.1 | 36.3 | −.01 | .803 |
| 3 | Grandiose abilities for age | .35 (.66) | .33 (.66) | 25.6 | 24.2 | −.02 | .749 |
| 14 | Overly outgoing with strangers | .22 (.58) | .20 (.54) | 15.6 | 14.4 | −.03 | .675 |
| 17 | Excessively silly/jokes/laughs | .43 (.72) | .40 (.68) | 31.9 | 30.5 | −.04 | .622 |
| 11 | Trouble staying on track | .90 (.88) | .85 (.84) | 62.5 | 61.3 | −.05 | .609 |
| 9 | Jumps topic to topic | .60 (.77) | .55 (.79) | 45.7 | 39.6 | −.06 | .471 |
| 16 | Rage attacks/prolonged tantrums | .34 (.65) | .30 (.65) | 26.6 | 22.1 | −.06 | .471 |
| 12 | Unusually productive/creative | .54 (.71) | .48 (.72) | 43.7 | 37.2 | −.09 | .280 |
| 1 | Super happy / on top of the world | .74 (.81) | .62 (.76) | 54.6 | 47.5 | −.16 | .019* |
| 7 | Talks too much/loud/fast | .73 (.84) | .58 (.79) | 52.7 | 42.4 | −.18 | .008** |
| 10 | Rushes around nonstop | .48 (.75) | .34 (.66) | 35.5 | 25.0 | −.20 | .003** |
| 6 | Periods of too much energy | .76 (.80) | .52 (.74) | 57.2 | 40.0 | −.30 | <.001*** |

*Note.* General-population complete-case sample (children 6–11, *n*=993; adolescents 12–18, *n*=1,236). Items are in descending order of Cohen's d, computed as (adolescent mean − child mean) / pooled *SD*, so positive values are higher in adolescents. *p* (FDR) = Welch t test, Benjamini–Hochberg adjusted across the 21-items; **p* < .05, ***p* < .01, ****p* < .001. % endorsed = percent selecting 1, 2 or 3.

**Table S5**

*Confirmatory Factor Structure and Model Fit, Clinical and General Population Samples*

| Model | $\chi^2$ | *df* | CFI | TLI | RMSEA [90% CI] | SRMR |
|---|---|---|---|---|---|---|
| Clinical sample (*N*=522, unweighted) | | | | | | |
| 1-factor | 1422.607 | 189 | 0.947 | 0.941 | 0.112 [0.107, 0.117] | 0.110 |
| 4-factor (correlated, manuscript structure) | 431.479 | 183 | 0.989 | 0.988 | 0.051 [0.045, 0.057] | 0.070 |
| Bifactor (4-group factors) — INADMISSIBLE[a] | N/A | N/A | 0.995 (not usable) | — | 0.037 (not usable) | — |
| Bifactor-(S-1), Elated Mood as reference (all items on g; 3 orthogonal specific factors) | 341.650 | 176 | 0.993 | 0.992 | 0.043 [0.036, 0.049] | 0.063 |
| Higher-order (4-first order factors) — RETAINED | 464.273 | 185 | 0.988 | 0.986 | 0.054 [0.048, 0.060] | 0.072 |
| General population sample (*N* = 2,229, weighted) | | | | | | |
| 1-factor | 1142.801 | 189 | 0.986 | 0.985 | 0.048 [0.045, 0.050] | 0.075 |
| 2-factor (correlated) | 795.976 | 188 | 0.991 | 0.990 | 0.038 [0.035, 0.041] | 0.061 |
| 3-factor (correlated, suggested by own EFA) | 694.972 | 186 | 0.993 | 0.992 | 0.035 [0.032, 0.038] | 0.058 |
| 4-factor (correlated) | 575.365 | 183 | 0.994 | 0.993 | 0.031 [0.028, 0.034] | 0.053 |
| Bifactor (4-group factors) | 492.114 | 168 | 0.995 | 0.994 | 0.029 [0.026, 0.032] | 0.050 |
| Bifactor-(S-1), Elated Mood as reference (all items on g; 3 orthogonal specific factors) | 529.676 | 176 | 0.995 | 0.994 | 0.030 [0.027, 0.033] | 0.052 |
| Higher-order (4-first order factors) — RETAINED | 629.680 | 185 | 0.994 | 0.993 | 0.033 [0.030, 0.036] | 0.056 |

Note: [a]The 4-group structure produces a Heywood case. The bifactor-(S-1) model loads all 21 items on a general factor anchored by the Elated Mood items (the reference domain) and adds orthogonal specific factors for Activity, Irritable Mood, and Strange Thoughts and Behaviors (main-text Figure 2); it converged without inadmissible estimates in either sample. All models are complete-case WLSMV solutions (general population *N*=2,229, estimated with sampling weights, effective *N*=1,156; clinical *N*=522, unweighted).

**Table S6**

*Measurement Invariance of the Higher-Order and Bifactor-(S-1) Models Across the Clinical and General-Population Samples*

| | | Weighted fit | | | Change vs. preceding model | | Unweighted change | |
|---|---|---|---|---|---|---|---|---|
| **Step** | **$\chi^2$ (df)** | **CFI** | **RMSEA** | **SRMR** | **ΔCFI** | **ΔRMSEA** | **ΔCFI** | **ΔRMSEA** |
| ***A. Higher-order model (four first-order factors, one second-order factor)*** | | | | | | | | |
| Configural | 1094.0 (362) | 0.992 | 0.038 | 0.059 | | | | |
| + Thresholds equal | 1111.0 (383) | 0.992 | 0.037 | 0.059 | 0.000 | −0.001 | 0.000 | −0.001 |
| + First-order loadings equal (metric) | 1202.2 (400) | 0.991 | 0.038 | 0.059 | −0.001 | 0.001 | 0.000 | 0.000 |
| + Second-order loadings equal | 1272.2 (403) | 0.991 | 0.040 | 0.060 | −0.001 | 0.001 | −0.001 | 0.001 |
| + Item intercepts equal (scalar) | 1667.7 (424) | 0.987 | 0.046 | 0.062 | −0.004 | 0.007 | −0.003 | 0.005 |
| + First-order factor intercepts equal | 2114.5 (427) | 0.982 | 0.054 | 0.065 | −0.005 | 0.007 | −0.003 | 0.006 |
| Partial scalar: flagged items free† | 1609.0 (415) | 0.987 | 0.046 | 0.062 | −0.004 | 0.006 | −0.002 | 0.004 |
| ***B. Bifactor-(S-1) model (Elated Mood as the reference general factor; three orthogonal specific factors)*** | | | | | | | | |
| Configural | 871.3 (352) | 0.994 | 0.033 | 0.054 | | | | |
| + Thresholds equal | 888.3 (373) | 0.994 | 0.032 | 0.054 | 0.000 | −0.001 | 0.000 | −0.001 |
| + Loadings equal (metric) | 1098.0 (403) | 0.993 | 0.035 | 0.056 | −0.002 | 0.004 | −0.001 | 0.002 |
| + Item intercepts equal (scalar) | 1985.5 (423) | 0.983 | 0.052 | 0.062 | −0.009 | 0.016 | −0.007 | 0.014 |
| Partial scalar: flagged items free† | 1626.6 (411) | 0.987 | 0.046 | 0.059 | −0.006 | 0.011 | −0.004 | 0.009 |

Note. Multiple-group WLSMV models for ordered-categorical items (theta parameterization; identification following Wu & Estabrook, 2016). The clinical sample (N = 522, unweighted) is the reference group; the general-population sample (N = 2,229) is weighted in the weighted columns and unweighted in the last two columns. †Items flagged for population-type DIF (Table S15) had their thresholds, loadings and intercepts freed: items 16, 20 and 21 (weighted) and items 3, 16, 20 and 21 (unweighted). Non-invariance is indicated by ΔCFI ≤ −.010 together with ΔRMSEA ≥ .015 (Chen, 2007; Cheung & Rensvold, 2002).

**Table S7**

*CMRS-P-21 Tests of Unidimensionality for Different Scale Scores*

| Scale | $\chi^2$ | *df* | CFI | TLI | RMSEA | SRMR |
|---|---|---|---|---|---|---|
| **General-Population Sample (General population)** | | | | | | |
| Total (21-items) | 1142.801 | 189 | .986 | .985 | .048 | .075 |
| Elated Mood (8) | 40.653 | 20 | .997 | .996 | .022 | .038 |
| Activity (6) | 88.825 | 9 | .994 | .990 | .063 | .048 |
| Irritable Mood (3)[a] | 0 | 0 | 1.000* | 1.000* | 0.000* | 0.000* |
| Strange Thoughts & Behaviors (4) | 1.392 | 2 | 1.000 | 1.000 | .000 | .013 |
| **Clinical Sample (Clinical)** | | | | | | |
| Total (21-items) | 1422.607 | 189 | .947 | .941 | .112 | .110 |
| Elated Mood (8) | 22.774 | 20 | .999 | .998 | .016 | .044 |
| Activity (6) | 23.715 | 9 | .998 | .997 | .056 | .031 |
| Irritable Mood (3)[a] | 0 | 0 | 1.000* | 1.000* | 0.000* | 0.000* |
| Strange Thoughts & Behaviors (4) | 6.597 | 2 | .995 | .986 | .066 | .061 |

Note. aIrritable Mood is a "just identified" model; no degrees of freedom (*df*) or $\chi^2$ are available. Each scale was fit as its own one-factor model on complete cases (general population *N*=2,229, weighted; clinical *N*=522, unweighted); item 13 is assigned to Strange Thoughts & Behaviors. $ECV_{SS}$: Explained Common Variance of a Specific Factor.

**Table S8**

*Standardized Higher-Order Model Loadings, Both Samples*

| Item | Content | Factor | Loading (Gen. Pop.) | Loading (Clinical) |
|---|---|---|---|---|
| 1 | Super happy / on top of the world | Elated Mood | .589 | .676 |
| 3 | Grandiose abilities for age | Elated Mood | .691 | .472 |
| 4 | Unrealistic powers, acts on them | Elated Mood | .861 | .626 |
| 5 | Needs less sleep, not tired | Elated Mood | .695 | .631 |
| 12 | Unusually productive/creative | Elated Mood | .716 | .765 |
| 14 | Overly outgoing with strangers | Elated Mood | .829 | .613 |
| 15 | Foolish or risky behavior | Elated Mood | .872 | .583 |
| 17 | Excessively silly/jokes/laughs | Elated Mood | .752 | .775 |
| 6 | Periods of too much energy | Activity | .828 | .837 |
| 7 | Talks too much/loud/fast | Activity | .820 | .841 |
| 8 | Racing thoughts | Activity | .826 | .872 |
| 9 | Jumps topic to topic | Activity | .812 | .872 |
| 10 | Rushes around nonstop | Activity | .895 | .818 |
| 11 | Trouble staying on track | Activity | .637 | .624 |
| 2 | Irritable, cranky, or mad | Irritable Mood | .713 | .736 |
| 16 | Rage attacks/prolonged tantrums | Irritable Mood | .873 | .854 |
| 18 | Rapid mood swings | Irritable Mood | .871 | .905 |
| 19 | Suspicious or strange thoughts | Strange Thoughts & Behaviors | .884 | .891 |
| 20 | Hears voices others don't | Strange Thoughts & Behaviors | .907 | .822 |
| 21 | Sees things others don't | Strange Thoughts & Behaviors | .933 | .796 |
| 13 | Sexually inappropriate behavior | Strange Thoughts & Behaviors | .842 | .481 |

Note: Item-level loadings on each of the four first-order factors, and each factor's own second-order loading on the general factor (g). Complete-case WLSMV estimates (general population *N*=2,229, weighted; clinical *N*=522, unweighted). ~~Item 13 is assigned to Strange Thoughts & Behaviors.~~

| Factor | Second-Order Loading (General population) | Second-Order Loading (Clinical) |
|---|---|---|
| Elated Mood | .994 | .973 |
| Activity | .871 | .816 |
| Irritable Mood | .856 | .714 |
| Strange Thoughts & Behaviors | .932 | .597 |

**Table S9**

*Variance Attributable to the General Factor and the Four Group Factors in the Higher-Order Model: General Population and Clinical Samples*

| | General population (N = 2,229) | | | Clinical (N = 522) | | |
|---|---|---|---|---|---|---|
| **Construct / Subscale** | **ω*H*/*HS*** | **$ECV_{SS}$** | ***H*** | **ω*H*/*HS*** | **$ECV_{SS}$** | ***H*** |
| General Factor (g) | .94 | .86* | .97 | .85 | .66* | .93 |
| Elated Mood (8 items) | .01 | .01 | .05 | .05 | .05 | .16 |
| Irritable Mood (3 items) | .23 | .27 | .40 | .43 | .49 | .61 |
| Strange Thoughts & Behaviors (4 items) | .12 | .13 | .32 | .54 | .64 | .73 |
| Increased Activity (6 items) | .22 | .24 | .53 | .31 | .34 | .64 |

*Note. ωH/HS = omega hierarchical (ωH for the general factor, ωHS for each group factor); $ECV_{SS}$ = specific-scale explained common variance; H = construct replicability index. *For the general factor (g), $ECV_{SS}$ represents global explained common variance (ECV). Indices are model-implied by the Schmid-Leiman transformation of the second-order model (four first-order factors loading on one general factor), fit by WLSMV on complete cases (general population N=2,229, weighted; clinical N=522, unweighted), with item 13 assigned to Strange Thoughts & Behaviors. Each item's general loading is its first-order loading times the factor's second-order loading, and its specific loading is its first-order loading times the square root of one minus the squared second-order loading. A second-order model is a bifactor model with proportionality constraints, so these values are not directly comparable to indices from a freely estimated bifactor model. Second-order loadings (general population / clinical): Elated Mood .99 / .97, Activity .87 / .82, Irritable Mood .86 / .71, Strange Thoughts & Behaviors .93 / .60. Benchmarks for specific-factor utility: ωHS ≥ .20, $ECV_{SS}$ ≥ .30, H ≥ .70. Elated Mood meets none in either sample, because its second-order loading leaves almost no residual variance; Strange Thoughts & Behaviors meets all three in the clinical sample but none in the general population.*

**Table S10**

*Whole-Scale Graded Response Model (GRM) Fit (21-Item and 10-Item Forms, Both Samples)*

| Sample | $M_2$ | *df* | RMSEA [90% CI] | SRMSR | TLI | CFI | Empirical Reliability |
|---|---|---|---|---|---|---|---|
| General population, 21-item ($N$ = 2,229) | 1582.386 | 189 | .080 [.076, .084] | .075 | .963 | .966 | .865 |
| Clinical, 21-item ($N$ = 522) | 1393.553 | 189 | .111 [0.105, 0.116] | .089 | .874 | .886 | .905 |
| General population, 10-item ($N$ = 2,229) | 350.493 | 35 | .088 [.080, .097] | .082 | .948 | .960 | .768 |
| Clinical, 10-item ($N$ = 522) | 246.575 | 35 | .108 [.095, .120] | .075 | .859 | .890 | .809 |

Note. Single-factor graded response model (mirt::mirt(), itemtype = "graded"); $M_2/C_2$ limited-information fit statistic (Cai & Hansen, 2013). $M_2/C_2$ was computed for all four models, including the clinical-sample 21-item GRM. General-population $M_2$-based indices are medians over 20 pseudo-populations drawn in proportion to the weights at the effective sample size, because limited-information statistics are not defined for weighted likelihoods; clinical values are unweighted. For the clinical 21-item sample a bifactor graded response model (general factor plus the four group factors) fit substantially better than the unidimensional model: $M_2$=502.7, *df*=168, RMSEA=.062 [.056, .068], SRMSR=.055, TLI=.961, CFI=.968.

**Table S11**

*Four Separate Per-Factor Graded Response Models (GRMs) Fit*

| Factor | Items | Empirical Reliability (General population) | Empirical Reliability (Clinical) |
|---|---|---|---|
| Elated Mood | 8 | .710 | .767 |
| Activity | 6 | .804 | .864 |
| Irritable Mood | 3 | .683 | .812 |
| Strange Thoughts & Behaviors | 4 | .417 | .551 |

Note: Each factor was calibrated as its own standalone GRM.

**Table S12**

*Graded Response Model Item Parameters: CMRS-P-21 and CMRS-P-10 (Clinical Sample)*

| | | CMRS-P-21 model | | | | CMRS-P-10 model | | | |
|---|---|---|---|---|---|---|---|---|---|
| # | Item | *a* | *b*1 | *b*2 | *b*3 | *a* | *b*1 | *b*2 | *b*3 |
| 1 | Super happy / on top of the world | 1.41 | -.12 | 1.66 | 2.57 | 1.33 | -.12 | 1.74 | 2.68 |
| 2 | Irritable, cranky, or mad | .99 | -.98 | .84 | 2.28 | 1.02 | -.95 | .83 | 2.23 |
| 3 | Grandiose abilities for age | .76 | -.33 | 1.42 | 2.74 | — | — | — | — |
| 4 | Unrealistic powers, acts on them | 1.19 | 1.66 | 2.80 | 3.63 | 1.23 | 1.62 | 2.74 | 3.54 |
| 5 | Needs less sleep, not tired | 1.22 | .74 | 2.05 | 2.85 | 1.19 | .76 | 2.10 | 2.92 |
| 6 | Periods of too much energy | 2.34 | -.29 | .61 | 1.16 | — | — | — | — |
| 7 | Talks too much/loud/fast | 2.30 | -.59 | .39 | 1.09 | — | — | — | — |
| 8 | Racing thoughts | 2.84 | .10 | .94 | 1.62 | 2.64 | .09 | .99 | 1.68 |
| 9 | Jumps topic to topic | 2.78 | .00 | .95 | 1.65 | 2.58 | .00 | .99 | 1.72 |
| 10 | Rushes around nonstop | 2.34 | .08 | .99 | 1.65 | — | — | — | — |
| 11 | Trouble staying on track | 1.25 | -1.27 | .13 | 1.31 | — | — | — | — |
| 12 | Unusually productive/creative | 1.84 | .02 | 1.31 | 2.25 | 1.85 | .01 | 1.31 | 2.25 |
| 13 | Sexually inappropriate behavior | .65 | 2.90 | 4.77 | 5.64 | .58 | 3.18 | 5.24 | 6.19 |
| 14 | Overly outgoing with strangers | 1.30 | 1.29 | 2.60 | 3.92 | — | — | — | — |
| 15 | Foolish or risky behavior | 1.11 | .74 | 2.32 | 3.72 | — | — | — | — |
| 16 | Rage attacks/prolonged tantrums | 1.45 | -.58 | .72 | 1.58 | 1.27 | -.64 | .77 | 1.71 |
| 17 | Excessively silly/jokes/laughs | 1.77 | -.27 | 1.02 | 2.01 | — | — | — | — |
| 18 | Rapid mood swings | 1.54 | -.55 | .64 | 1.39 | — | — | — | — |
| 19 | Suspicious or strange thoughts | 1.16 | .95 | 2.46 | 3.68 | — | — | — | — |
| 20 | Hears voices others don't | .84 | 2.08 | 3.84 | 4.95 | .78 | 2.20 | 4.07 | 5.24 |
| 21 | Sees things others don't | .83 | 2.34 | 4.28 | 5.37 | — | — | — | — |

*Note.* Unidimensional graded response model (mirt, marginal maximum likelihood) fit separately to the clinical sample (*n*=522 with complete CMRS-P-21 data; the 10-item model was fit to the same 522 cases). *a* = discrimination; *b*1–*b*3 = category thresholds on the latent (θ) scale, which is standardized within the clinical sample and is therefore not on the same metric as the general-population table; the two samples are placed on a common metric in Table S15. Items 3, 6, 7, 10, 11, 14, 15, 17, 18, 19 and 21 are not in the 10-item form. Empirical reliability of the θ estimates: .905 (21-item), .809 (10-item). Item 13 is the weakest discriminator in this sample (*a* = 0.65).

**Table S13**

*Item-Level Fit (RMSD) for the Single-Factor Graded Response Model: General Population vs. Clinical Sample, in Descending Order of General-Population RMSD*

| Item | General population RMSD | Clinical RMSD | Difference (clinical − general) | General population at n = 522: median [2.5th, 97.5th pct.] | Substantively different across samples? |
|---|---|---|---|---|---|
| ***Panel A. CMRS-P-21 single-factor GRM (general population n = 2,229; clinical n = 522)*** | | | | | |
| 11 | .119† | .101† | −.019 | .126 [.072, .161] | No — flagged in both samples |
| 2 | .109† | .098 | −.011 | .119 [.078, .169] | No — general population only |
| 7 | .098 | .084 | −.014 | .099 [.057, .152] | No |
| 9 | .084 | .073 | −.011 | .085 [.055, .127] | No |
| 12 | .082 | .076 | −.006 | .077 [.048, .128] | No |
| 1 | .070 | .048 | −.022 | .090 [.061, .134] | No |
| 5 | .067 | .067 | −.000 | .076 [.047, .109] | No |
| 8 | .062 | .062 | +.000 | .072 [.046, .125] | No |
| 18 | .059 | .112† | +.053 | .069 [.046, .095] | Yes — misfit in clinical sample |
| 16 | .056 | .123† | +.067 | .064 [.040, .101] | Yes — misfit in clinical sample |
| 17 | .056 | .082 | +.026 | .071 [.048, .105] | No |
| 6 | .050 | .100 | +.050 | .070 [.044, .102] | No |
| 19 | .047 | .066 | +.019 | .047 [.028, .079] | No |
| 10 | .043 | .058 | +.015 | .053 [.037, .076] | No |
| 20 | .041 | .091 | +.049 | .044 [.021, .069] | Larger in clinical; below 0.10 |
| 14 | .041 | .087 | +.046 | .052 [.029, .078] | Larger in clinical; below 0.10 |
| 3 | .038 | .099 | +.061 | .068 [.040, .104] | No |
| 21 | .037 | .045 | +.008 | .042 [.018, .064] | No |
| 13 | .035 | .080 | +.045 | .041 [.022, .071] | Larger in clinical; below 0.10 |
| 15 | .035 | .120† | +.084 | .044 [.027, .072] | Yes — misfit in clinical sample |
| 4 | .024 | .074 | +.050 | .037 [.017, .070] | Larger in clinical; below 0.10 |
| ***Panel B. CMRS-P-10 single-factor GRM (same cases as Panel A)*** | | | | | |
| 2 | .167† | .203† | +.035 | .157 [.078, .212] | No — flagged in both samples |
| 1 | .136† | .100 | −.036 | .142 [.099, .199] | No — general population only |
| 9 | .126† | .147† | +.021 | .127 [.096, .169] | No — flagged in both samples |
| 8 | .115† | .117† | +.002 | .115 [.096, .151] | No — flagged in both samples |
| 16 | .108† | .152† | +.044 | .096 [.065, .132] | Yes — misfit in clinical sample |
| 5 | .092 | .145† | +.053 | .101 [.073, .142] | Yes — misfit in clinical sample |
| 12 | .090 | .094 | +.004 | .103 [.080, .129] | No |
| 20 | .065 | .111† | +.047 | .061 [.047, .081] | Yes — misfit in clinical sample |
| 4 | .056 | .102† | +.046 | .055 [.036, .084] | Yes — misfit in clinical sample |
| 13 | .053 | .075 | +.023 | .053 [.037, .078] | No |

*Note.* RMSD = root mean square deviation between observed mean item scores and the model-expected item scores within ten expected-a-posteriori θ quantile bins; † = RMSD > 0.10 (the protocol's misfit flag). RMSD computed on the smaller clinical sample (*n*=522) runs higher than RMSD computed on the general population (*n*=,229) even when the model fits equally well, because bin means are noisier. The '*n*=522' column therefore gives the median and 2.5th–97.5th percentiles of general-population RMSD across 100 random subsamples of *n*=522. 'Yes' = clinical RMSD > 0.10 and above that matched-*n* range (misfit that is specific to the clinical sample); 'Larger in clinical; below 0.10' = above the matched-*n* range but not above the 0.10 flag. Differences that fall inside the matched-*n* range are not treated as substantive. The 10-item model was fit to the same cases as the 21-item model.

**Table S14**

*Graded Response Model Item Parameters: CMRS-P-21 and CMRS-P-10 (General-Population Sample)*

| # | Item | CMRS-P-21 model | | | | CMRS-P-10 model | | | |
|---|---|---|---|---|---|---|---|---|---|
| | | ***a*** | ***b*1** | ***b*2** | ***b*3** | ***a*** | ***b*1** | ***b*2** | ***b*3** |
| 1 | Super happy / on top of the world | 1.24 | .01 | 1.87 | 3.44 | 1.32 | .00 | 1.79 | 3.29 |
| 2 | Irritable, cranky, or mad | 1.26 | -.09 | 1.96 | 3.38 | 1.40 | -.09 | 1.84 | 3.17 |
| 3 | Grandiose abilities for age | 1.53 | 1.01 | 2.24 | 3.41 | — | — | — | — |
| 4 | Unrealistic powers, acts on them | 2.51 | 1.46 | 2.23 | 3.10 | 2.94 | 1.39 | 2.13 | 3.00 |
| 5 | Needs less sleep, not tired | 1.56 | .65 | 2.16 | 3.41 | 1.67 | .62 | 2.08 | 3.28 |
| 6 | Periods of too much energy | 2.23 | .09 | 1.50 | 2.45 | — | — | — | — |
| 7 | Talks too much/loud/fast | 2.10 | .10 | 1.43 | 2.29 | — | — | — | — |
| 8 | Racing thoughts | 2.17 | .41 | 1.62 | 2.39 | 2.33 | .40 | 1.58 | 2.32 |
| 9 | Jumps topic to topic | 2.04 | .25 | 1.54 | 2.50 | 1.92 | .26 | 1.58 | 2.58 |
| 10 | Rushes around nonstop | 2.76 | .63 | 1.66 | 2.47 | — | — | — | — |
| 11 | Trouble staying on track | 1.27 | -.51 | 1.44 | 2.69 | — | — | — | — |
| 12 | Unusually productive/creative | 1.73 | .37 | 2.04 | 2.87 | 1.75 | .36 | 2.02 | 2.84 |
| 13 | Sexually inappropriate behavior | 2.10 | 1.73 | 2.58 | 3.19 | 1.98 | 1.76 | 2.65 | 3.30 |
| 14 | Overly outgoing with strangers | 2.18 | 1.32 | 2.18 | 3.10 | — | — | — | — |
| 15 | Foolish or risky behavior | 2.65 | 1.18 | 2.10 | 2.78 | — | — | — | — |
| 16 | Rage attacks/prolonged tantrums | 1.90 | .93 | 2.14 | 2.95 | 1.73 | .98 | 2.22 | 3.08 |
| 17 | Excessively silly/jokes/laughs | 1.85 | .65 | 1.97 | 2.94 | — | — | — | — |
| 18 | Rapid mood swings | 2.05 | .41 | 1.86 | 2.81 | — | — | — | — |
| 19 | Suspicious or strange thoughts | 2.48 | 1.16 | 2.29 | 3.09 | — | — | — | — |
| 20 | Hears voices others don't | 2.64 | 1.78 | 2.27 | 3.24 | 2.49 | 1.80 | 2.30 | 3.35 |
| 21 | Sees things others don't | 2.86 | 1.65 | 2.28 | 2.89 | — | — | — | — |

**Table S15**

*Population-Type DIF, Partial Invariance, and Total-Score Practical Impact (CMRS-P-21, General Population vs. Clinical Sample)*

| ***Panel A. Items Flagged for Population-Type DIF (3 of 21; lordif, $R^2$ change ≥ .02, α=.01; weighted)*** | | | | |
|---|---|---|---|---|
| **Content (Item)** | **$R^2$ change** | **Signed area** | **Unsigned area** | **DIF type** |
| Rage attacks/prolonged tantrums (16) | .041 | +.30 | .30 | Higher; uniform |
| Hears voices others don't (20) | .029 | −.26 | .32 | Lower; mostly uniform |
| Sees things others don't (21) | .034 | −.37 | .42 | Lower; uniform |

| ***Panel B. Configural/Metric/Scalar Invariance, Full vs. Partial (Unidimensional IRT Graded Response Model)*** | | | | |
|---|---|---|---|---|
| **Model** | **CFI** | **RMSEA** | **ΔCFI vs. metric** | **Verdict** |
| Configural | .952 | .063 | — | — |
| Metric (slopes constrained) | .943 | .067 | — | — |
| Scalar, full (all 21-items constrained) | .923 | .073 | −.020 | Fails (>±.010) |
| Scalar, partial (18 anchors; 3 DIF items freed) | .931 | .070 | −.012 | Fails (>±.010) |

| ***Panel C. Total-Score Practical Impact (Differential Test Functioning)*** | | |
|---|---|---|
| Mean \|expected total-score difference\| (θ = −3 to 3) | **0.56 points** | 0.42 within θ = ±2; clinical − general latent mean = 0.67 SD |
| Largest difference, clinical higher / clinical lower | **+0.82 / −2.42 points** | Net bias about 6% of the 6.97-point observed difference in mean totals |
| For reference: main-text Table 2's 95% Dependability RCI | **14.47 points** | all differences below this |
| For reference: main-text Table 2's MID | **4.50 points** | all differences below this |

***Note.*** Panel A: lordif DIF (GRM, $R^2$ change ≥ .02, α=.01) using weighted general-population sample (weight=1 for the clinical sample). Signed/unsigned area = mean and mean absolute difference in expected item score (clinical minus general) over θ = −4 to 4, computed from the partial-scalar model's group-specific item parameters on a common latent metric (general-population *N*=2,229 vs. clinical *N*=522). A positive signed area means the item runs higher in the clinical sample at the same latent severity. Direction: uniform if |signed|/unsigned ≥ .85, mostly uniform if ≥ .70, otherwise partly crossing.

Panel B: weighted fit indices are medians over 10 pseudo-populations drawn in proportion to the weights at the effective sample size, combined with all clinical respondents, configural/metric/scalar based on the unidimensional GRM; partial model anchors the 18 non-flagged items and freely estimates the 3 DIF items per group.

Panel C: differential test functioning (DTF) — expected total score at a given θ from the partial-scalar model (18 anchors identical across groups by construction; group means and variances free, so both groups share one latent metric (general-population mean 0, SD 1)), compared across θ = −3 to 3 in 0.25 steps; the net bias is the difference averaged with the clinical group's latent normal density as weights.

**Table S16**

*Criterion-Related Validity Comparison: CMRS-P-21 vs. CMRS-P-10 (General-Population Sample, Listwise Deletion)*

| Measure | *N* | CMRS-P-21 *r* | CMRS-P-10 *r* | Delta *r* | Cohen's *q* | Steiger's *z* | *p* (FDR) |
|---|---|---|---|---|---|---|---|
| VADPRS Hyperactive Symptoms | 2169 | .63 | .54 | .09 | .13 | 11.89 | <.001*** |
| PSC-17 Total | 2169 | .62 | .56 | .06 | .10 | 8.62 | <.001*** |
| EDI-Reactivity-SF | 2169 | .62 | .58 | .05 | .07 | 6.32 | <.001*** |
| VADPRS Symptoms | 2169 | .61 | .52 | .08 | .12 | 11.41 | <.001*** |
| CHRT-16 | 2169 | .60 | .58 | .03 | .05 | 4.05 | <.001*** |
| EDI-Dysphoria | 2169 | .60 | .59 | .01 | .02 | 1.94 | .061 |
| VQIDS-P-A | 2169 | .57 | .54 | .03 | .04 | 4.11 | <.001*** |
| PSC-17 Externalizing | 2169 | .55 | .49 | .06 | .08 | 7.97 | <.001*** |
| PSC-17 Attention | 2169 | .54 | .46 | .08 | .10 | 9.70 | <.001*** |
| VADPRS Inattentive Symptoms | 2169 | .51 | .43 | .07 | .09 | 8.97 | <.001*** |
| PROMIS-SRI | 2169 | .49 | .48 | .01 | .01 | 1.32 | .197 |
| N-Gauge | 2169 | .48 | .46 | .02 | .02 | 2.42 | .019* |
| PSC-17 Internalizing | 2169 | .45 | .44 | .01 | .02 | 1.66 | .108 |
| NQLS | 2169 | .30 | .27 | .03 | .03 | 3.40 | .001** |
| FIRM | 2169 | .28 | .27 | .01 | .01 | .79 | .429 |
| VADPRS Performance | 2169 | .26 | .22 | .05 | .05 | 5.17 | <.001*** |
| VADPRS Social Performance | 2169 | .25 | .21 | .04 | .04 | 4.09 | <.001*** |
| VADPRS Academic Performance | 2169 | .21 | .17 | .04 | .04 | 4.81 | <.001*** |
| PDS Girl | 1029 | .03 | .06 | -.04 | -.04 | -2.74 | .009** |
| PDS Total | 2169 | .01 | .05 | -.04 | -.04 | -4.54 | <.001*** |
| PDS Boy | 1140 | .00 | .03 | -.03 | -.03 | -2.62 | .012* |

Note: *p* (FDR) values were adjusted using the Benjamini–Hochberg procedure. **p* < .05, ** *p* < .01, *** *p* < .001; no marker = not significant. Since the NQLS is reverse keyed (higher score = better quality of life), values were flipped to positive for viewing purposes of this table. Listwise deletion: all rows are based on the same 2,169 parents who had complete CMRS-P-21 and CMRS-P-10 totals and complete data on all non-sex-specific criteria; the PDS Girl (*n*=1,029) and PDS Boy (*n*=1,140) rows use the girls and boys within that sample. Pearson *r*, Cohen's *q* (difference between Fisher-transformed correlations) and $\Delta r$ are reported to two decimals; $\Delta r$ and *q* are computed from unrounded correlations. Steiger's *z* tests the difference between dependent, overlapping correlations. Rows are ordered by the CMRS-P-21 correlation. General-population correlations are weighted Pearson correlations

(YouGov post-stratification weights); Steiger's *z* and the FDR adjustment use the Kish effective sample size (about 1,125 for the common sample of 2,169 parents). This listwise analysis differs from Tables S17a–c, which use per-criterion complete cases (see the notes to Tables S17a–c); for example, the 21- versus 10-item difference for EDI-Dysphoria is $q = .022$ ($p = .053$, $p_{FDR} = .061$) here and $q = .028$ ($p_{FDR} = .016$) in Table S17b.

**Table S17a**

*Criterion-Related Validity by Scoring Method (General-Population Sample)*

| Measure | n | *r* 21UW | *r* 21EAP | *r* 10UW | *r* 10EAP |
|---|---|---|---|---|---|
| NQLS | 2228 | -.304 | -.291 | -.275 | -.271 |
| PDS Total | 2229 | .011 | -.010 | .050 | .038 |
| VADPRS Symptoms | 2211 | .604 | .579 | .519 | .507 |
| FIRM | 2229 | .277 | .265 | .271 | .258 |
| PSC-17 Total | 2220 | .623 | .599 | .559 | .543 |
| PROMIS-SRI | 2214 | .496 | .454 | .485 | .450 |
| N-Gauge | 2229 | .484 | .460 | .463 | .443 |
| EDI-D | 2229 | .601 | .537 | .583 | .530 |
| VQIDS-P-A | 2228 | .565 | .532 | .533 | .516 |
| CHRT-16 | 2214 | .600 | .563 | .570 | .537 |
| Median \|*r*\| (10 criteria) | | .530 | .496 | .502 | .479 |

Note: Pearson correlations (*r*). UW = unit-weighted (raw sum) total score. EAP = expected a posteriori theta score from a single-factor graded response model, fit separately for the 21-item and 10-item item sets. All correlations in Tables S17a–c are weighted Pearson correlations (YouGov post-stratification weights); the EAP scores come from graded response models calibrated with the same weights. Unlike Table S16, which is listwise (n = 2,169 parents complete on both CMRS-P forms and on all 19 non-sex-specific criteria), each row here uses per-criterion complete cases: every parent with a complete CMRS-P-21 (hence a complete CMRS-P-10) and a score on that criterion, with no deletion on the other criteria (n = 2,211–2,229, as shown). Estimates therefore differ slightly from Table S16; for VQIDS-P-A the correlation with the 21-item total is .565 here (n = 2,228) and .568 in Table S16 (n = 2,169).

**Table S17b**

*Pairwise Correlation Comparisons Across CMRS-P Forms and Scoring Methods (Cohen's q, Steiger's Z)*

| Measure | 21 vs 10 (UW) | 21 vs 10 (EAP) | EAP vs UW (21-item) | EAP vs UW (10-item) |
|---|---|---|---|---|
| NQLS | -.032*** | -.021 | .015 | .004 |
| PDS Total | -.040*** | -.048*** | -.021 | -.012 |
| VADPRS Symptoms | .126*** | .103*** | -.038** | -.015 |
| FIRM | .007 | .007 | -.014 | -.014 |
| PSC-17 Total | .099*** | .082*** | -.039** | -.023 |
| PROMIS-SRI | .014 | .005 | -.055*** | -.046*** |
| N-Gauge | .027** | .021 | -.031* | -.025* |
| EDI-D | .028* | .009 | -.095*** | -.077*** |
| VQIDS-P-A | .046*** | .021 | -.048*** | -.022 |
| CHRT-16 | .046*** | .036** | -.057*** | -.048*** |
| Median *q*, NQLS sign-corrected (Wilcoxon) | .030* | .021* | -.039** | -.023** |

Note. Each cell is Cohen's *q* (Fisher *z*-difference) for the labeled pairwise comparison, via Steiger's *Z* for dependent, overlapping correlations sharing one variable in common. UW = unit-weighted (raw sum) total score. EAP = expected a posteriori theta score. *$p$_FDR<.05, **$p$_FDR<.01, ***$p$_FDR<.001; no marker = not significant after correction. Correlations are weighted. Steiger's z uses the Kish effective sample size of each row's analytic sample (about 1,156 for the per-criterion samples of 2,228–2,229 parents), and p-values are adjusted (Benjamini–Hochberg) within each column across the 10 criteria shown. Table S16 adjusts across 21 criteria in the listwise sample (effective n about 1,125), so a difference can be significant here but not there: for EDI-Dysphoria the 21- versus 10-item comparison (unit-weighted totals) is $q = .028$, $p_{FDR} = .016$ here but $q = .022$, $p_{FDR} = .061$ in Table S16.

**Table S17c**

*Correlation Comparisons via Steiger's Z*

| Comparison | $r_1$ | $r_2$ | Shared/related *r* | Steiger's *Z* | *p* |
|---|---|---|---|---|---|
| CMRS-P-21 unit-weighted total vs. CMRS-P-21 EAP theta (both vs. VQIDS-P-A, baseline) | *r = .565* | *r = .532* | (shared criterion) | not separately tested (see Table S17b) | *p* < .001 |
| CMRS-P-21 unit-weighted vs. CMRS-P-21 EAP theta (both vs. VQIDS-P-A, retest, *N* = 167) | *r = .342* | *r = .408* | (shared criterion) | *z* = −1.44 | *p = .151* |
| CMRS-P-21 full-form total vs. CMRS-P-10 brief-form total (both vs. VQIDS-P-A, baseline, *N* = 2,228) | *r = .565* | *r = .533* | *r = .952 (P21-P10 total)* | *z* = 4.267 | *p < .001* |

Note. All correlations are weighted Pearson correlations. Rows 1 and 3 use the 2,228 parents with a complete CMRS-P-21 and a complete VQIDS-P-A (per-criterion complete cases, effective n about 1,156); row 2 uses the 167 retest pairs with their baseline weights (effective n = 81). Steiger's z uses the effective sample size. Row 3 repeats the VQIDS-P-A comparison reported in Tables S16 (listwise, n = 2,169: r = .568 vs. .537, z = 4.11), S17a–b (per-criterion, n = 2,228: r = .565 vs. .533, q = .046) and S18.

**Table S18**

*CMRS-P-10 Brief Form: Psychometrics and Concordance With the CMRS-P-21 (General Population, Complete Cases, N = 2,229)*

| Statistic | Value |
|---|---|
| *N (complete cases on the 21-item scale, hence on all 10 brief-form items)* | 2,229 |
| *N (complete on the 10 brief-form items alone; not used here)* | 2,240 |
| Total score, weighted *M* (*SD*), *N* = 2,229 (effective *N* = 1,156) | 4.05 (4.32) |
| Total score range (potential 0–30) | 0–28 observed |
| 1-factor model fit ($\chi^2$, *df*, CFI, TLI, RMSEA, SRMR) | $\chi^2$ = 253.938, *df* = 35, CFI = .982, TLI = .977, RMSEA = .053 [.047, .059], SRMR = .074 |
| 2-factor correlated model fit ($\chi^2$, *df*, CFI, TLI, RMSEA, SRMR) | $\chi^2$ = 117.586, *df* = 34, CFI = .993, TLI = .991, RMSEA = .033 [.027, .040], SRMR = .050 |
| ECV (general factor) | .74 (target ≥ .70) |
| omega_H (general factor) | .77 (target ≥ .70–.80) |
| Maximum omega_HS (either group factor) | .22 (elevated if ≥ .30) |
| Specific-factor gate outcome | Standard (essentially unidimensional; no elevated group factor) |
| Empirical IRT reliability (GRM, EAP) | .768 |
| Correlation with CMRS-P-21 full-form total (weighted, *N* = 2,229) | *r* = .952 |
| Correlation with VQIDS-P-A criterion, baseline (weighted, *N* = 2,228; effective *N* = 1,156) | r = .533 (vs. CMRS-P-21 full-form *r = .565*; Steiger's *z* = 4.267, *p* < .001 — see Table S17c) |

**Table S19**

*Comparison of AIC, BIC, and Degrees of Freedom (df) values across GAMLSS distribution families*

| | | **CMRS-P-21** | | | **CMRS-P-10** | | |
|---|---|---|---|---|---|---|---|
| **GAMLSS Distribution Family** | **Type** | **AIC** | **BIC** | ***df*** | **AIC** | **BIC** | ***df*** |
| Zero-Adjusted Negative Binomial type I (ZANBI) | D | 14247.04 | 14281.29 | 6.00 | 11176.91 | 11211.16 | 6.00 |
| Negative Binomial type II (NBII) | D | 14253.38 | 14281.93 | 5.00 | 11176.87 | 11205.42 | 5.00 |
| **Negative Binomial type I (NBI)** | **D** | **14255.30** | **14283.84** | **5.00** | **11172.23** | **11200.78** | **5.00** |
| Zero-Inflated Poisson Inverse Gaussian (ZIPIG) | D | 14256.10 | 14290.35 | 6.00 | 11160.83 | 11195.09 | 6.00 |
| Zero-Inflated Negative Binomial type I (ZINBI) | D | 14257.30 | 14291.55 | 6.00 | 11173.04 | 11207.30 | 6.00 |
| Zero-Adjusted Poisson Inverse Gaussian (ZAPIG) | D | 14258.94 | 14293.20 | 6.00 | 11163.48 | 11197.74 | 6.00 |
| Zero-Adjusted Beta Binomial (ZABB) | D | 14323.12 | 14357.37 | 6.00 | 11237.15 | 11271.40 | 6.00 |
| Poisson Inverse Gaussian (PIG) | D | 14334.19 | 14362.73 | 5.00 | 11220.78 | 11249.32 | 5.00 |
| Beta Binomial (BB) | D | 14334.53 | 14363.08 | 5.00 | 11245.98 | 11274.53 | 5.00 |
| Zero-Adjusted Poisson (ZAP) | D | 22936.94 | 22965.48 | 5.00 | 13353.89 | 13382.43 | 5.00 |
| Zero-Inflated Poisson (ZIP) | D | 22936.96 | 22965.50 | 5.00 | 13351.00 | 13379.55 | 5.00 |
| Poisson (PO) | D | 25218.74 | 25241.58 | 4.00 | 14767.19 | 14790.02 | 4.00 |

Note: Only discrete (integer-valued) candidate families were fit, to the complete cases ($N$=2,229), with the YouGov weights entered as prior weights (rescaled to sum to $N$), a sex main effect, linear age, an age-by-sex interaction on $\mu$, constant $\sigma$, and (where applicable) constant $\nu$. AIC = Akaike Information Criterion; BIC = Bayesian Information Criterion; *df* = effective degrees of freedom. Type: D = discrete (the zero-inflated and zero-adjusted families are discrete with an added mass at zero). NC = did not converge. † = converged with an additive-fit convergence warning. Families are ordered by weighted CMRS-P-21 BIC; weighted AIC/BIC are comparable only with each other. Bold = selected model (NBI for both forms). ZANBI (CMRS-P-21) and ZIPIG (CMRS-P-10) had the lowest BICs, by 2.6 and 5.7 points over NBI; the Q-statistics from 25 pseudo-population refits (drawn in proportion to the weights at the effective sample size) did not favor them over NBI, and NBI was retained for consistency across forms and with the unweighted analysis. The center Q-statistic (Z1) was significant in most refits for every family, reflecting the linear age function. The weighted diagnostics are a coarser check than the unweighted ones.

**Table S20**

*Cross-Validated Model Selection for the Age and Sex Structure of the Weighted CMRS-P Norms*

| | | CMRS-P-21 | | | CMRS-P-10 | | |
|---|---|---|---|---|---|---|---|
| **Mean structure (NBI, constant σ)** | **df** | **CV log-lik. (SE)** | **Δ vs. best (SE of Δ)** | ***BIC*** | **CV log-lik. (SE)** | **Δ vs. best (SE of Δ)** | ***BIC*** |
| Sex only (no age effect) | 3 | −3.2011 (0.0063) | −0.0018 (0.0006) | 14284.9 | −2.5119 (0.0054) | −0.0040 (0.0008) | 11210.8 |
| Linear age, sex-additive | 4 | −3.2022 (0.0063) | −0.0029 (0.0006) | 14292.6 | −2.5121 (0.0054) | −0.0042 (0.0008) | 11215.0 |
| **Linear age × sex (selected)** | **5** | **−3.1993 (0.0063)** | **0 (best)** | **14283.8** | **−2.5079 (0.0054)** | **0 (best)** | **11200.8** |
| Quadratic age, sex-additive | 5 | −3.2032 (0.0064) | −0.0039 (0.0006) | 14299.6 | −2.5127 (0.0056) | −0.0048 (0.0008) | 11221.0 |
| Quadratic age × sex | 7 | −3.2014 (0.0064) | −0.0021 (0.0002) | 14298.9 | −2.5095 (0.0056) | −0.0016 (0.0002) | 11215.2 |
| Cubic age, sex-additive | 6 | −3.2042 (0.0064) | −0.0049 (0.0006) | 14307.1 | −2.5132 (0.0057) | −0.0053 (0.0009) | 11226.8 |
| Cubic age × sex (preregistered form) | 9 | −3.2027 (0.0064) | −0.0034 (0.0003) | 14313.0 | −2.5105 (0.0056) | −0.0026 (0.0003) | 11228.8 |

Note: Weighted negative binomial (NBI) models with constant dispersion were compared by 10 × 10 repeated, stratified 10-fold cross-validation (folds balanced within year of age by sex), scoring the case-weighted log-likelihood of the held-out scores per observation (higher is better); age was centered at 12 years and scaled by 6 years before polynomial terms were formed. SE = standard error of the mean across the ten folds (each averaged over the ten repeats); Δ vs. best is the paired difference from the best-performing model, with the standard error of that paired difference. BIC is from the weighted fit to all 2,229 complete cases. Selection rule (specified uniformly for both forms): among the linear, quadratic and cubic age functions, choose the least complex model whose cross-validated performance is within one paired standard error of the best, and accept added complexity only if the BIC also improves. Sex-only is shown as a reference. [Specified after the CMRS-P results and before norming the remaining measures; the preregistered cubic age × sex form is reported here and in Table S21 as a sensitivity analysis — confirm wording against the preregistration.]

**Table S21**

*Primary Weighted Norms With Bootstrap Intervals, Sensitivity Analyses, and Effective Sample Size by Age and Sex (95th and 50th Percentiles)*

| Scale and age | Sex | Primary (weighted) | | *Unweighted* | Wtd. cubic | Cov.-std. | *Effective n (n)* |
|---|---|---|---|---|---|---|---|
| | | P95 [95% CI] | P50 [95% CI] | | | | |
| CMRS-P-21, age 6 | Male | 30 [26, 35] | 7 [6, 8] | 30 / 7 | 31 / 7 | 29 / 7 | 66 (80) |
| CMRS-P-21, age 6 | Female | 22 [19, 25] | 5 [4, 6] | 23 / 5 | 21 / 5 | 27 / 6 | 59 (82) |
| CMRS-P-21, age 9 | Male | 28 [25, 31] | 6 [6, 7] | 27 / 6 | 27 / 6 | 28 / 6 | 61 (89) |
| CMRS-P-21, age 9 | Female | 23 [21, 26] | 5 [5, 6] | 23 / 5 | 24 / 6 | 26 / 6 | 48 (67) |
| CMRS-P-21, age 12 | Male | 26 [24, 28] | 6 [5, 7] | 25 / 6 | 26 / 6 | 27 / 6 | 55 (108) |
| CMRS-P-21, age 12 | Female | 25 [23, 28] | 6 [5, 6] | 23 / 5 | 25 / 6 | 25 / 6 | 48 (105) |
| CMRS-P-21, age 15 | Male | 24 [21, 27] | 6 [5, 6] | 23 / 5 | 25 / 6 | 26 / 6 | 56 (104) |
| CMRS-P-21, age 15 | Female | 27 [24, 31] | 6 [5, 7] | 23 / 5 | 27 / 6 | 25 / 5 | 50 (117) |
| CMRS-P-21, age 18 | Male | 23 [18, 27] | 5 [4, 6] | 21 / 5 | 22 / 5 | 26 / 6 | 8 (9) |
| CMRS-P-21, age 18 | Female | 30 [24, 35] | 7 [6, 8] | 23 / 5 | 35 / 8 | 24 / 5 | 9 (12) |
| CMRS-P-10, age 6 | Male | 13 [12, 16] | 3 [3, 4] | 13 / 3 | 14 / 3 | 13 / 3 | 66 (80) |
| CMRS-P-10, age 6 | Female | 10 [8, 11] | 2 [2, 2] | 11 / 2 | 9 / 2 | 13 / 3 | 59 (82) |
| CMRS-P-10, age 9 | Male | 13 [12, 14] | 3 [3, 3] | 12 / 3 | 13 / 3 | 13 / 3 | 61 (89) |
| CMRS-P-10, age 9 | Female | 11 [10, 12] | 2 [2, 3] | 11 / 2 | 11 / 3 | 13 / 3 | 48 (67) |
| CMRS-P-10, age 12 | Male | 12 [11, 13] | 3 [3, 3] | 12 / 3 | 12 / 3 | 13 / 3 | 55 (108) |
| CMRS-P-10, age 12 | Female | 12 [11, 14] | 3 [3, 3] | 12 / 3 | 12 / 3 | 13 / 3 | 48 (105) |
| CMRS-P-10, age 15 | Male | 12 [10, 13] | 3 [2, 3] | 11 / 2 | 12 / 3 | 13 / 3 | 56 (104) |
| CMRS-P-10, age 15 | Female | 14 [12, 16] | 3 [3, 4] | 12 / 3 | 14 / 3 | 13 / 3 | 50 (117) |
| CMRS-P-10, age 18 | Male | 11 [9, 13] | 2 [2, 3] | 11 / 2 | 12 / 3 | 13 / 3 | 8 (9) |
| CMRS-P-10, age 18 | Female | 16 [13, 19] | 4 [3, 5] | 13 / 3 | 20 / 5 | 13 / 3 | 9 (12) |

Note: Primary = survey-weighted negative binomial model, linear age × sex (1,000 bootstrap resamples of respondents; percentile 95% intervals). Unweighted = the same linear age × sex model without weights. Wtd. cubic = the weighted cubic age × sex model (the preregistered functional form). Cov.-std. = covariate-standardized: unweighted NBI model with linear age and sex plus the weighting variables (parent gender, education, Hispanic ethnicity, race, region, and parent age centered within child age), with the predicted score distribution standardized to the weighted national covariate composition (percentiles taken from the mixture distribution). Effective $n = (\Sigma w)^2/\Sigma w^2$ within the year of age and sex; $n$ = unweighted count. At ages 17 and 18 the weighted estimates rest on very few effective respondents, and the female age trend in the primary and cubic models was not reproduced without weights or after covariate standardization.

**Table S22**

*Item Severity Ordering From the Graded Response Model: Mean Threshold ($\bar{b}$) on a Common Metric, General-Population and Clinical Samples*

| Rank | Item | a | b1 | b2 | b3 | General population: $\bar{b}$ [95% CI] | Rank interval | Clinical: $\bar{b}$ [95% CI] | Clinical rank |
|---|---|---|---|---|---|---|---|---|---|
| 1 | Easily distracted (11) | 1.32 | −0.50 | 1.26 | 2.40 | 1.05 [0.93, 1.18] | 1–2 | 1.05 [0.93, 1.18] | 1 |
| 2 | Talks too much/fast (7) | 2.18 | 0.09 | 1.31 | 2.08 | 1.16 [1.08, 1.25] | 1–2 | 1.16 [1.08, 1.25] | 2 |
| 3 | Too much energy (6) | 2.19 | 0.13 | 1.44 | 2.23 | 1.27 [1.18, 1.38] | 3–3 | 1.27 [1.18, 1.38] | 4 |
| 4 | Jumps topic to topic (9) | 2.03 | 0.32 | 1.58 | 2.50 | 1.47 [1.36, 1.59] | 4–7 | 1.47 [1.36, 1.59] | 5 |
| 5 | Rapid mood swings (18) | 2.00 | 0.38 | 1.68 | 2.42 | 1.49 [1.37, 1.63] | 4–7 | 1.49 [1.37, 1.63] | 6 |
| 6 | Racing thoughts (8) | 2.16 | 0.47 | 1.64 | 2.40 | 1.50 [1.38, 1.63] | 4–7 | 1.50 [1.38, 1.63] | 7 |
| 7 | Rushes around nonstop (10) | 2.62 | 0.66 | 1.66 | 2.39 | 1.57 [1.48, 1.70] | 5–8 | 1.57 [1.48, 1.70] | 8 |
| 8 | Irritable, cranky or mad (2) | 1.26 | −0.10 | 1.80 | 3.10 | 1.60 [1.46, 1.77] | 5–9 | 1.60 [1.46, 1.77] | 9 |
| 9 | Excessive joking (17) | 1.92 | 0.60 | 1.84 | 2.77 | 1.74 [1.58, 1.91] | 8–11 | 1.74 [1.58, 1.91] | 10 |
| 10 | Unusually productive (12) | 1.71 | 0.42 | 2.04 | 2.90 | 1.79 [1.64, 1.93] | 9–11 | 1.79 [1.64, 1.93] | 11 |
| 11 | Super happy, on top of the world (1) | 1.19 | 0.10 | 2.03 | 3.53 | 1.88 [1.70, 2.16] | 9–13 | 1.88 [1.70, 2.16] | 12 |
| 12 | Rage attacks/tantrums (16)* | 1.89 | 0.93 | 2.14 | 2.95 | 2.01 [1.80, 2.25] | 11–15 | 1.20 [1.05, 1.34] | 3 |
| 13 | Grandiose ability claims (3) | 1.40 | 0.94 | 2.12 | 3.10 | 2.05 [1.89, 2.25] | 11–15 | 2.05 [1.89, 2.25] | 13 |
| 14 | Foolish/risky behavior (15) | 2.23 | 1.23 | 2.20 | 2.97 | 2.14 [1.97, 2.33] | 12–16 | 2.14 [1.97, 2.33] | 14 |
| 15 | Needs less sleep (5) | 1.40 | 0.80 | 2.35 | 3.50 | 2.22 [2.02, 2.45] | 13–19 | 2.22 [2.02, 2.45] | 15 |
| 16 | Sees things (21)* | 2.83 | 1.66 | 2.28 | 2.89 | 2.27 [2.03, 2.69] | 13–21 | 4.31 [3.28, 6.18] | 21 |
| 17 | Suspicious/strange thoughts (19) | 2.11 | 1.26 | 2.41 | 3.24 | 2.30 [2.09, 2.55] | 15–20 | 2.30 [2.09, 2.55] | 16 |

| | | | | | | | | | |
|---|---|---|---|---|---|---|---|---|---|
| 18 | Overly social with strangers (14) | 1.91 | 1.46 | 2.39 | 3.38 | 2.41 [2.17, 2.69] | 15–21 | 2.41 [2.17, 2.69] | 17 |
| 19 | Hears voices (20)* | 2.62 | 1.78 | 2.27 | 3.24 | 2.43 [2.17, 2.69] | 15–21 | 3.94 [3.04, 5.38] | 20 |
| 20 | Unrealistic powers (4) | 2.10 | 1.63 | 2.45 | 3.26 | 2.45 [2.23, 2.77] | 17–20 | 2.45 [2.23, 2.77] | 18 |
| 21 | Sexually inappropriate (13) | 1.66 | 1.97 | 2.90 | 3.49 | 2.79 [2.49, 3.21] | 19–21 | 2.79 [2.49, 3.21] | 19 |

Note. Graded response model (mirt) fit as a linked partial-scalar multiple-group model: the 18 items without population-type DIF are constrained equal across samples; items 16, 20 and 21 (*) are freed, so their parameters differ by sample. The metric is defined by the general-population sample (mean 0, SD 1; clinical latent mean 0.67 SD higher, SD ratio 0.95). a = discrimination; b1–b3 = thresholds for scores ≥ 1, ≥ 2 and = 3 (general-population calibration); $\bar{b}$ = mean of the three thresholds, the severity index used to order the items (higher = more severe). Intervals are 2.5th–97.5th percentiles of 200 bootstrap resamples (respondents resampled within sample, weights carried); rank intervals are the corresponding percentile ranks of $\bar{b}$ among the 21 items. The general-population sample is weighted. The order of $\bar{b}$ in the two samples is highly similar (Spearman $\rho = .92$) because the 18 anchor items are constrained equal; the differences come from the three freed items.

**Table S23**

*Sex and Age-Stratum Differential Item Functioning (Weighted and Unweighted) for the CMRS-P-21 Items*

| Item | Sex DIF ($R^2$ change) | | Age-stratum DIF ($R^2$ change) | | |
|---|---|---|---|---|---|
| | Weighted | Unweighted | Weighted | Unweighted | Age: % weighted bootstrap flags |
| Super happy, on top of the world (1) | .000 | .001 | .002 | .000 | 0% |
| Irritable, cranky or mad (2) | .006 | .005 | .017 | .032† | 33% |
| Grandiose ability claims (3) | .001 | .002 | .003 | .002 | 0% |
| Unrealistic powers (4) | .003 | .002 | .003 | .001 | 2% |
| Needs less sleep (5) | .001 | .001 | .007 | .006 | 5% |
| Too much energy (6) | .010 | .008 | .017 | .013 | 40% |
| Talks too much/fast (7) | .001 | .000 | .007 | .005 | 2% |
| Racing thoughts (8) | .001 | .000 | .007 | .005 | 0% |
| Jumps topic to topic (9) | .000 | .001 | .002 | .002 | 0% |
| Rushes around nonstop (10) | .003 | .003 | .014 | .011 | 20% |
| Easily distracted (11) | .000 | .000 | .003 | .002 | 0% |
| Unusually productive (12) | .001 | .001 | .001 | .000 | 0% |
| Sexually inappropriate (13) | .009 | .004 | .015 | .009 | 37% |
| Overly social with strangers (14) | .001 | .000 | .011 | .004 | 32% |
| Foolish/risky behavior (15) | .000 | .002 | .002 | .004 | 2% |
| Rage attacks/tantrums (16) | .002 | .001 | .001 | .001 | 0% |
| Excessive joking (17) | .002 | .001 | .003 | .004 | 0% |
| Rapid mood swings (18) | .010 | .008 | .018 | .028† | 60% |
| Suspicious/strange thoughts (19) | .001 | .000 | .011 | .011 | 15% |
| Hears voices (20) | .008 | .006 | .015 | .014 | 52% |
| Sees things (21) | .004 | .005 | .009 | .006 | 38% |

Note. lordif (graded response model; ordinal logistic regression; McFadden pseudo-$R^2$ change for the model with group and group × θ terms vs. θ alone; flag if $R^2$ change ≥ .02 at α = .01) on the 2,229 complete cases; general-population weights in the weighted columns. Sex: boys (n = 1,171) vs. girls (n = 1,058). Age strata: 6–9 (n = 675), 10–12 (n = 531) and 13–18 years (n = 1,023), a three-group comparison. † = flagged ($R^2$ change ≥ .02). Bootstrap column: percentage of 60 weighted bootstrap resamples (respondents resampled with their weights) in which the item was flagged for age DIF; no item was flagged for sex DIF in more than 10% of 60 resamples. Items are unflagged under weighting for both facets; items 2 and 18 are flagged by age without weights.

**Figure S1a**

*CMRS-P-21 Worm Plot, Negative Binomial Type I (NBI) Distribution, Survey-Weighted Linear Age × Sex Model*

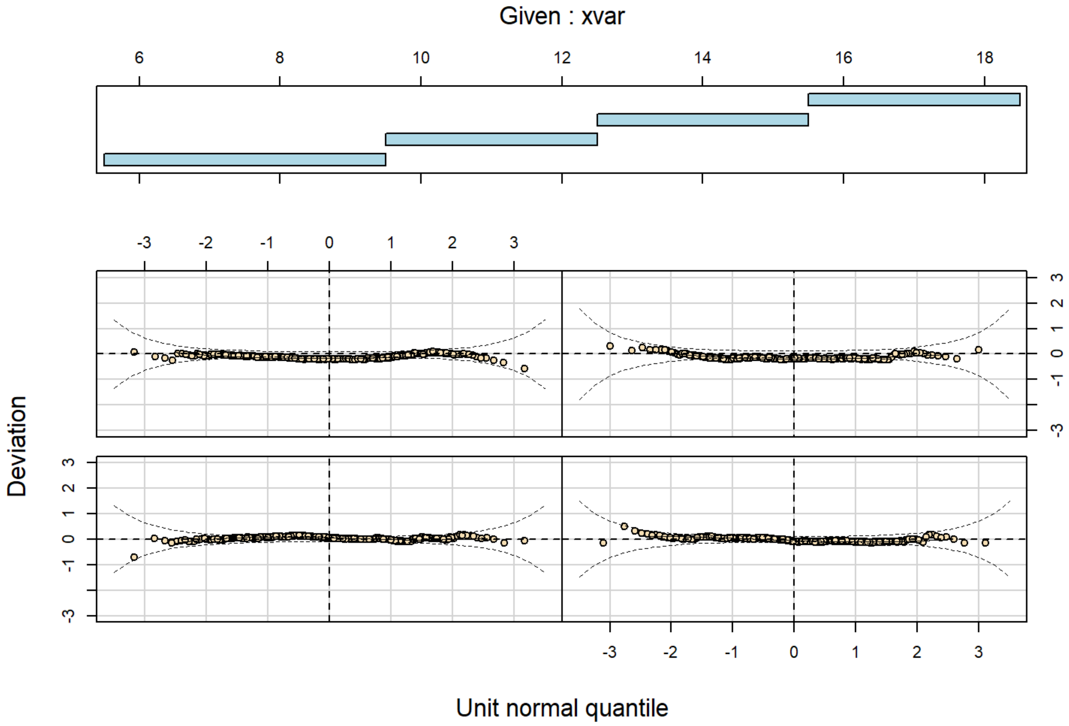


**Figure S1b**

*CMRS-P-10 Worm Plot, Negative Binomial Type I (NBI) Distribution, Survey-Weighted Linear Age × Sex Model*

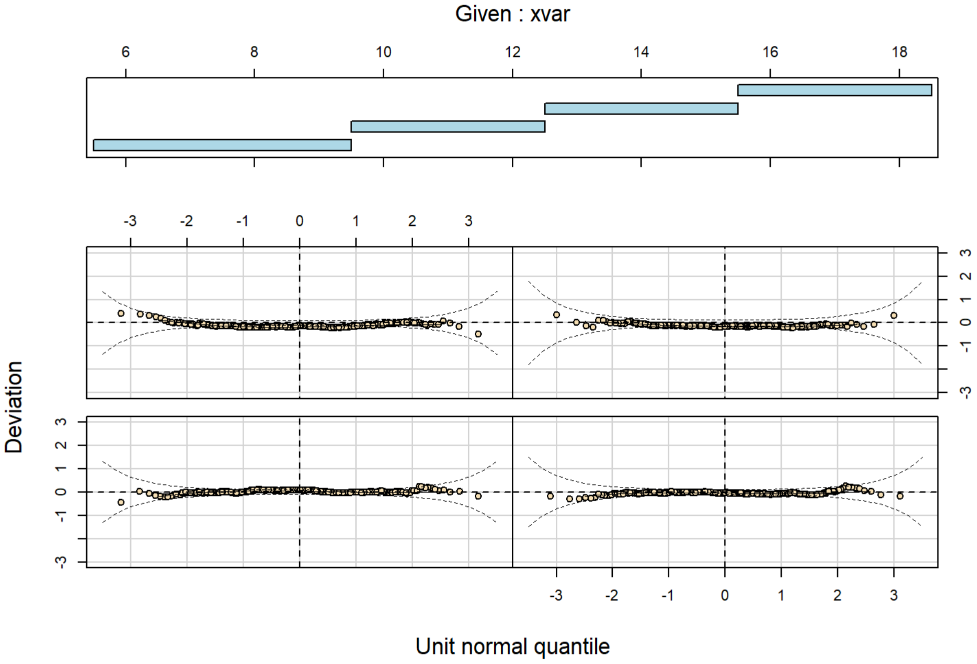

**Figure S2a**

*CMRS-P-21 Centile Chart by Sex, Negative Binomial Type I (NBI) Distribution, Survey-Weighted Linear Age × Sex Model*

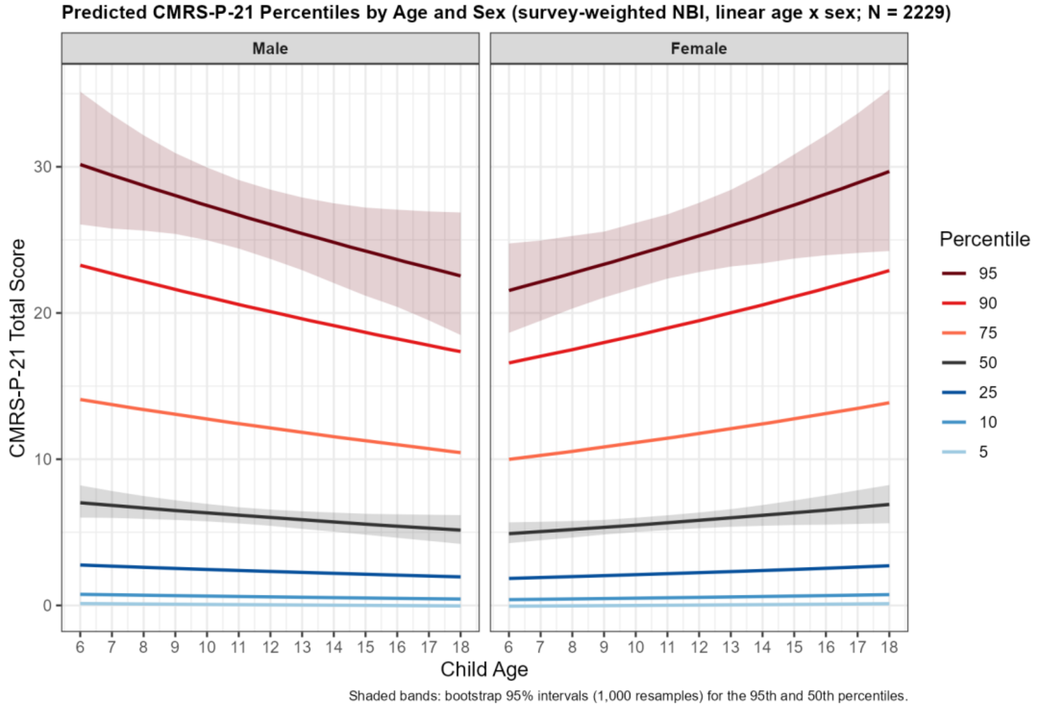


**Figure S2b**

*CMRS-P-10 Centile Chart by Sex, Negative Binomial Type I (NBI) Distribution, Survey-Weighted Linear Age × Sex Model*

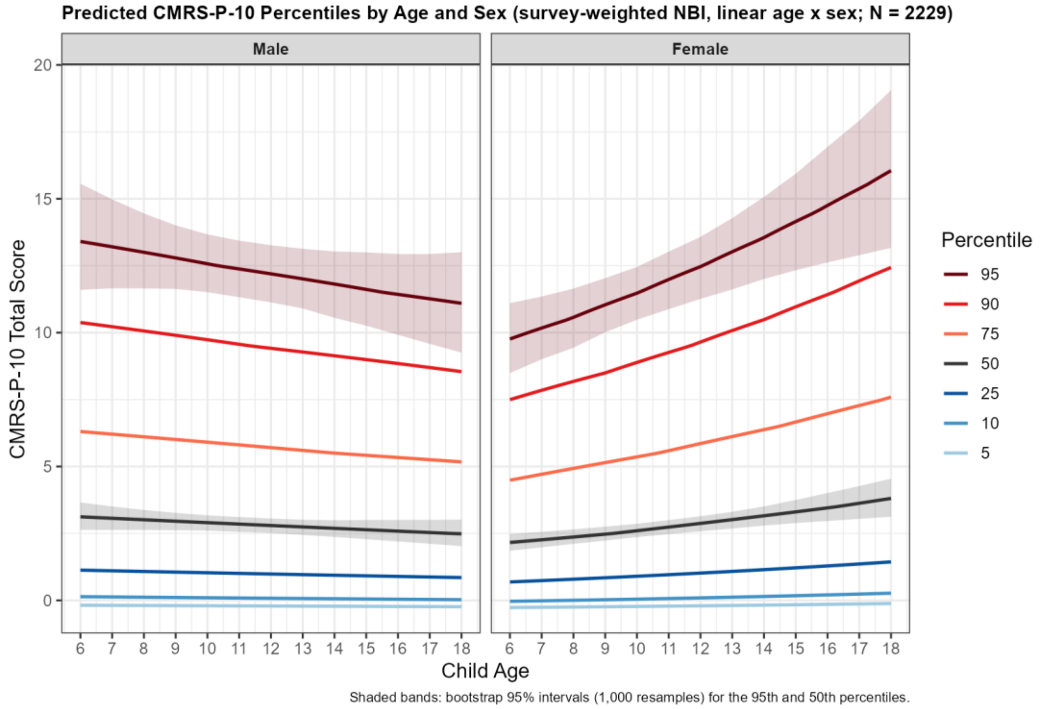

**Figure S3**

*Primary Weighted Norms and Sensitivity Analyses: 95th and 50th Percentiles by Age and Sex*

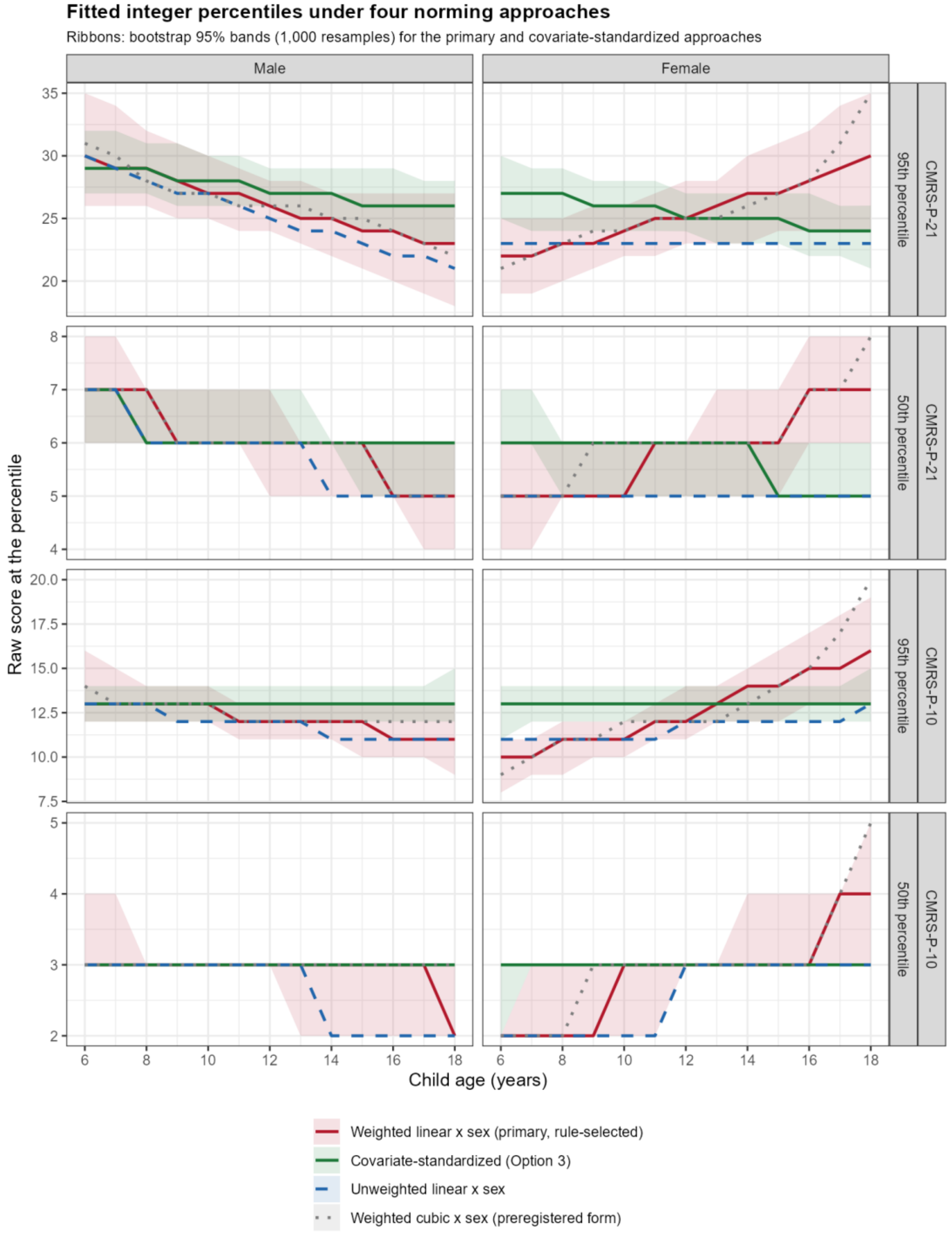


Note. Percentile lines from the survey-weighted linear age × sex NBI model (primary), the same model unweighted, the weighted cubic age × sex model, and the covariate-standardized model; ribbons show bootstrap 95% intervals (1,000 resamples) for the primary and covariate-standardized norms. Scores are integers, so fitted percentiles are step functions of the integer scale.

#### Figure S4. Higher-Order Factor Structure (General-Population/BENCHMARK Sample)

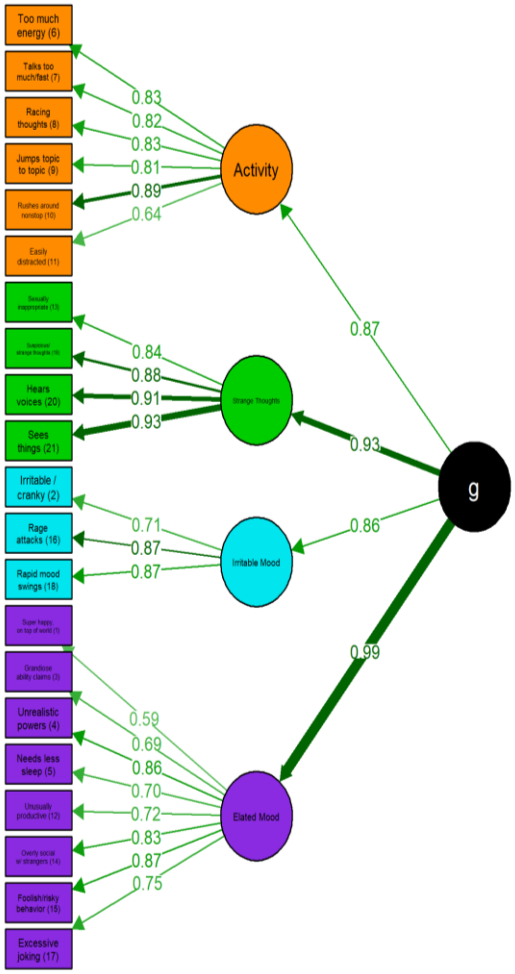


Note. Standardized path diagram of the retained higher-order CFA model — four first-order factors (Elated Mood, Activity, Irritable Mood, Strange Thoughts & Behaviors) loading onto a single second-order general factor (g) — BENCHMARK (general-population) sample. Complete cases (N=2,229); item 13 is assigned to Strange Thoughts & Behaviors.
